# Large-Scale Gene-Smoking Interactions and Fine Mapping Study Identifies Multiple Novel Blood Pressure Loci in over 1 Million Individuals

**DOI:** 10.1101/2025.10.06.25337440

**Authors:** Mengyu Zhang, Michael R. Brown, Amy R. Bentley, Thomas W. Winkler, Raymond Noordam, Pavithra Nagarajan, Bohong Guo, Songmi Lee, Karen Schwander, Wenyi Wang, Kenneth Westerman, Jeffrey R. O’Connell, Farah Ammous, Anna D. Argoty-Pantoja, Traci M. Bartz, Chiara Batini, Palwende R. Boua, Heather J. Cordell, Laia Díez-Ahijado, Latchezar Dimitrov, Anh N. Do, Jiawen Du, Mary F. Feitosa, Ayush Giri, Franco Giulianini, Valborg Gudmundsdottir, Xiuqing Guo, Sarah E. Harris, Natalie R. Hasbani, Janina M. Herold, Keiko Hikino, Edith Hofer, Fang-Chi Hsu, Anne U. Jackson, Minjung Kho, Aldi T. Kraja, Leo-Pekka Lyytikäinen, Aline Meirhaeghe, Manon Muntaner, Masahiro Nakatochi, Giuseppe Giovanni Nardone, Teresa Nutile, Nicholette D. Palmer, Alessandro Pecori, Varun S. Rao, Rainer Rauramaa, Mihir M. Sanghvi, Aurora Santin, Botong Shen, Heather M. Stringham, Fumihiko Takeuchi, Ye An Tan, Jingxian Tang, Sébastien Thériault, Olga Trofimova, Stella Trompet, Peter J. van der Most, Ya Xing Wang, Zhe Wang, Yujie Wang, Erin B. Ware, Stefan Weiss, Ananda Rajitha Wickremasinghe, Chenglong Yu, Wanying Zhu, Md Abu Yusuf Ansari, Pramod Anugu, Bernhard Banas, R. Graham Barr, Til B. Basnet, Eric Boerwinkle, Carsten A. Böger, Max Breyer, Ulrich Broeckel, Luke Bryant, Brian Cade, Silvia Camarda, Pasqualina Cennamo, William Checkley, Miao-Li Chee, Guanjie Chen, Kayesha Coley, Stacey D. Collins, Martin H. de Borst, Lisa de las Fuentes, Ian J. Deary, Charles T. Dupont, Christian Enzinger, Tariq Faquih, Jessica D. Faul, Lilian Fernandes Silva, Victoria Gauthier, Adam D. Gepner, Mathias Gorski, Hans Jörgen Grabe, Mariaelisa Graff, Charles C. Gu, Jiang He, Sami Heikkinen, Bertha Hidalgo, Heather M. Highland, James E. Hixson, Michelle M. Hood, Steven C. Hunt, Marguerite Ryan Irvin, Masato Isono, Mika Kähönen, Sharon L. R Kardia, Carrie A. Karvonen-Gutierrez, Anuradhani Kasturiratne, Tomohiro Katsuya, Joel D. Kaufman, Heikki A. Koistinen, Pirjo Komulainen, Bernhard K. Krämer, Lenore J. Launer, Hampton Leonard, Daniel Levy, Lifelines Cohort Study, Jianjun Liu, Pedro Marques-Vidal, Angel Martinez-Perez, Koichi Matsuda, John J. McNeil, Yuri Milaneschi, J. Jaime Miranda, John L. Morrison, Michael Nalls, Maggie CY Ng, Ilja M. Nolte, Jill M. Norris, Anniina Oravilahti, Amit Patki, Lauren E. Petty, Patricia A. Peyser, Giulia Pianigiani, Laura M. Raffield, Olli T. Raitakari, Michele Ramsay, Ken M. Rice, Paul M. Ridker, Lorenz Risch, Martin Risch, Daniela Ruggiero, Edward A. Ruiz-Narvaez, Tom C. Russ, Charumathi Sabanayagam, Nataraja Sarma Vaitinadin, Reinhold Schmidt, Laura J. Scott, Surina Singh, Colleen M. Sitlani, Roelof A.J Smit, Jennifer A. Smith, Quan Sun, E Shyong Tai, Kent D. Taylor, Paola Tesolin, Yih Chung Tham, The Biobank Japan Project, Chikowore Tinashe, Rob M. van Dam, Julien Vaucher, Uwe Völker, Chaolong Wang, Otis D. Wilson, Tien-Yin Wong, Jianzhao Xu, Ken Yamamoto, Jie Yao, Mitsuhiro Yokota, Kristin L. Young, Martina E. Zimmermann, Philippe Amouyel, Jennifer E. Below, Sven Bergmann, Antonio Bernabe-Ortiz, Laura Bierut, Michael Boehnke, Donald W. Bowden, Jean-Tristan Brandenburg, Daniel I. Chasman, Ching-Yu Cheng, Marina Ciullo, Maria Pina Concas, David Conen, Simon R. Cox, Luc Dauchet, Hithanadura Janaka de Silva, Marcus Dörr, Todd L. Edwards, Ervin R. Fox, Nora Franceschini, Barry I. Freedman, Giorgia Girotto, Vilmundur Gudnason, Sioban D. Harlow, Iris M. Heid, Adriana M. Hung, Sahoko Ichihara, Cashell E. Jaquish, Catherine John, Jost B. Jonas, J. Wouter Jukema, Norihiro Kato, Bernard D. Keavney, Tanika N. Kelly, Markku Laakso, Paul Lacaze, Timo Lakka, Seunggeun Lee, Terho Lehtimäki, Ching-Ti Liu, Ruth J.F Loos, Kari E. North, Brenda Penninx, Michael A. Province, Bruce M. Psaty, Susan Redline, Frits R. Rosendaal, Charles N. Rotimi, Jerome I. Rotter, Maria Sabater-Lleal, Helena Schmidt, Xueling Sim, Harold Snieder, Beatrice Spedicati, Klaus J. Stark, Chikashi Terao, Lynne E. Wagenknecht, David R. Weir, Wei Zhao, Xiaofeng Zhu, Patricia B. Munroe, Yan V. Sun, James Gauderman, Alisa K. Manning, Myriam Fornage, Hugues Aschard, Heming Wang, Paul S. de Vries, Gao Wang, Dabeeru C. Rao, Alanna C. Morrison, Han Chen

## Abstract

Cigarette smoking influences blood pressure (BP) levels. Studying and accounting for potential gene-smoking interactions can help discover novel loci and provide insights into biological pathways for smoking-associated BP regulation. We conducted a genome-wide association meta-analysis involving 1,188,241 individuals from 66 studies in five ancestry groups, analyzing systolic BP, diastolic BP, and pulse pressure while considering interactions between genetic variants and three smoking exposures: smoking status, cigarettes per day, and pack years. These analyses identified twelve novel loci for BP at genome-wide significance (*P* < 5 × 10^−9^), and highlighted biological processes including tight junction integrity, mitochondrial health, vascular relaxation, and endothelial function. In smoking status-stratified analyses, smoking modifies the genetic effect of six variants on BP. To prioritize likely causal, we developed and applied SuSiEgxe, a fine-mapping method based on a two-degree-of-freedom joint test using gene-environment interaction summary statistics. Fine-mapped loci uncovered immune-related pathway for smoking-associated BP regulation.

## 1 Introduction

The exploration of genetic variants related to blood pressure (BP) regulation has been significantly advanced through genome-wide association studies (GWAS). Gene-by-environment (G×E) interaction studies are critical for understanding how genetic predispositions may amplify or mitigate the effects of lifestyle choices. Among these lifestyle exposures, cigarette smoking is a major risk factor for high BP (hypertension) and has been shown to influence BP regulation through various biological mechanisms. Given that hypertension is a core risk factor for cardiovascular diseases, smoking may contribute to cardiovascular risk directly or through its impact on BP^1, 2, 3^. Investigating gene-by-smoking interactions in BP GWAS can reveal novel loci and enhance understanding of the biological pathways that regulate BP. Particularly, it is crucial to identify BP variants with heterogeneous associations in smokers and nonsmokers or across different levels of smoking exposure, e.g., cigarettes per day or pack years. Such analyses reveal genetic factors that may influence BP differently according to smoking status and helps to refine the understanding of how environmental exposures like smoking interact with genetic variation.

Previous studies have investigated gene-smoking interactions in BP traits, particularly within the Cohorts for Heart and Aging Research in Genomic Epidemiology (CHARGE) consortium. However, they did not include marginal genetic effect-only analyses that excluded smoking, and it was unknown if any of the loci found through the two-degree-of-freedom (2df) joint test could have been discovered without smoking or gene-smoking interaction in the model^4^. This gap makes it difficult to determine whether the loci newly identified through the 2df joint test could have been detected without accounting for smoking or gene-smoking interactions. Moreover, there was no analysis with fine mapping to determine the causal single-nucleotide polymorphisms (SNPs) underlying the observed interactions. Fine mapping aims to narrow down the potential causal variants by analyzing linkage disequilibrium (LD) patterns (CAVIAR^5^, SuSiE^6, 7^, FINEMAP^8^, etc.) and functional annotations (eCAVIAR^9^, PAINTOR^10^, etc.). However, existing fine mapping methods were developed focusing on the marginal genetic effect only. Fine-mapping approaches within the context of gene-environment interactions remain underdeveloped, especially using genomic summary statistics.

In this study, we conducted genome-wide association meta-analyses incorporating gene-smoking interactions to identify genetic loci associated with BP traits and to assess the modifying effect of cigarette smoking on the genetic architecture of BP. We also conducted fine mapping with our developed tool SuSiEgxe that uses summary statistics from the 2df joint test in variant-smoking status (CURSMK) interaction meta-analyses. Our analysis included a large sample size (1,188,241 individuals from five ancestry groups) to maximize statistical power for discovery. To account for potential heterogeneity in interaction effects, we report results stratified by ancestry group, in addition to our primary cross-population meta-analyses.

## 2 Results

The study received approval from the Institutional Review Board of The University of Texas Health Science Center at Houston and was conducted in compliance with all applicable ethical regulations. Ethical approval for data collection was obtained from the relevant review boards of each participating cohort, and informed consent was obtained from all participants.

### 2.1 Overview

The investigation includes meta-analyses of 66 studies, and 1,188,241 samples from five ancestry groups: 804,275 (68%) European ancestry (EUR), 246,825 (21%) East Asian ancestry (EAS), 86,580 (7%) African ancestry (AFR), 37,949 (3%) Hispanic admixed ancestry (HIS), 12,612 (1%) South Asian ancestry (SAS). Sex-stratified meta-analyses were performed to evaluate sex heterogeneity in the associations as well. We focused on systolic blood pressure (SBP), diastolic blood pressure (DBP) and pulse pressure (PP) while considering interactions between genetic variants and three smoking exposures: current smoking status (CURSMK), and within current smokers, cigarettes per day (CPD), and pack years (PY). Meta-analyses of all combinations of smoking exposures and BP traits were conducted within each ancestry and across all five ancestries (Cross-Population Meta-Analysis; CPMA). The descriptive statistics of the studies can be found in Supplementary Table S1. We considered two models for each smoking-BP combination within each population group separately. Model 1 is a joint analysis of the genetic main effect and smoking interaction effect where we considered a one-degree-of-freedom (1df) interaction test for the interaction term and a 2df joint test that considered the main and interaction term. Model 2 only analyzed the marginal genetic effect without accounting for smoking exposures.

With the criteria described in the Methods section, we identified 1,753 independent loci by accounting for interaction effects with CURSMK, CPD or PY from either 1df interaction test or 2df joint test in ancestry specific and cross-population analyses. No loci were identified in SAS. The 1df interaction tests identified two novel BP loci and ten significant known BP loci (Table 1-2). The 2df joint tests identified ten novel BP loci (Table 1) and 106 known BP loci (Supplementary Table S6) that were not identified by marginal tests in Model 2 (see details in Methods section). Among the 12 novel loci in total that reached a stringent genome-wide significance threshold (*P* < 5 × 10^−9^), two were for SBP, six for DBP, and four for PP (Supplementary Figure S1).

**Table 1.**
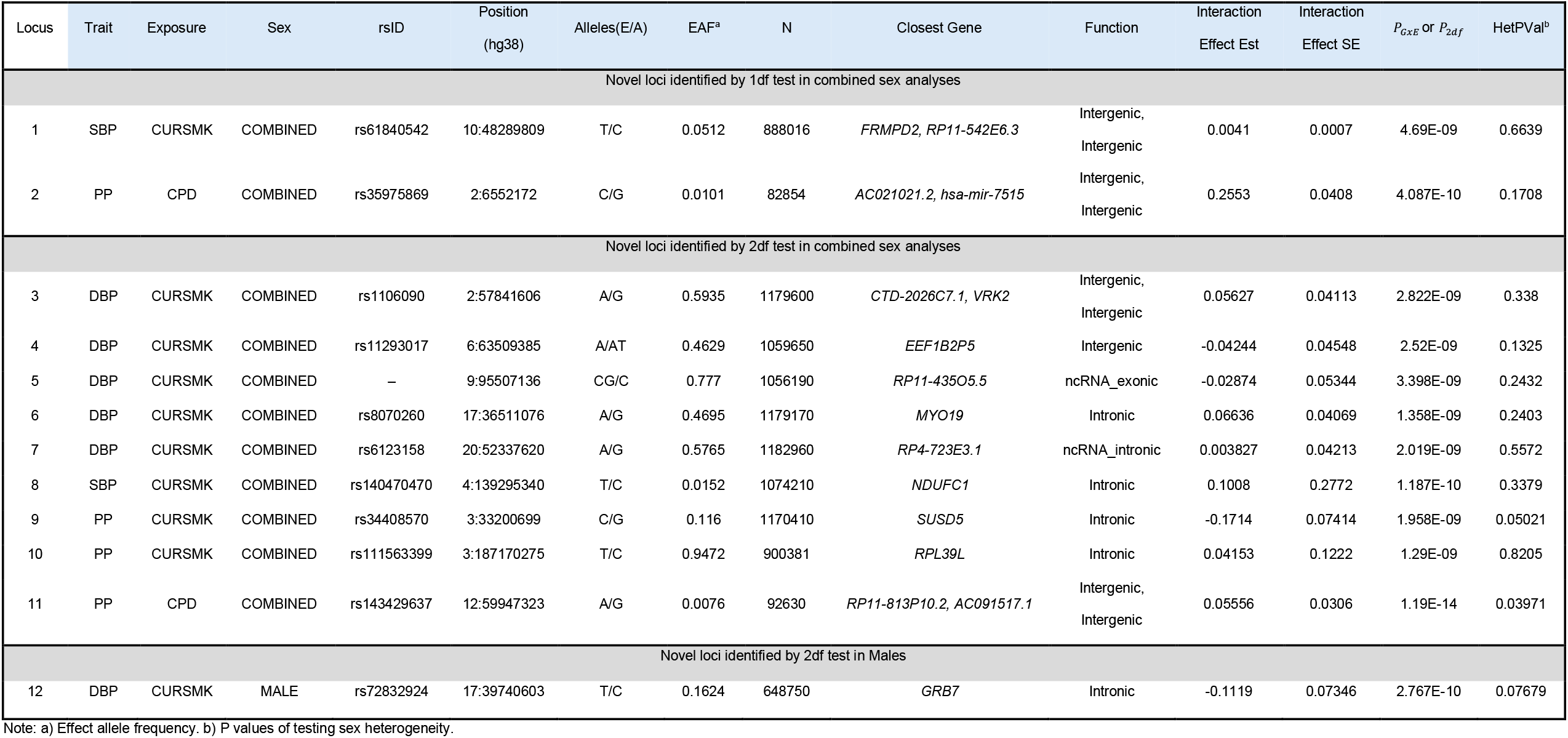
Novel BP Loci Identified in Cross-Population Meta-Analysis.

#### Variant-smoking interaction and joint analyses in CPMA

In the combined sex analyses (Supplementary Figure S2 A), the 1df interaction test revealed four loci showing variant-smoking interaction effects on BP outcomes, including two novel loci: rs35975869 (*AC021021*.*2*; *P*_*GxE*_ = 4.1 × 10^−10^) for the variant-CPD interaction effect on PP, and rs61840542 (*FRMPD2*; *P*_*GxE*_ = 4.7 × 10^−9^) for the variant-CURSMK interaction effect on SBP. There was no evidence of a difference in either of these interactions by sex. In the 2df joint test, 826 loci were identified, and nine were novel (Table 1). In males, the 2df joint test revealed two novel BP loci: rs72832924 (*GRB7*; *P*_2*df*_ = 2.8 × 10^−10^) showed the joint effect of variant-CURSMK interaction on DBP and was only identified in males, and rs143429637 (*RP11-813P10*.*2*; *P*_2*df*_ = 2.1 × 10^−24^) showed the joint effect of variant-CPD interaction on PP. There were no associations for 1df interaction among males or females, and no novel BP loci were identified with the 2df joint test among females (Supplementary Table S4).

#### Variant-smoking interaction and joint analyses in EUR

In the EUR combined sex analyses (Supplementary Figure S2 B), one novel locus with lead variant rs35975869 (*AC021021*.*2*; *P*_*GxE*_ = 1.5 × 10^−9^) was identified by the 1df interaction test in the analysis of variant-CPD interaction on PP. In the 2df joint test, rs143429637 (*RP11-813P10*.*2*; *P*_2*df*_ = 1.1 × 10^−16^) was a novel BP locus revealed by the analysis of variant-CPD interaction on PP. These loci were also identified in CPMA. In males, 292 statistically significant loci were identified in the 2df joint test, and one of them with lead variant rs143429637 (*RP11-813P10*.*2*; *P*_2*df*_ = 3.2 × 10^−27^) was novel. There were no associations for the 1df interaction among males, and no novel BP loci were identified with the 2df joint test among females.

#### Variant-smoking interaction and joint analyses in EAS

There were no novel associations from the 1df interaction or 2df joint test in EAS. In the combined sex analyses (Table 2, Supplementary Figure S2 C), the known BP locus with lead variant rs3782886 (*BRAP*) was identified in variant-CURSMK interaction for both SBP (*P*_*GxE*_ = 7.0 × 10^−24^) and PP (*P*_*GxE*_ = 1.5 × 10^−11^). The known BP locus with lead variant rs116873087 (*NAA25*) was identified in variant-CURSMK interaction on DBP (*P*_*GxE*_ = 7.0 × 10^−19^).

**Table 2.**
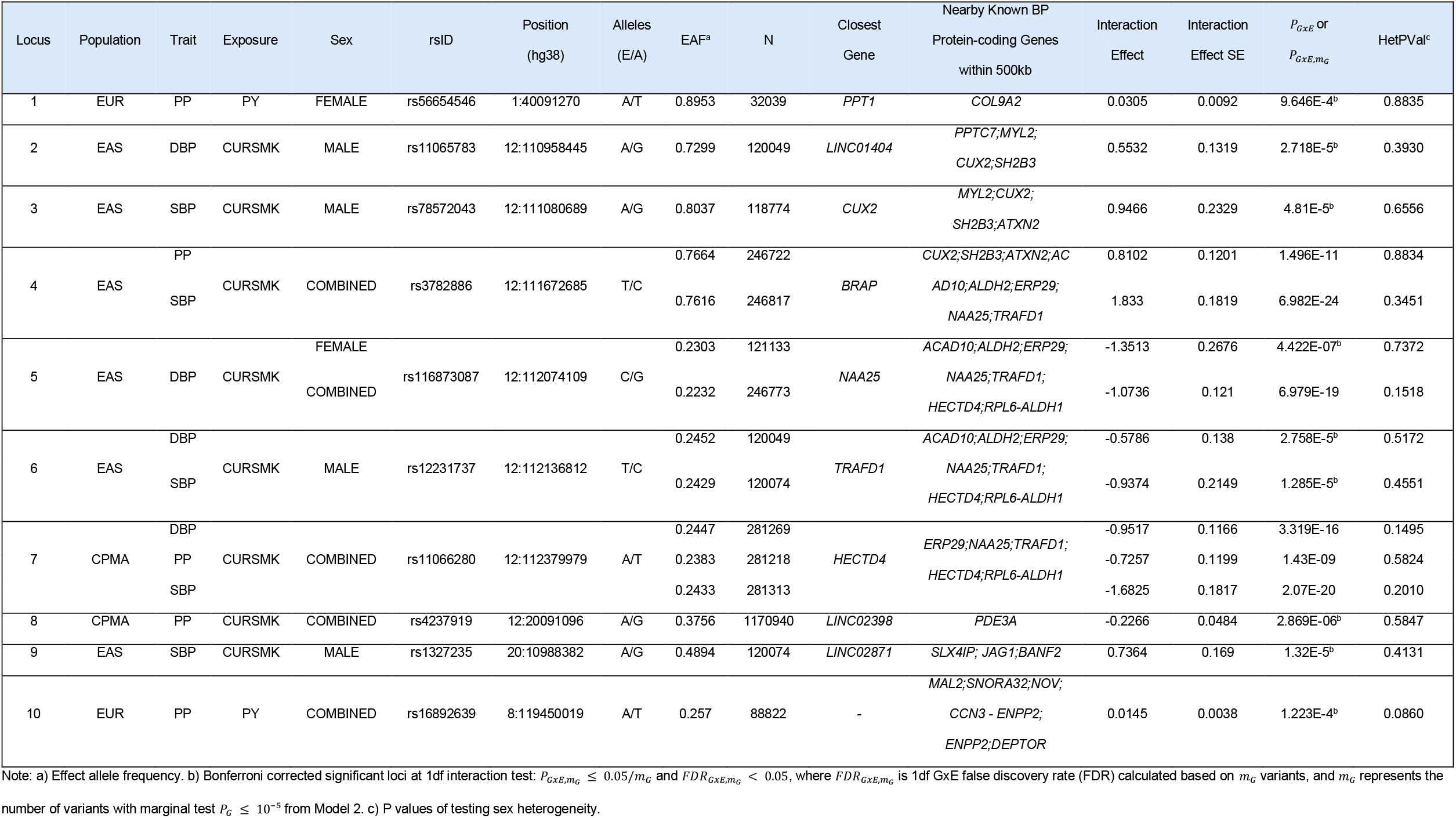
BP Known Loci Identified Using the 1df Interaction Model.

#### Variant-smoking interaction and joint analyses in AFR

There were no novel associations from the 1df interaction or 2df joint test in AFR. The 2df joint test revealed nine significant known BP loci in the combined sex analyses (Supplementary Figure S2 D). Two known BP loci with lead variants rs1716694 (*ULK4*; *P*_2*df*_ = 6.4 × 10^−10^) and rs74884236 (*MFSD1*; *P*_2*df*_ = 4.4 × 10^−9^) were only found in females (Supplementary Table S7).

#### Variant-smoking interaction and joint analyses in HIS

There were no novel associations from the 1df interaction or 2df joint test in HIS. The 2df joint test found two known BP loci in the combined sex analyses (Supplementary Figure S2 E). The variant rs935168 (*KCNK3*; *P*_2*df*_ = 2.1 × 10^−12^) was identified in variant-CURSMK interaction study on DBP, and rs2193950 (*P*_2*df*_ = 2.2 × 10^−9^) was identified in the variant-CURSMK interaction with SBP (Supplementary Table S7).

### 2.2 Smoking status-stratified analyses for significant 1df interaction variants

We further investigated the smoking-status stratified associations for the significant 1df interaction variants with the framework developed by Laville, et al^11^ and applied with tool **j2s** (https://gitlab.pasteur.fr/statistical-genetics/j2s). The results (Table 3) demonstrated differences in the genetic effects on blood pressure between current smokers and non-current smokers. rs3782886-T is a missense variant at *BRAP*, and it has a significant association with higher PP among smokers (β = 1.09, *P* = 6.9 × 10^−22^), but not among non-smokers (β = 0.27, *P* = 3.9 × 10^−8^) in EAS. Similarly, rs116873087-C and rs12231737-T are intronic variants at *NAA25*, and *TRAFD1* respectively. The two loci have significant association with lower DBP among smokers but not among non-smokers in females and males. rs11066280-A is an intronic variant at *HECTD4*, and it has a significant association with lower PP among smokers but not among non-smokers. Additionally, rs4237919-A and rs1327235-A are at a gene for long intergenic non-protein coding RNA and are significantly associated with PP and SBP among non-smokers, respectively.

**Table 3.** Smoking Status-Stratified Summary Statistics at Significant 1df Interaction Loci Known for BP.

| Locus<sup>a</sup> | Population | Trait | Exposure | Sex | rsID | Position<br>(hg38) | Alleles<br>(E/A) | EAF<sup>b</sup> | N | N<br>(E = 1)<sup>a</sup> | Genetic<br>Effect<br>(E = 1) | Genetic<br>Effect SE<br>(E = 1) | P (E = 1) | N<br>(E = 0)<sup>b</sup> | Genetic<br>Effect<br>(E = 0) | Genetic<br>Effect SE<br>(E = 0) | P (E = 0) |
| --- | --- | --- | --- | --- | --- | --- | --- | --- | --- | --- | --- | --- | --- | --- | --- | --- | --- |
| 2 | EAS | DBP | CURSMK | MALE | rs11065783 | 12:110958445 | A/G | 0.7299 | 120049 | 32416 | 1.101 | 0.114 | 3.793E-22 | 87632 | 0.527 | 0.068 | 1.271E-14 |
| 3 | EAS | SBP | CURSMK | MALE | rs78572043 | 12:111080689 | A/G | 0.8037 | 118774 | 32079 | 2.779 | 0.204 | 3.595E-42 | 86694 | 1.771 | 0.120 | 1.570E-49 |
| 4 | EAS | PP | CURSMK | COMBINED | rs3782886 | 12:111672685 | T/C | 0.7664 | 246722 | 40820 | 1.087 | 0.113 | **6.855E-22** | 205901 | 0.268 | 0.049 | **3.917E-08** |
|  |  | SBP |  |  |  |  |  | 0.7616 | 246817 | 40843 | 2.727 | 0.172 | 2.478E-56 | 205973 | 0.886 | 0.075 | 6.575E-32 |
| 5 | EAS | DBP | CURSMK | FEMALE | rs116873087 | 12:112074109 | C/G | 0.2303 | 121133 | 8196 | -1.584 | 0.256 | **6.204E-10** | 112936 | -0.240 | 0.067 | **3.428E-04** |
|  |  |  |  | COMBINED |  |  |  | 0.2232 | 246773 | 40828 | -1.692 | 0.114 | 2.923E-50 | 205944 | -0.623 | 0.049 | 1.477E-36 |
| 6 | EAS | DBP | CURSMK | MALE | rs12231737 | 12:112136812 | T/C | 0.2452 | 120049 | 8196 | -1.462 | 0.249 | **4.186E-09** | 112936 | -0.251 | 0.066 | **1.299E-04** |
|  |  | SBP |  |  |  |  |  | 0.2429 | 120074 | 32430 | -2.889 | 0.186 | 3.749E-54 | 87643 | -1.854 | 0.112 | 7.202E-62 |
| 7 | CPMA | DBP | CURSMK | COMBINED | rs11066280 | 12:112379979 | A/T | 0.2447 | 281269 | 53924 | -1.511 | 0.111 | 3.142E-42 | 227344 | -0.567 | 0.048 | 1.338E-31 |
|  |  | PP |  |  |  |  |  | 0.2383 | 281218 | 53915 | -0.993 | 0.114 | **3.251E-18** | 227302 | -0.257 | 0.049 | **1.669E-07** |
|  |  | SBP |  |  |  |  |  | 0.2433 | 281313 | 53935 | -2.528 | 0.175 | 2.439E-47 | 227377 | -0.837 | 0.076 | 5.785E-28 |
| 8 | CPMA | PP | CURSMK | COMBINED | rs4237919 | 12:20091096 | A/G | 0.3756 | 1170940 | 224496 | -0.093 | 0.046 | **0.042** | 946443 | 0.129 | 0.020 | **2.878E-10** |
| 9 | EAS | SBP | CURSMK | MALE | rs1327235 | 20:10988382 | A/G | 0.4894 | 120074 | 32430 | 0.097 | 0.145 | **0.505** | 87643 | -0.618 | 0.089 | **4.897E-12** |

Note: a) Same locus has the same number same as in Table 2. b) E = 1 is exposed group to smoking. c) E = 0 is unexposed group to smoking

### 2.3 Functional annotations for novel loci

The gene mappings for *LGSN* (rs11293017), *TSHZ2* (rs6123158), *RPL39L* (rs111563399), *GSDMB* (rs72832924), and *ORMDL3* (locus 12) were validated by tissue-specific eQTL evidence from GTExv8, which was observed in whole blood, lung, or subcutaneous adipose tissue (Supplementary Table S12). Tissue-specific (GTExv8) eQTL associations were observed at rs61840542 (Table 1, locus 1) in artery tibial tissues. Additionally, the expression of *STARD* and *GRB7* were associated with the novel locus with lead variant rs72832924 (Table 1, locus 12), which was exclusively identified in males, based on eQTL evidence from GTExv8 cells cultured fibroblasts tissue. Similarly, the gene mappings for *PTCH1* (9:95507136:CG_C), *TSHZ2* (rs6123158), and *GLB1* (rs34408570) were observed in cells cultured fibroblasts tissue based on eQTL GTExv8 evidence.

Many genes were linked to novel loci through chromatin interactions (Supplementary Table S12). These interactions were observed in various tissues and cell types, including the left and right ventricles of the heart, aorta, lung, GM12878 (lymphoblastoid cell line), IMR90 (lung fibroblast cell line), mesenchymal stem cells, and mesendoderm cells.

We performed gene-set enrichment analysis using genes mapped through positional, expression quantitative trait loci (eQTL), and chromatin interaction data (Supplementary Figure S3). The carbohydrate phosphatase activity gene set was associated with the novel locus with lead variant 9:95507136:CG_C (adjusted *P* = 0.036). The 17q12 Copy Number Variation Syndrome gene set was linked to the novel locus with lead variant rs8070260 (*MYO19*; adjusted *P* = 7.8 × 10^−69^). Additionally, GWAS catalog gene sets related to asthma were mapped to the novel locus with lead variant rs72832924, which was identified specifically in males.

### 2.4 New findings from 2df fine mapping

Causal variants were identified by our new 2df fine mapping tool SuSiEgxe (Supplementary Table S13), followed by the gene mapping and functional annotations by FUMA. We investigated one novel locus on gene *MYO19*, and five loci from previously reported genes associated with BP (Supplementary Table S3): *KCNK3, ACAD10, MAEA, CCDC162P, and ASB3* (Supplementary Table S14).

For the known BP locus with lead variant rs12476527 at gene *KCNK3*, which was also reported in our previous work^4^, fine mapping narrowed the region (2:26681997-26710019) from 28 kb to 18.9 kb and identified a 13-variant credible set from 27 significant variants in EAS (Figure 1A), and narrowed the region (2:26673079-26714801) from 41.7 kb to 11.6 kb and identified a nine-variant credible set from 30 significant variants in CPMA (Figure 1B). Causal variants were mapped to genes *KCNK3, NRBP1*, and *CENPA* by eQTL evidence in heart atrial appendage tissue and heart left ventricle tissue. Chromatin interactions were observed in GM12878 (lymphoblastoid cell line), IMR90 (lung fibroblast cell line), mesenchymal stem cells, and mesendoderm cells (Supplementary Table S14). The gene-set enrichment analysis highlighted the pathways of fatty acid β-oxidation, mitochondrial metabolism, and valproic acid metabolism (Supplementary Figure S4 A).

**Figure 1.**
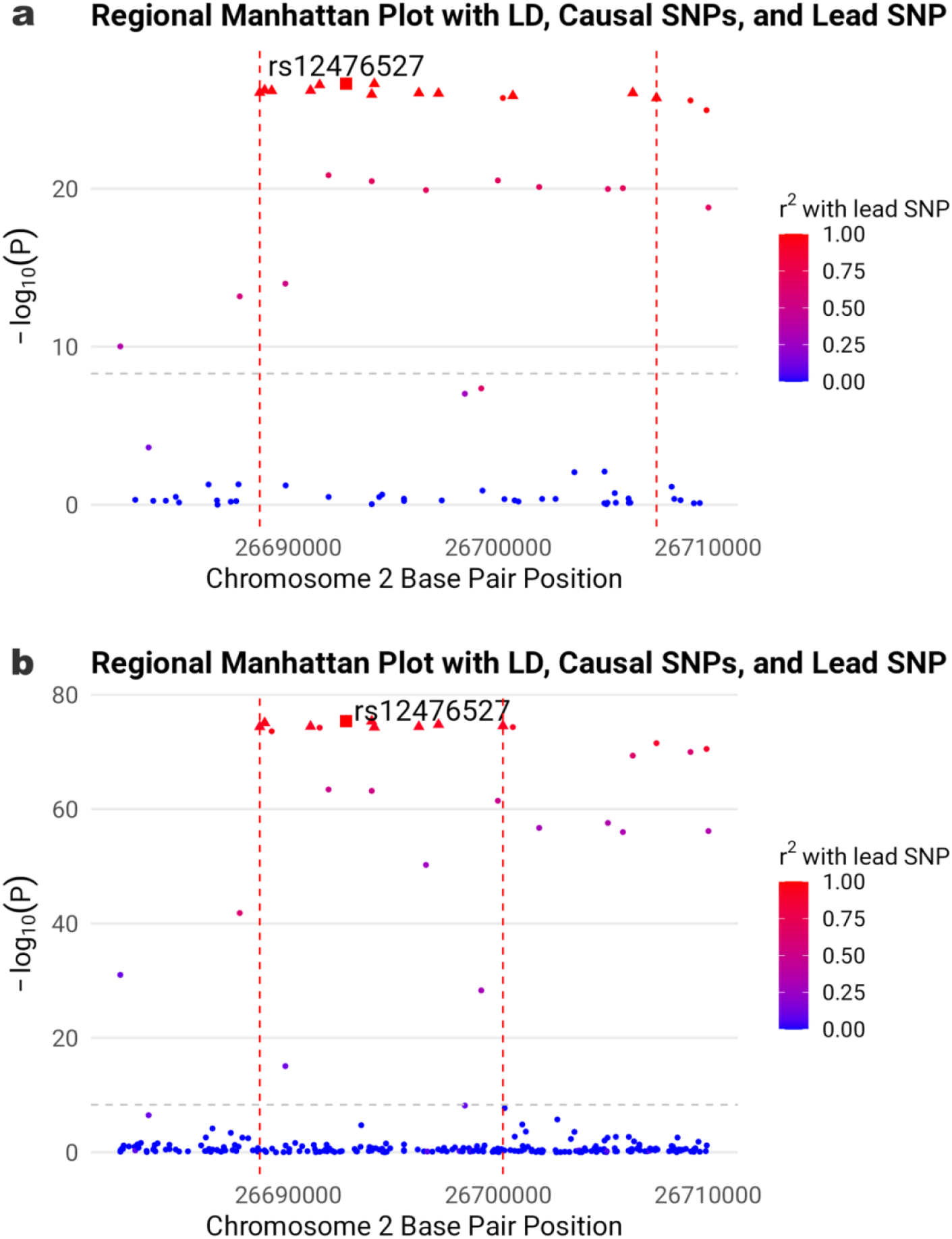
Two-degree-of-freedom fine mapping of locus with lead variant 2:26692756:T_G (rs12476527) on gene *KCNK3*. **a**) East Asian ancestry. **b**) Cross-population meta-analysis. The figure shows causal SNPs (triangles) on gene *KCNK3* for variant-CURSMK interaction on DBP. Lead variant is indicated by squares. The grey dashed horizontal line marks the threshold for genome-wide significance at − log_10_ 5 × 10^−9^. The red dashed vertical lines mark the range of credible sets.

The novel *MYO19* locus with lead variant rs8070260 was identified by the 2df joint test in Model 1 (*P*_2*df*_ = 1.4 × 10^−9^), whereas it was not identified through the marginal test in Model 2 (*P*_1*df*_ = 8.3 × 10^−9^). Fine mapping analysis using the 2df joint test pinpointed three potential causal variants, including rs1109442, rs8070260, and rs7222903 (Supplementary Figure S5 A), which exhibited consistent functional annotations when incorporating all variants within the locus (Supplementary Table S14, Supplementary Figure S4 B).

Similarly, loci with lead variants rs60722337 at *MAEA*, rs6902892 at *CCDC162P*, and rs74598641 at *GPR75-ASB3 / ASB3* (Supplementary Figure S5 B-D) were identified by the 2df joint test in Model 1 but were not detected by the marginal test in Model 2. Causal variants were mapped to genes *MAEA, UVSSA, CRIPAK, CTBP1, CD164, SMPD2, AK9, FIG4, PSME4, GPR75*, and *GPR75-ASB3* by eQTL evidence in tissue whole blood, artery aorta, artery coronary, artery tibial, heart atrial appendage, heart left ventricle, lung, and cells cultured fibroblasts. Chromatin interactions were observed in various tissues and cell types, including the left and right ventricles of the heart, aorta, lung, GM12878 (Lymphoblastoid cell line), IMR90 (lung fibroblast cell line), mesenchymal stem cells, and mesendoderm cells (Supplementary Table S14).

The locus with lead variant rs11066015 at gene *ACAD10* was only identified in the East Asian population (EAS) (Supplementary Figure S5 E). Chromatin interactions between causal variants and genes *ATXN2, BRAP, ACAD10, ALDH2, MAPKAPK5*, and *TRAFD1* were observed in IMR90 (lung fibroblast cell line), mesenchymal stem cells, and mesendoderm cells (Supplementary Table S14). The mapped genes show a highly significant enrichment for traits linked to coronary artery disease, coronary heart disease, hypertension, mean arterial pressure, and alcohol consumption (Supplementary Figure S4 C).

### 2.5 Genome-wide MAGMA analysis and gene-set enrichment

MAGMA gene-based and gene-set tests were applied to the full list of 2df joint p-values of CPMA for all smoking and BP combinations in the combined sex analyses (Supplementary Table S5).

We observed 19 significant protein-coding genes mapped based on 2df joint p-values from multiple analyses (Supplementary Table S8). The gene *ULK4* was mapped in five analyses: DBP-CURSMK (*P* = 1.2 × 10^−17^), DBP-CPD (*P* = 1.8 × 10^−6^), DBP-PY (*P* = 2.3 × 10^−6^), PP-CPD (*P* = 4.8 × 10^−7^), and PP-PY (*P* = 2.8 × 10^−8^). Additionally, the genes *HECTD4, NAA25*, and *TRAFD1* were found based on the tests of variant-smoking interaction with traits SBP and DBP.

MAGMA gene-set analyses (Supplementary Table S9) highlighted several gene sets and biological processes that affect the association between smoking and BP. Significant gene sets were related to the regulation of RNA metabolic processes (DBP-CURSMK, *P* = 1.1 × 10^−15^), calcium channel activity in cardiac muscle cells (DBP-CURSMK, *P* = 4.5 × 10^−15^; SBP-CURSMK, *P* = 4.0 × 10^−7^), muscle structure development (PP-CURSMK, *P* = 5.0 × 10^−14^), muscle tissue development (PP-CURSMK, *P* = 9.9 × 10^−13^), and elastic fiber assembly (PP-CURSMK, *P* = 3.1 × 10^−12^).

Gene set enrichment analyses using Functional Mapping and Annotation of Genome-Wide Association Studies (FUMA)^12^ GENE2FUNC (Supplementary Table S10) highlighted several biological processes that affect the blood, heart, or lungs. Selected significant pathways were the regulation of BP (DBP-CURSMK, adjusted *P* = 0.015; SBP-CURSMK, adjusted *P* = 0.019), tube morphogenesis (PP-CURSMK, adjusted *P* = 0.011), and regulation of tube size (DBP-CURSMK, adjusted *P* = 0.004; SBP-CURSMK, adjusted *P* = 0.033), heart development, regulation of heart contraction, heart valve morphogenesis, the circulatory system processes and development with genes *ACE, AGT, NOS3, NPPA, NPPB*, etc. involved. Notably, the pathways related to type I interferon in the immune system were found in SBP, DBP, and quantitative smoking exposure CPD and PY analyses with genes *PTPN11, OAS1, OAS3*, and *OAS2*.

We assessed whether the positionally mapped genes were overrepresented in traits and diseases previously identified in the GWAS Catalog based on a 2df joint test in CPMA with the GENE2FUNC module in FUMA (Supplementary Table S11). Traits associated with SBP, DBP, PP, hypertension, and mean arterial pressure demonstrate highly significant enrichment across various smoking exposure groups with genes such as *KCNK3, ULK4, BRAP*, etc. Additionally, coronary artery disease and myocardial infarction exhibit significant enrichment across multiple gene sets with genes such as *CYP17A1, CNNM2, ACAD10, ALDH2, NAA25, HECTD4*, etc.

## 3 Discussion

This study presents a comprehensive analysis of gene-smoking interactions on BP across five ancestry groups. A total of 66 studies and 1,188,241 participants were included in the analysis. Including EUR, EAS, AFR, HIS, and SAS allowed for a broad exploration of genetic associations. Our study identified 12 novel loci, associated with BP traits, with a focus on SBP (n = 2 loci), DBP (n = 6 loci), and PP (n = 4 loci).

Newly identified protein-coding genes showed potential pathways for regulating BP through the interaction with smoking. The GxE 1df interaction test identified a novel locus (Table 1) adjacent to *FRMPD2* (rs61840542, *P*_*GxE*_ = 4.7 × 10^−9^), which is a scaffolding protein that maintains epithelial cell polarity and tight junction integrity to support correct cell-cell adhesion and barrier function^13^ and in glutamate-gated ion channels expressed in the central nervous system^14^. The disruption of *FRMPD2*-mediated signaling may lead to increased permeability in the lungs and blood vessels, and then promotes inflammation and disease progression through impaired tight junctions and epithelial integrity triggered by smoking^15^.

The GxE 1df interaction test identified 10 BP loci (Table 2) that were reported to have marginal effects in previous studies and highlighted their effect on BP with interaction of smoking. The locus with lead variant rs56654546 at *PPT1* was identified in EUR females. The locus with lead variant rs78572043 at *CUX2* was identified in EAS males. Loci with lead variants rs3782886 at *BRAP*^16^, rs116873087 at *NAA25*^17, 18^, *and rs12231737 at TRAFD1*^18^ were only identified in EAS. A locus with lead variant rs11066280 at *HECTD4*^19, 20^ *was identified in CPMA. These loci are located near the protein-coding genes known to be related to BP, including COL9A2*^21^, *PPTC7*^18^, *MYL2*^*19*^, *SH2B3*^22^, *ATXN2*^*17*, *23*^, *ACAD10*^*17*^, *ALDH2*^*20*^, *ERP29*^*24*^, *RPL6-ALDH1*^*20*^, *PDE3A*^25^, *SLX4IP*^25^, *JAG1*^*25*, *26*, *27*^, *BANF2*^25^, *MAL2*^*24*^, *SNORA32*^*28*^, *MIR548AZ - CCN3*^*17*^, *NOV*^*26*^, *CCN3 - ENPP2*^*17*^, *ENPP2*^18^. *CUX2* was associated with DBP and SBP through N^6^-methyladenosine (m^6^A)-associated SNP rs7398833 in East Asian population^29^. In a study of elderly Japanese population that accounted for smoking status, the minor allele of rs3782886 at *BRAP* was inversely associated with hypertension only in participants with high hematopoietic activity^30^. *NAA25*, in a region with *ERP29*^31^, was previously reported to be associated with SBP and DBP^32^.

The smoking status-stratified analyses further illustrate how smoking modifies the genetic effect on BP. One significant 1df variant rs3782886-T at *BRAP* is associated with increased BP among smokers, while three loci at *NAA25, TRAFD1* and *HECTD4* were associated with decreased BP among smokers. *TRAFD1* is a protein-coding gene that is primarily related to immune and inflammatory responses via its interactions with NF-κB and TNF receptor signaling pathways^33, 34^. Cigarette smoking activates the NF-kB pathway, which up-regulates various pro-inflammatory genes, leading to increased levels of inflammatory markers and cytokines, and contributing to its pro-inflammatory effects on the immune system^35^, potentially increasing the susceptibility to hypertension and cardiovascular diseases through inflammatory mechanisms and vascular dysfunction. *HECTD4* is a protein-coding gene involved in protein breakdown and recycling. It helps regulate the process of tagging proteins for degradation through the ubiquitin system. Shencun Fang, et al^36^. identified a potential role of the circHECTD1/HECTD1 pathway in regulating endothelial-mesenchymal transition (EndMT), where both endothelial and epithelial cells contribute to fibroblast accumulation, ultimately promoting the development of pulmonary fibrosis in silicosis, and cardiovascular diseases.

The GxE 2df joint model newly identified protein-coding genes *MYO19* (rs8070260, *P*_2*df*_ = 1.4 × 10^−9^), *VRK2* (rs1106090, *P*_2*df*_ = 2.8 × 10^−9^), *NDUFC1* (rs140470470, *P*_2*df*_ = 1.2 × 10^−10^), *RPL39L* (rs111563399, *P*_2*df*_ = 1.3 × 10^−9^), and *GRB7* (rs72832924, *P*_2*df*_ = 2.8 × 10^−10^ only in males). They showed potential pathways for regulating BP through the joint effect of genetic main effect and interaction with smoking (Table 1). A novel locus with the lead variant rs13032423 mapped to *VRK2*, was previously reported to be associated with BP through an interaction with long total sleep time^37^. Along with the novel locus with lead variant rs1106090 discovered in this study, these findings highlight *VRK2* as potentially relevant to cardiovascular health. *GRB7* has been previously associated with asthma, another trait influenced by smoking exposure^38, 39^. *MYO19* functions as a protein that controls both the movement and operations of mitochondria inside cells. Maintaining mitochondrial health is crucial for essential energy production and cellular stress response mechanisms. Smoking can cause oxidative stress alongside inflammation with the potential to harm mitochondrial structures. Since *MYO19* plays a role in mitochondrial dynamics, disruptions in its function could worsen the effects of smoking and hypertension on cellular health, particularly in the cardiovascular system. This pathway has been verified in mouse experiments by Sergey Dikalov, et al^40^. *NDUFC1* is a subunit of mitochondrial complex I, which plays a key role in the electron transport chain and ATP production. ATP is necessary for maintaining various cellular functions, including the regulation of endothelial nitric oxide synthase (eNOS), which is responsible for nitric oxide (NO) synthesis, a key player in vascular relaxation and endothelial function^41^. Dysfunction in *NDUFC1* may reduce the efficiency of the electron transport chain, leading to decreased ATP production, which can exacerbate endothelial dysfunction and smooth muscle cell impairment in blood vessels, contributing to the progression of high BP. *RPL39L* also influences cell proliferation by regulating protein synthesis and mitochondrial function^42^. Furthermore, we have discovered one molecular function involving the novel locus with lead variant 9:95507136:CG_C that is related to carbohydrate phosphatase activity (Supplementary Figure S3 A). Enzyme activities involve catalyzing the removal of phosphate groups from carbohydrates or carbohydrate derivatives. When mitochondrial structures are damaged due to smoking-induced oxidative stress, it may disrupt the normal activity of these enzymes, which results in altered metabolic processes and contributes to the pathogenesis of smoking-related diseases, including metabolic disorders, and eventually leads to elevation in BP.

There were 106 loci identified through GxE 2df joint model that were not significant in the marginal genetic tests without accounting for smoking, which supports they were driven by gene by smoking interactions (Supplementary Table S6). Some loci are at genes *PRDM16*^*17*, *43*^ (rs111063488), *NOS3*^28, 43, *44*^ *(rs117564322), CREB3L1*^45^ (11:46323702:G_GA), *GSDME*^*17*^ (rs2521769), *NPR1*^28, 43^ *(rs35479618), PRKD3*^*28*, *46*^ (rs59916529), and *YEATS2*^47^ (rs262969). *PRDM16* deficiency is associated with an increased risk of cardiomyopathy and cardiac mortality^48^, and its deficiency in vascular smooth muscle cells leads to disrupted BP circadian variation^49^. *NOS3* (*E298D* and *T-786C* polymorphisms) was previously reported to be associated with the risk of incident coronary heart disease with the interaction of cigarette smoking^50^. Recently, a study showed a positive impact of *NOS3* on angiogenesis through enhanced endothelial cells derived from induced pluripotent stem cells (iPSC-ECs)^51^. Additionally, loss of *CREB3L1* expression leads to upregulation of *FGFbp1* and pleiotrophin, both of which positively regulate angiogenesis^52^. Therefore, *NOS3* and *CREB3L1* are potentially associated with hypertension through the pathway of angiogenesis regulation^53, 54^. *GSDME*-mediated pyroptosis is associated with inflammation of lung^55^ and atherosclerosis^56^, and there is a strong link between atherosclerosis and hypertension^57^. *NPR1* in vascular endothelial cells is the receptor for natriuretic peptides and thus an important regulator of BP via salt/water balance^58^; *NPR1* deficiency also leads to augmented expression of pro-inflammatory cytokines^59^.

Several genes were discovered through the MAGMA and gene mapping and have been associated with BP (Supplementary Table S8-S11). *ULK4*, one of the genes found in five analyses for DBP and PP and other GWAS, encodes a member of the unc-51-like serine/threonine kinase (STK) family and plays a role in neuronal growth and endocytosis. It was reported in many GWAS for BP and hypertension^22, 60, 61, 62^. *HECTD4* and *TRAFD1* were also found in multiple MAGMA gene-based analyses. We also found several genes from novel loci tied to fibroblasts based on eQTL GTEx/v8 evidence, such as *PTCH1, TSHZ2, GLB1, STARD3*, and *GRB7* (Supplementary Table S12).

In this study, we developed and applied fine mapping to the 2df joint test, allowing us to pinpoint the set of potential causal SNPs, particularly for loci that were not identified in the 1df marginal tests at a significance threshold of *P* < 5 × 10^−9^. For instance, the novel locus with lead variant rs8070260 in *MYO19* was identified through the joint analysis of the genetic main effect and the variant-CURSMK interaction effect (*P*_2*df*_ = 1.4 × 10^−9^) but was not detected in the marginal genetic effect analysis (*P*_1*df*_ = 8.3 × 10^−9^). This underscores the value of fine mapping based on the 2df joint test, which leverages its higher statistical power after accounting for heterogeneous genetic effects by smoking exposures. Our 2df fine mapping narrowed down 116 (89.9%) association regions out of 129 selected loci (Supplementary Table S13). More importantly, our 2df fine mapping method showed multi-ancestry fine mapping resulted in narrower credible sets, likely due to the increased stringency of LD when combining data across populations. Out of 29 loci on which we performed both CPMA and single ancestry fine mapping, 19 (65.5%) loci showed a narrower credible set in the multi-ancestry fine mapping.

Based on the causal variants in 95% credible sets, we found more potential pathways or related genes in the regulation of BP and the interaction with smoking. A novel locus with lead variant rs8070260 in *MYO19* was mapped to genes based on genomic position and chromatin interaction, and the gene-set enrichment analysis indicates the mapped genes were related to 17q12 copy number variation (CNV) syndrome (Supplementary Figure S4 A). 17q12 CNV syndrome has been linked to diabetes and kidney disease, both of which are related to hypertension and cardiovascular disease^63, 64, 65^. The genetic deletions or duplications in this region may affect pathways involved in vascular function, potentially increasing susceptibility to high BP and heart disease.

We investigated three more loci that were only discovered in the 2df joint test and not significant in the marginal genetic association test (M2). They were located at rs60722337 (*MAEA*), rs6902892 (*CCDC162P*), and rs74598641 (*GPR75-ASB3* / *ASB3*). The causal variants were mapped to genes *RNF212, GPR75*, etc. *MAEA* gene functions in transcriptional regulation while impacting the cell cycle and apoptosis^66, 67^ and may play a role in cancer development and immune system functioning. *CCDC162P* was reported to be associated with human red blood cell traits^68^. *ASB3* is involved in immune regulation, cell signaling, and the degradation of specific proteins, mainly through its interaction with the ubiquitin-proteasome system^69, 70^.

Our study benefits from a large sample size and multiple population groups, allowing us to identify both population-specific and shared genetic effects, and the LD structure in multi-ancestry populations provides narrower credible sets in fine mapping compared to population-specific fine mapping. Our study utilizes advanced statistical methods, including 2df fine mapping, to improve the resolution of genetic signals. Additionally, functional annotation and enrichment analyses provide mechanistic insights into the biological pathways underlying genetic influences on BP traits with smoking interactions.

However, several limitations must be acknowledged. First, despite the large sample size, the statistical power to detect genetic effects remains limited in ancestry groups with a smaller number of participants such as South Asians and Hispanic admixed populations. Second, while we aimed to control for potential confounders such as population stratification, residual confounding may still exist, particularly for complex traits influenced by both genetic and environmental factors. Third, the quality of fine mapping in meta-analysis highly depends on the quality of LD in the reference panel used. If LD information is sourced from a reference panel that does not match the population group, this could lead to misleading results on the causal variants.

In conclusion, the identified novel loci and functional insights from enrichment analyses and fine mapping highlight multiple biological pathways involved in BP regulation, modulated by the effects of smoking. These pathways include those associated with cardiovascular complications, pulmonary function through fibroblast cells, energy metabolism (specifically mitochondrial and fatty acid metabolism), the immune system, and the broader circulatory system. Our findings underscore the significant role of genetic variation in BP regulation, particularly in the context of smoking interaction.

## 4 Methods

### 4.1 Participating studies

Participants over 18 years old from all studies were stratified into five broad ancestry groups: European ancestry (EUR), African ancestry (AFR), Hispanic admixed (HIS), East Asian ancestry (EAS) and South Asian ancestry (SAS), and analyzed by sex groups (male, female, and combined). All studies followed a centralized analysis protocol for investigating the genome-wide gene-smoking interaction effect of BP. There were five types of studies: cross-sectional studies of unrelated subjects, cross-sectional family studies of related subjects, longitudinal studies of unrelated subjects, longitudinal family studies of related subjects, and case-control studies. All contributing studies received approval from their respective institutional review boards, and all participants provided written informed consent under the Declaration of Helsinki. Further details about the participating cohorts are provided in the Supplementary Note.

### 4.2 Phenotypes and Lifestyle Variables

Resting/sitting SBP (mmHg), DBP (mmHg) and PP are considered as BP outcomes. Averages of multiple readings of SBP and DBP were used in the analysis. SBP and DBP were further adjusted for subjects taking any anti-hypertensive medications by adding 15 mmHg to SBP and 10 mmHg to DBP^71^. PP values were then derived by subtracting DBP from SBP. Extreme values, more than 6 standard deviations (SD) above or below the mean, were winsorized respectively for the three BP outcomes. We considered three smoking variables, CURSMK, CPD and PY. CURSMK is a dichotomous variable and coded as 1 if a participant is a current cigarette smoker and as 0 if a participant is not a current cigarette smoker. CPD was only derived in current smokers. PY was calculated using the following formula: PY = (CPD / 20) * years of smoking. Current smokers were excluded from CPD and PY analyses if they had a value of 0 or missing for that variable. Both CPD and PY were winsorized separately in males and females after exclusion. If the number of current smokers in any group (for instance, a population group or sex stratum) was less than 100 after exclusions and winsorization, only the CURSMK analysis was conducted in that group.

### 4.3 Genotype Data

Genotypes were imputed using reference panels from the Trans-Omics for Precision Medicine (TOPMed) Imputation Server, Haplotype Reference Consortium (HRC) or 1000 Genomes Project (1000G). SNPs with a minor allele frequency (MAF) less than 0.001 in the total sample (sex-combined group), low imputation quality (*r*^2^ < 0.3 if using MACH or *INFO* < 0.3 if using IMPUTE2) and any SNPs mapping to sex chromosomes or mitochondria were excluded (Supplementary Table S2).

### 4.4 Study-specific GWAS

We considered two models for each smoking-BP combination within each population-sex group separately. Model 1 is a joint analysis of the main and interaction effects.

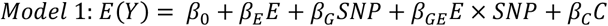

where Y is the BP phenotype; *E* is one of the smoking variables CURSMK, CPD, or PY; SNP is the dosage of the genetic variant; and *C* is a set of covariates including age, age^2^, sex when analyzing males and females combined, field center (as appropriate), genetic principal components (PCs), study-specific confounders, and their interaction with smoking variables *E*, including age x E, age^2^ x *E*, and sex x *E* when analyzing males and females combined. Under the null hypothesis of β_*GE*_ = 0, we assessed the interaction effect between the smoking variable E and genetic variants with a 1df test. Under the null hypothesis of β_*G*_ = β_*GE*_ = 0, a 2df joint test was conducted to assess the joint effect of genetic main effect and interaction effect. Model 2 only analyzes the marginal genetic effect under the null hypothesis of β_*G*_ = 0, and the same covariates are included.

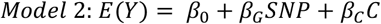

Subjects with no GWAS data or missing data for any of the common covariates —including age, sex, and PCs—were excluded from the analysis.

For cross-sectional studies with unrelated subjects, model-based p-values from linear model were reported. Tools such as **LinGxEScanR** v1.0 (https://github.com/USCbiostats/LinGxEScanR), **MMAP** (https://github.com/MMAP/MMAP.github.io), **GEM v1.4.3** (https://github.com/large-scale-gxe-methods/GEM), and **EPACTS** (https://github.com/statgen/EPACTS) were utilized. A two-step approach was implemented for cross-sectional studies with related subjects: 1) linear mixed models using a kinship matrix to account for family relationships were generated using **MMAP**; and 2) residuals from the linear mixed model were subsequently analyzed as the phenotype using **GEM v1.4.3**. For longitudinal studies, a single visit for each population group was chosen if it has contemporaneous measures of BP, use of anti-hypertensive medications, and smoking, to maximize the sample size and methods used as stated above. For case-control studies, all analyses were performed within case and control samples separately.

### 4.5 Quality Control

EasyQC2^72^ (www.genepi-regensburg.de/easyqc2) was applied to perform quality control (QC) on study-level and meta-level summary statistics within each population group, sex group, and exposure type. In study-specific QC, hg19 build genomic coordinates were lifted over to hg38 genomic coordinates. Variants were excluded if they were monomorphic, had an INFO score (a measure of imputation quality) below 0.6, had an effect allele frequency outside the range of 0.01 to 0.99 with an INFO score below 0.8, or if the minor allele count for either the unexposed group (E=0) or the exposed group (E=1) was below 10. To implement a t-distribution based approach, the effective sample sizes of independent observations in the exposed group or the unexposed group *N*_*E*_ estimated in each study and the MAF of each genetic variant were used to compute approximate degrees of freedom (*DF*)^73^.

### 4.6 Meta-analysis

For the 1df interaction test in Model 1 and marginal genetic effect in Model 2, inverse-variance weighted fixed-effects meta-analysis was performed with METAL^74^ software. For the 2df joint test in Model 1, inverse-covariance-matrix weighted joint fixed-effects meta-analysis of the genetic main effect and the variant by environment interaction was performed in METAL following Manning et al^75^. All meta-analyses were conducted separately for males, females, and the combined group. They were initially performed within each population group (EUR, AFR, SAS, EAS, HIS) and subsequently across populations, with genomic control correction applied to both the population-specific meta-analyses and CPMA. The analyses excluded variants within 1 Mb of the major histocompatibility complex (MHC) region. Additionally, we removed variants with only one population contributed in CPMA and considered variants with at least 20,000 subjects in all analyses. Additional criteria were applied in population-specific meta-analyses, requiring *DF*_*ALL*_ ≥ 20, *DF*_*E*0_ ≥ 10, *DF*_*E*1_ ≥ 10, *INFO* ≥ 0.5, *N* ≥ 100 for each cohort/study, where *DF* is degrees of freedom approximated by *N*_*E*_ and MAF.

### 4.7 Identification of genome-wide independent loci

We used the EasyStrata2^76^ package in R to identify and prioritize loci from significant findings obtained through the 1df interaction test and 2df joint test.

A variant is considered significant if it satisfies the following criteria: (1) 1df interaction test *P*_*GxE*_ ≤ 5 × 10^−9^ and *FDR*_*GxE*_ < 0.05 or 1df interaction test 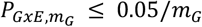 and 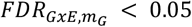, (2) 2df joint test *P*_2*df*_ ≤ 5 × 10^−9^ and *FDR*_2*df*_ < 0.05. *FDR*_*GxE*_ is 1df GxE false discovery rate (FDR) calculated based on all variants genome-wide. 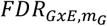is 1df GxE FDR calculated based on *m*_*G*_ variants, and *m*_*G*_ represents the number of variants with marginal test *P*_*G*_ ≤ 10−^5^ from Model 2. 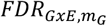is 1df GxE FDR calculated based on *m*_*G*_ variants. *FDR*_2*df*_is 2df joint FDR calculated based on all variants genome-wide. Significant variants and nearby variants within 500 kb regions were grouped together, and independent loci were identified by LD threshold *r*^2^ < 0.1 using TOPMed-imputed 1000G reference panels. Loci were classified as novel or known by assessing their overlap with regions within 1 Mb of variants reported in previous GWAS studies for BP (see Supplementary Table S3 for known BP loci).

Heterogeneity across sexes was assessed using two-sample Z-tests under the null hypothesis of homogeneity across sexes. For each top interaction locus, two-sample Z-tests^77^ were conducted to evaluate sex-specific effect size differences. A significance threshold of *P* = 1 × 10^−6^ was applied to these heterogeneity tests.

### 4.8 Smoking status-stratified analyses

For the variants identified as significant in the 1df interaction test according to the criteria described above, we conducted smoking status-stratified analyses using the tool j2s^11^ (https://gitlab.pasteur.fr/statistical-genetics/j2s). Summary statistics for each variant were generated separately in smokers and non-smokers. These stratified results were used to illustrate whether smoking status modified the genetic associations with BP.

### 4.9 Gene-based analyses and functional annotations

Protein-coding genes were prioritized, and additional annotations for variant mapping were obtained using FUMA v1.5.2.

MAGMA (v1.08)^78^ gene-based and gene-set analyses in FUMA were conducted using the full spectrum of SNP p-values from 2df joint test. SNPs are mapped to 19,232 protein-coding genes with 10 kb window size around the genes. Genome-wide significance was defined as *P* < 2.6 × 10^−6^ based on Bonferroni correction. Gene sets were obtained from MsigDB v7.0, and 15,496 total gene sets were tested. Significant gene set was defined as *P* < 3.23 × 10^−6^ based on Bonferroni correction. To further prioritize genes, the FUMA’s SNP2GENE pipeline was applied to identify candidate genes based on positional mapping. Finally, pathway enrichment analysis of the prioritized genes was performed using the FUMA’s GENE2FUNC pipeline to identify enriched biological pathways, and GWAS catalog reported genes.

For novel loci, the FUMA’s SNP2GENE pipeline was applied to identify candidate genes based on positional mapping (mapped to genes within 10 kb), chromatin interaction mapping (*FDR* ≤ 1 × 10^−6^, 250 bp upstream - 500 bp downstream of transcription start site), and eQTL dataset GTEx v8. Then, gene-set enrichment analysis was performed on mapped genes using FUMA’s GENE2FUNC.

### 4.10 Fine Mapping

We developed SuSiEgxe, a fine mapping approach that uses a 2df joint test in variant-CURSMK interaction meta-analyses using summary statistics. We applied SuSiEgxe to our meta-analysis results of SBP, DBP, and PP, accounting for genetic effect modification by smoking status (current vs. non-smoking). Fine mapping in the framework of the Sum of Single Effects (SuSiE) regression model^6, 7^ was conducted in EAS, EUR, and a CPMA of five super-populations combined. Identified causal SNPs in 95% coverage credible sets were mapped to genes based on their genomic position, eQTL, and 3D chromatin interactions, followed by FUMA’s GENE2FUNC gene-set enrichment analysis. More details about SuSiEgxe can be found in the Appendix.

## Data Availability

All data produced in the present study are available upon publication.

## Data Availability

The summary statistics will be made available upon publication.

## Code availability

All computation central to the conclusions of this meta-analysis was conducted with the following open-source software: LinGxEScanR (https://github.com/USCbiostats/LinGxEScanR), GEM v1.4.3 (https://github.com/large-scale-gxe-methods/GEM), MMAP (https://mmap.github.io/), EPACTS (https://github.com/statgen/EPACTS), EasyQC2 and EasyStrata2 (www.genepi-regensburg.de/charge-gli), METAL (https://csg.sph.umich.edu/abecasis/metal/download/, https://genome.sph.umich.edu/wiki/Meta_Analysis_of_SNPxEnvironment_Interaction), j2s (https://gitlab.pasteur.fr/statistical-genetics/j2s), and SuSiEgxe (https://github.com/MengyuZhang1307/SuSiEgxe). Partial plots were generated by R package qqman (https://github.com/stephenturner/qqman).

## Acknowledgment

This study was largely supported by grants from the U.S. National Heart, Lung, and Blood Institute (NHLBI), the National Institutes of Health, R01HL118305, R01HL145025, and R01HL156991. The content is solely the responsibility of the authors and does not necessarily represent the official views of the National Institutes of Health. Study-specific acknowledgements are provided in the Supplementary Note.

## Author Contributions

M.Z. and M.R.B. conducted centralized project data analyses, data consolidation, meta-analyses, quality control, bioinformatics analysis, and contextual interpretation. M.Z., M.R.B., A.R.B., T.W.W., R.N., P.N., H.W., P.S.de V., G.W., D.C.R., A.C.M and H.C. were part of the writing group and participated in project workflow design, interpretation of results, and drafting the manuscript. A.N.D., L.d.l.F., C.C.G., and D.C.R., participated in centralized study coordination. T.W.W., J.R.O., P.B.M., J.G., A.K.M., H.A., H.C. acted as collaborators facilitating project design, specific code scripts, or specialized analyses. All other co-authors participated in final result interpretation, cohort-level study concept and design, cohort-level phenotype data acquisition and/or quality control, cohort-level genotype data acquisition and/or quality control, and/or cohort-level data-analysis and interpretation. All authors approved the final version of the paper that was submitted to the journal.

## Conflict of Interest/Disclosures

Bruce Psaty serves on the Steering Committee of the Yale Open Data Access Project funded by Johnson & Johnson. Catherine John has a funded research collaboration with Orion for collaborative research projects outside the submitted work. Maria Sabater-Lleal is supported by a Miguel Servet contract from the ISCIII Spanish Health Institute (CPII22/00007) and co-financed by the European Social Fund. HJG has received travel grants and speaker’s honoraria from Indorsia, Neuraxpharm, Servier and Janssen. Susan Redline received personal fees from Eli Lilly and is an unpaid consultant to Apnimed. Han Chen receives consulting fees from Character Biosciences. The remaining authors declare no competing interests.

## Appendix

SuSiE model with gene-environment interaction terms (SuSiEgxe)

Consider a SuSiE model with gene by environment interaction term:

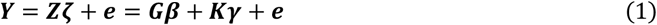

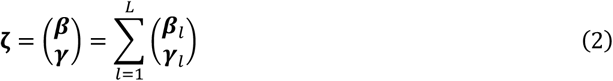

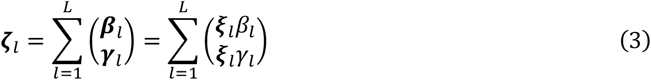

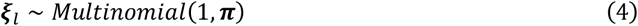

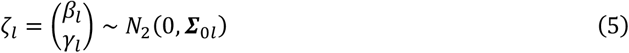

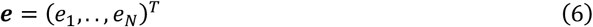

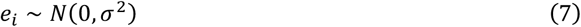

**Y** is an *N* × 1 vector of phenotypes. 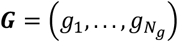 is genotypes of *N*_*g*_ SNPs for *n* subjects, and 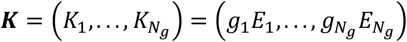 is variant-environment interactions of *N*_*g*_ SNPs for *N* subjects. β_1_, …, β_*L*_, **γ**_1_, …, **γ**_*L*_ are 2*L* single-effect vectors that have only one non-zero entry (equal to β_*l*_ and γ_*l*_ respectively), and their summation is the overall effect vector **β** and **γ**. (β_*l*_, γ_*l*_) follows a bivariate normal distribution with 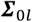 as the prior covariance matrix. **ξ**_*l*_ ∈ {0,1}^*p*^ is an indicator vector following multinomial distribution which has only one non-zero entry. **π** = (π_1_, …, π_*p*_) is the prior inclusion probabilities of each SNP in ***G***. *e*_*i*_ is an independent error terms with hyperparameter σ^2^ as its variance. Assume columns of ***G*** is centered, and ***G*** and binary ***E*** are independent.

When L = 1, it becomes single-effect regression (SER) model:

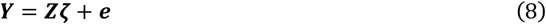

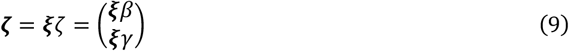

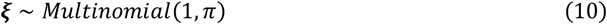

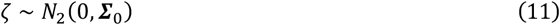

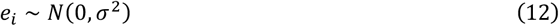

### IBSS-ss algorithms

The estimation of **ζ**_*l*_ in equation (3) given **ζ**_1_, …, **ζ**_*L*−1_ can be obtained by iteratively fitting a SER model with Iterative Bayesian stepwise selection using sufficient or summary statistics (IBSS-ss) (Zou et al. 2022). IBSS computes a variational approximation to the posterior distribution described below.

Posterior distribution and credible sets computation under SER model

Considering SER model equations (9–12), the posterior distribution of **ζ** = **ξ**ζ is

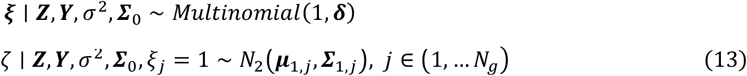

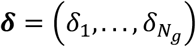 is the vector of posterior inclusion probability (PIPs), where δ_*j*_: = *Pr*(ξ_*j*_ = 1 ∣ ***Z, Y***, σ^2^, **Σ**_0_). By performing *N*_*g*_ Bayesian simple linear regressions, the posterior mean is

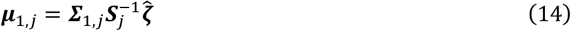

where **Σ**_1,*j*_ is posterior covariance, ***S***_*j*_ is standard errors of estimates of joint effect 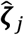from summary data, and

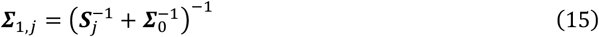

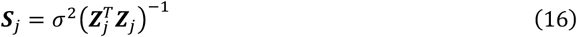

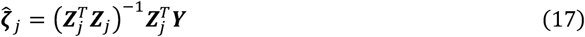

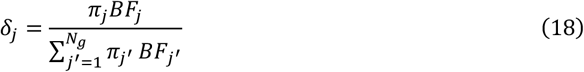

and the Bayes Factor (BF) for comparing this model to null model (ζ = 0) is

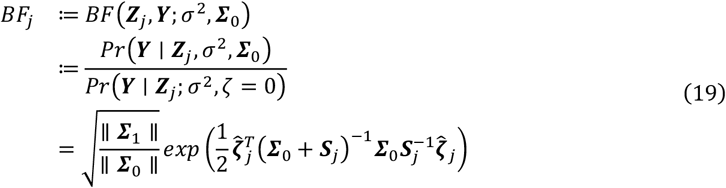

Equations above show that the posterior distribution of **ζ** can be written from ***Z***^*T*^***Z*** and ***Z***^*T*^***Y***. I denote the function that returns it as *SER* − *ss*(***Z***^*T*^***Z, Z***^*T*^***Y***; σ^2^, **Σ**_0_): = (**δ, μ**_1_, **Σ**_1_) under SER model.

A credible set is the set of variants with top PIPs. Variants are ranked in descending order and then added sequentially, starting from the highest PIP, until the cumulative PIP reaches the predefined 95% threshold. This set contains the most likely variants contributing to the trait with a specified probability of interest.

Sufficient statistics and approximations from GWAS meta-analysis summary data Assume that the sample size *N* is large and effect sizes are small. Considering a SER model for ξ_*j*_ = 1, meaning only variant *j* has effect on *Y*, the score test statistics of 2df joint test is

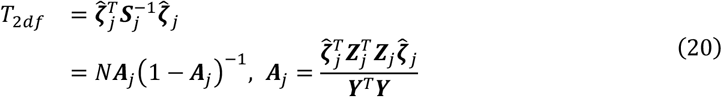

Therefore, the residual variance is

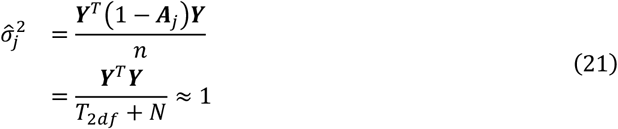

and

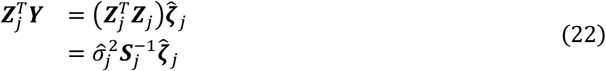

***Z***^*T*^***Z*** can be approximated using LD matrix ***R*** from the reference panel. Besides the correlation between variants ***g***_*i*_ and ***g***_*j*_, the correlation between variants ***g***_*i*_ and variant-environment interaction ***K***_*j*_, correlation between ***K***_*i*_ and ***K***_*j*_ are considered as well. Since *E* is binary, 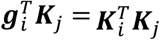. From equation (16), we know

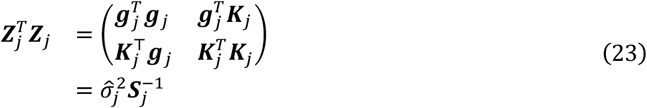

Assuming ***g*** and ***E*** are independent, we have

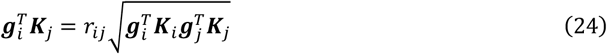

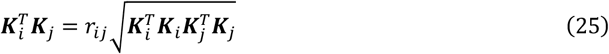

where *r*_*ij*_ is the LD between variant *i* and *j*. Therefore, combining equation (23) and LD matrix from reference panel or in-sample LD matrix, sufficient statistic ***Z***^*T*^***Z*** can be constructed and its semi-positive definiteness is guaranteed.

### Details in IBSS-ss in SuSiEgxe

The evidence lower bound (ELBO) of SuSiE model equations (1–7) can be obtained by maximizing the ELBO for the SER model *l* and its ELBO is defined as

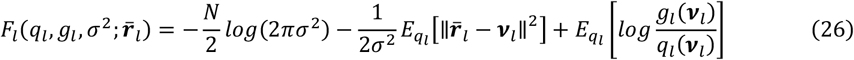

where ***v***_*l*_ = ***Z*ζ**_*l*_, **ζ**_*l*_ ~ *g*_*l*_, *g*_*l*_ is a pre-specified prior distribution for **ζ**_*l*_, *q*_*l*_ ∈ 𝒬 and 𝒬 is all possible distribution on 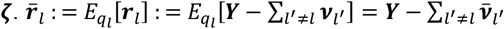 is expected residuals, where 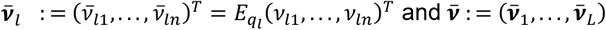 are expected value under *q*_*l*_.

The second term of equation (26) is the expected residual sum of squares (ERSS) under the variational approximation *q*_*l*_, which is derived as

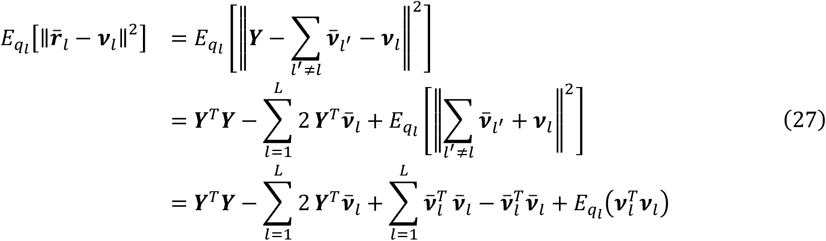

where 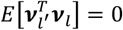 since the posterior mean for different *l* are independent to each other. 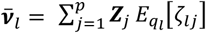, and 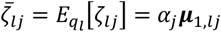 is the first moment of ζ_*lj*_. For the last term of equation (27),

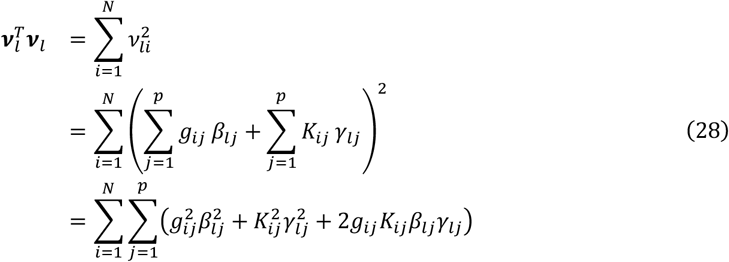

where 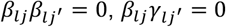 and 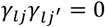 because only one element in **ζ**_*l*_ is non-zero under SER model *l*. Thus,

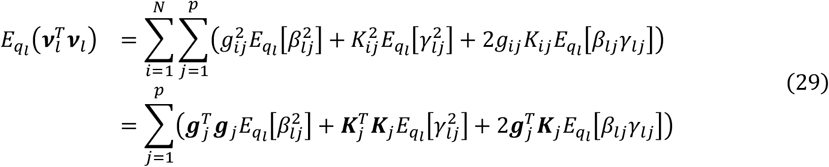

where the expectation terms are the second moment of ζ_*lj*_

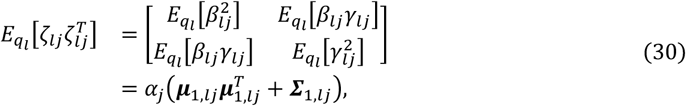

and 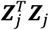 same as in equation (23)

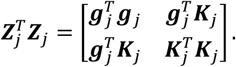

The third term of equation (26) is −*D*_*KL*_ (*q*_*l*_ ∥ *g*_*l*_), where *D*_*KL*_ (*q*_*l*_ ∥ *g*_*l*_) is Kullback–Leibler (KL) divergence from *q*_*l*_ to *g*_*l*_.

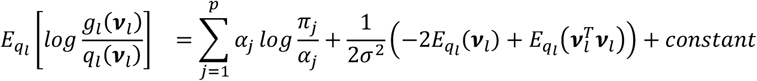

The second term is posterior expected log-likelihood for SER model *l*. The IBSS-ss algorithm for SuSiEgxe can be found in Algorithm 1.

#### Algorithm 1

IBSS-ss for Gene-environment interaction studies.

Require: Sufficient statistics 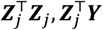, LD matrix from reference panel ***R***

Require: Number of effects, *L*; initial estimates of hyperparameters *σ*^2^, *∑*_0_.

Require: Initial estimates of the posterior mean single effects, 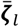, for *l* = 1, …, *L*.

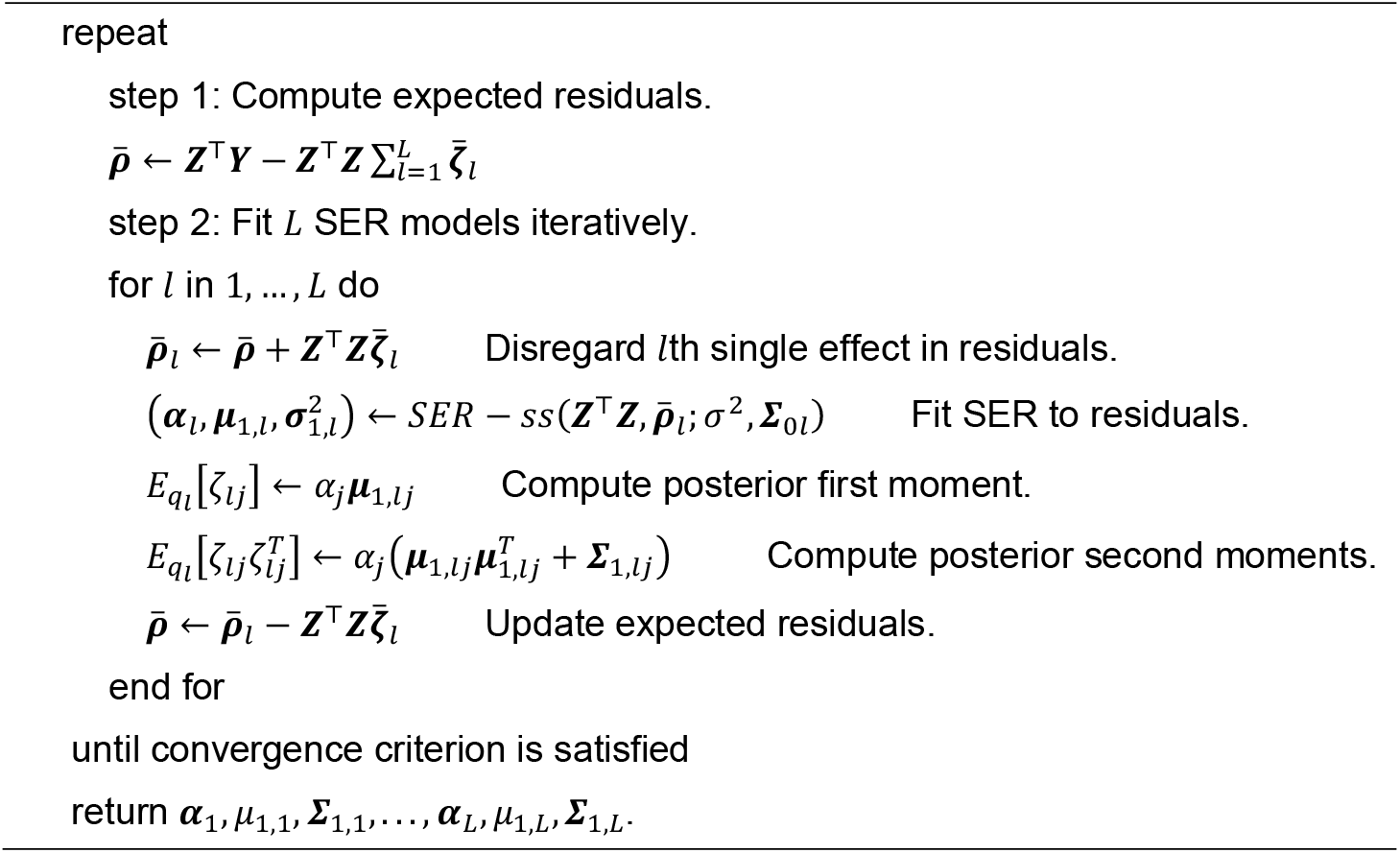

## Notes

### Author Declarations

IRB of The University of Texas Health Science Center at Houston gave ethical approval for this work.

