## Supplementary Notes for "Large-Scale Gene-Smoking Interactions and Fine Mapping Study Identifies Multiple Novel Blood Pressure Loci in over 1 Million Individuals"

### Supplemental Material

### 1. Study Descriptions

**AGES (Age Gene/Environment Susceptibility Reykjavik Study)**: The AGES Reykjavik study originally comprised a random sample of 30,795 men and women born in 1907-1935 and living in Reykjavik in 1967. A total of 19,381 people attended, resulting in a 71% recruitment rate. The study sample was divided into six groups by birth year and birth date within month. One group was designated for longitudinal follow up and was examined in all stages; another was designated as a control group and was not included in examinations until 1991. Other groups were invited to participate in specific stages of the study. Between 2002 and 2006, the AGES Reykjavik study re-examined 5,764 survivors of the original cohort who had participated before in the Reykjavik Study.

**ARIC (Atherosclerosis Risk in Communities):** The ARIC study is a population-based prospective cohort study of cardiovascular disease sponsored by the National Heart, Lung, and Blood Institute (NHLBI). ARIC included 15,792 individuals, predominantly European American and African American, aged 45-64 years at baseline (1987-89), chosen by probability sampling from four US communities. Cohort members completed three additional triennial follow-up examinations, a fifth exam in 2011-2013, a sixth exam in 2016-2017, and a seventh exam in 2018-2019. Several additional visits have occurred since Visit 7, with Visit 11 currently in progress and expected to conclude in August 2025. Three sequential recordings for systolic and diastolic blood pressure were obtained from participants in a seated position after a five-minute rest period. Cigarette smoking status and consumption data were collected via a standardized questionnaire. Informed consent is obtained upon the participant's arrival at the clinic. The ARIC study has been described in detail previously (PMC8667593; Wright JD, Folsom AR, Coresh J, et al. The ARIC (Atherosclerosis Risk In Communities) Study: JACC Focus Seminar 3/8. J Am Coll Cardiol. 2021 Jun 15;77(23):2939-2959).

**ASCOT - Anglo-Scandinavian Outcomes Trial:** This study comprises hypertensive cases from the Anglo-Scandinavian Cardiac Outcomes Trial. For BP ascertainment: 160mmHg SBP or 100mmHg DBP untreated; or 140mmHg SBP or 90mmHg DBP treated with one or more antihypertensive therapy. Patients were recruited in the UK and Ireland and Scandinavia, and were 40 years of age or greater and had 3 additional cardiovascular risk factors. Exclusions were contraindications to antihypertensive therapies; secondary or malignant hypertension; previous clinical myocardial infarction or current treated angina; cerebrovascular event within previous 3 months; heart failure; uncontrolled arrhythmias; clinically important haematological or biochemical abnormalities. Sample selection for the GxL projects was based on available datasets for traits and lifestyle measures.

**ASPREE (ASPirin in Reducing Events in the Elderly):** The ASPREE study (n=19,114) is a randomized double-blind, placebo-controlled clinical trial, which aimed to determine whether daily 100-mg aspirin extended disability-free survival in healthy adults aged 70 years and older (65 years of age and older for U.S. minorities) with no history of diagnosed cardiovascular diseases, dementia, physical disability, or other life-threatening illness at enrolment. The design and protocol of the ASPREE trial have been reported previously.[1-4] All participants provided written informed consent. The study was approved by the Monash University and Alfred Hospital Human Research Ethics Committee (390/15) in Australia and site-specific Institutional Review Boards in the United States and registered on Clinicaltrials.gov (NCT01038583).

Out of the 19,114 ASPREE trial participants, a total of 12,031 genotyped, unrelated participants (genetic relationship <0.05) aged ≥70 years with European ancestry (determined by genetic principal component analysis) were included in the CHARGE gene-lifestyle interaction studies.

1. McNeil, J.J., Woods, R.L., Nelson, M.R. et al., 2017. Baseline characteristics of participants in the ASPREE (ASPirin in Reducing Events in the Elderly) study. *The Journals of Gerontology: Series A*, *72*(11), 1586-1593.
2. McNeil, J.J., Woods, R.L., Nelson, M.R. et al., 2018. Effect of aspirin on disability-free survival in the healthy elderly. *New England Journal of Medicine*, *379*(16), 1499-1508.
3. McNeil, J.J., Wolfe, R., Woods, R.L. et al., 2018. Effect of aspirin on cardiovascular events and bleeding in the healthy elderly. *New England Journal of Medicine*, *379*(16), 1509-1518.
4. McNeil, J.J., Nelson, M.R., Woods, R.L. et al., 2018. Effect of aspirin on all-cause mortality in the healthy elderly. *New England Journal of Medicine*, *379*(16), 1519-1528.

**ASPS (The Austrian Stroke Prevention Study):** The ASPS study is a single center prospective follow-up study on the effects of vascular risk factors on brain structure and function in the normal elderly population of the city of Graz, Austria. The procedure of recruitment and diagnostic work-up of study participants has been described previously [1,2]. A total of 2007 European Caucasian participants were randomly selected from the official community register stratified by gender and 5 year age groups. Individuals were excluded from the study if they had a history of neuropsychiatric disease, including previous stroke, transient ischemic attacks, and dementia, or an abnormal neurologic examination determined on the basis of a structured clinical interview and a physical and neurologic examination. During two study periods between September 1991 and March 1994 and between January 1999 and December 2003 an extended diagnostic work-up including magnetic resonance imaging and neuropsychological testing was done in 1076 individuals aged 45 to 85 years randomly selected from the entire cohort: 509 from the first period and 567 from the second. Genotyping was performed in 996 participants. The study protocol was approved by the ethics committee of the Medical University of Graz, Austria, and written informed consent was obtained from all subjects.

[1] Schmidt R, Fazekas F, Kapeller P, Schmidt H, Hartung HP. Mri white matter hyperintensities: Three
year follow-up of the austrian stroke prevention study. *Neurology*. 1999;53:132-139

[2] Schmidt R, Lechner H, Fazekas F, Niederkorn K, Reinhart B, Grieshofer P, et al. Assessment of
cerebrovascular risk profiles in healthy persons: Definition of research goals and the austrian stroke prevention study (asps). *Neuroepidemiology*. 1994;13:308-313

**ASPS-Fam (Austrian Stroke Prevention Family Study):** The Austrian Stroke Prevention Family Study (ASPS-Fam) is a prospective single-center, community-based study on the cerebral effects of vascular risk factors in the normal elderly population of the city of Graz, Austria [1,2]. The ASPS-Fam represents an extension of the Austrian Stroke Prevention Study (ASPS), which was established in 1991 [3,4]. Between 2006 and 2013, study participants of the ASPS and their first grade relatives were invited to enter ASPS-Fam. Inclusion criteria were no history of previous stroke or dementia and a normal neurologic examination. A total of 419 individuals from 176 families were included into the study. The number of members per family ranged from two to six. The entire cohort underwent an extended diagnostic workup including clinical history, blood tests, cognitive testing, magnetic resonance imaging and a thorough vascular risk factor assessment. The study protocol was approved by the ethics committee of the Medical University of Graz, Austria, and written informed consent was obtained from all subjects.

[1] Seiler, S. et al. Magnetization transfer ratio relates to cognitive impairment in normal elderly. Frontiers in aging neuroscience 6, 263, doi:10.3389/fnagi.2014.00263 (2014).

[2] Ghadery, C. et al. R2* mapping for brain iron: associations with cognition in normal aging. Neurobiology of aging 36, 925-932, doi:10.1016/j.neurobiolaging.2014.09.013 (2015).

[3] Schmidt, R. et al. Assessment of cerebrovascular risk profiles in healthy persons: definition of research goals and the Austrian Stroke Prevention Study (ASPS). Neuroepidemiology 13, 308-313, doi:10.1159/000110396 (1994).

[4] Schmidt, R., Fazekas, F., Kapeller, P., Schmidt, H. & Hartung, H. P. MRI white matter hyperintensities: three-year follow-up of the Austrian Stroke Prevention Study. Neurology 53, 132-139 (1999).

**AugUR:** The German AugUR study is a prospective cohort study of the general old-aged population in and around the city of Regensburg with a focus on general chronic diseases. AugUR included 2,449 individuals, predominantly Europeans, aged 70-95 years at baseline (recruited between 2013 and 2019), with ongoing follow-up examinations. The study protocol included a standardized interview regarding social and life-style factors, medication history, quality-of-life, and existing diagnoses of common diseases. The participants were subjected to medical examinations and general measurements of body shape and fitness. Standard laboratory measurements were performed on fresh and biobanked blood and urine samples. The design of the AugUR study has been described previously (Stark K, Olden M, Brandl C, et al. The German AugUR study: study protocol of a prospective study to investigate chronic diseases in the elderly. BMC Geriatr 2015;15).

**AWI-Gen (The Africa Wits-INDEPTH partnership for Genomic Studies):** The Africa Wits-INDEPTH partnership for Genomic Studies (AWI-Gen) is a National Institutes of Health (NIH) funded Collaborative Centre of the Human Heredity and Health in Africa (H3Africa) Consortium investigating the genomic and environmental risk factors for cardiometabolic diseases in Africans. The study was cross-sectional in nature and was investigating populations from six sub-Saharan African sites in four countries (Burkina Faso, Ghana, Kenya and South Africa) in East, West and Southern Africa. Study participants provided informed consent and were enrolled for data collection from 2012 to 2016. Over 12 000 sub-Saharan Africans were enrolled form rural and urban settings and aged from 40 to 60 years with a parity between male and female individuals[1–3]. All participants completed a questionnaire with questions on demography, health history and behaviour. Anthropometric measurements were taken and fasting venous blood samples were collected for genotyping (H3Africa array) and phenotyping (biomarkers) purposes as described in the AWI-Gen resource paper and the detailed AWI-Gen methods paper[1–3]. Ultrasound scans were performed to assess the right and left common carotid intima-media thickness (cIMT).

Smoking data were collected using the CAGE questionnaire and previously published [4–6]. Blood pressure traits were defined as previously described[7]. All data were redefined to fit with the CHARGE-GLI analysis plan.

The H3Africa genotyping array, designed as an African-common-variant-enriched GWAS array (Illumina) with ~2.3 million SNPs (https://www.h3abionet.org/h3africa-chip), was used to genotype genomic DNA using the Illumina FastTrack Service. The following pre-imputation QC steps were applied to the entire AWI-Gen genotype data set. Individuals with a missing SNP calling rate greater than 0.05 were removed. SNPs with a genotype missingness greater than 0.05, MAF less than 0.01 and failing Hardy-Weinberg equilibrium (HWE) at a P-value less than 0.0001 were removed. Non-autosomal and mitochondrial SNPs, and ambiguous SNPs that did not match the GRCh37 references alleles or strands were also removed. Imputation was performed on the cleaned dataset (with 1,729,661 SNPs and 10,903 individuals) using the Sanger Imputation Server and the African Genome Resource as reference panel. EAGLE2 was used for pre-phasing and the default PBWT algorithm was used for imputation. After imputation, poorly imputed SNPs with info scores less than 0.6, MAF less 0.01, and HWE P-value less than 0.00001 were excluded using the automated Nextflow pipeline h3abionet/h3gwas [8].

The AWI-Gen study was approved by the Wits Human Research Ethics Committee (Medical) under clearance numbers M121026, M16021 and M170880. All enrolled participants signed informed consent and study procedures were compliant with the Declaration of Helsinki.

**BBJ (Biobank Japan Project):** The BBJ is a national biobank project and a nationwide hospital-based genome cohort that enrolled patients from 12 collaborating medical institutions since 2013 (https://biobankjp.org/en/index.html). The BBJ consists of DNA samples and clinical data, including laboratory data, related to 47 target diseases from approximately 200,000 patients (Nagai et al, J Epidemiol 2017). Genotyping was performed on the Illumina HumanOmniExpressExome BeadChip or a combination of the Illumina HumanOmniExpress and HumanExome BeadChips (Terao et al, Nature 2020). Genotypes were imputed using our new reference panel developed by using 3,256 high depth whole genome sequencing data from Japanese population combined with the 1000 Genome project (Tanaka et al, JACI 2021). All patients recruited for the BioBank Japan Project provided written informed consent. All relevant ethical regulations were complied with throughout this study.

**BES (Beijing Eye Study):** The Beijing Eye Study is a population-based study that assesses the associated and risk factors of ocular and general diseases in a Chinese population. The study was initialized in 2001 and collected data from 4439 subjects aged ≥ 40 years and living in seven communities in the Beijing area. Three of these communities were located in a rural district and four were located in an urban district. The BES was followed-up in 2006, with 3251 of the original subjects participating, and in 2011, with 2695 subjects returning for the follow-up examination. At the examinations in 2006 and 2011, trained research staffs asked the subjects questions from a standard questionnaire providing information on the family status, level of education, income, quality of life, psychic depression, physical activity, and known major systemic diseases. Fasting blood samples were taken for measurement of concentrations of substances such as blood lipids, glucose, and glycosylated hemoglobin. Individuals were classified as self-reported non-smokers or self-reported current smokers. Alcohol consumption habits based on number of drinks per day were collected. Physical activity was assessed in questions on the number of hours per day and number of days per week spent on intensively or moderately performed sport activities, spent on walking, on riding a bicycle, and spent on sitting. All variables used in analyses were taken from examinations in 2006 or in 2011. The BES subjects were genotyped on two arrays, Illumina Human610-Quad (N = 832) and Illumina OmniExpress (N = 814).

**BioMe:** The BioMe Biobank is an ongoing, prospective, hospital- and outpatient- based population research program operated by The Charles Bronfman Institute for Personalized Medicine (IPM) at Mount Sinai. BioMe has enrolled over 60,000 participants since September 2007. BioMe is an Electronic Medical Record (EMR)-linked biobank that integrates research data and clinical care information for consented patients at The Mount Sinai Medical Center, which serves diverse local communities of upper Manhattan with broad health disparities. BioMe includes individuals of African ancestry (AA, 25%), Hispanic Latino ancestry (HL, 36%), European ancestry (EA, 30%), and of other ancestry (9%). The BioMe disease burden is reflective of health disparities in the local communities. BioMe operations are fully integrated in clinical care processes, including direct recruitment from clinical sites waiting areas and phlebotomy stations by dedicated BioMe recruiters independent of clinical care providers, prior to or following a clinician standard of care visit. Recruitment currently occurs at a broad spectrum of over 30 clinical care sites.

**BioVU (VUMC):** BioVU is Vanderbilt's biorepository of de-identified samples extracted from discarded blood collected during routine clinical testing and linked to de-identified medical records in the Synthetic Derivative (SD). The SD consists of de-identified clinical data obtained from patients attending all clinics associated with the Vanderbilt University Medical Center hospital system. Clinical data are abstracted from multiple sources including diagnostic and procedure codes, basic demographics, discharge summaries, progress notes, health history, multi-disciplinary assessments, laboratory values, imaging reports, medication orders, and pathology reports.

Blood pressure measures extracted from EHR records reflect the earliest outpatient measurements taken after and within 1-year exposure assessment in adults >= 18 years of age. When measurements were available for multiple days for the earliest time since exposure measurement, a visit was randomly chosen. When the chosen visit had multiple measurements per-day, an average measurement was computed.

**BRIGHT (British Genetics of Hypertension):** Participants of the BRIGHT Study were recruited from the Medical Research Council General Practice Framework and other primary care practices in the UK. Each case had a history of hypertension diagnosed prior to 60 years of age with confirmed blood pressure recordings corresponding to seated levels >150/100mmHg (1 reading) or mean of 3 readings >145/95 mmHg. BRIGHT is focused on recruitment of hypertensive individuals with BMI<30. Sample selection for the GxL projects was based on available datasets for traits and lifestyle measures.

**CAGE-Amagasaki (Cardio-metabolic Genome Epidemiology Network, Amagasaki Study):** The Amagasaki Study (CAGE-Amagasaki) is an ongoing population-based cohort study of 5,743 individuals (3,435 males and 2,310 females), aged >18 years and recruited for a baseline examination between September 2002 to August 2003. Participants were interviewed by trained personnel to obtain information on medical and lifestyle variables, and consented to provide DNA for genotyping of molecular variants to investigate genetic susceptibility for so-called lifestyle-related diseases such as hypertension and cardiovascular disorder.

**CAGE-KING (Kita-Nagoya Genomic Epidemiology Study):** The Kita-Nagoya Genomic Epidemiology (KING) study (ClinicalTrials.gov identifier: NCT00262691) is an ongoing community-based prospective observational study of the genetic basis of cardiovascular disease and its risk factors[1]. The study collected 3,975 Japanese subjects aged 50-80 years, who underwent community-based annual health check-ups between May 2005 and December 2007. Part of the KING Study samples (n=998) were included in the GWAS analysis. Systolic blood pressure and diastolic blood pressure were measured using the USM-700G machine (Elquest Co., Japan) in the seated position. The mean of two readings was used for the analysis.

1. Asano, H. et al. Plasma resistin concentration determined by common variants in the resistin gene and associated with metabolic traits in an aged Japanese population. Diabetologia 53, 234-46 (2010).

**CARDIA (The Coronary Artery Risk Development in Young Adults):** CARDIA is a prospective multicenter study of the development and determinants of clinical and subclinical cardiovascular disease and their risk factors. It began in 1985-1986 with a group of 5,115 black and white men and women aged 18-30 years, recruited from four US centers. The recruitment was done from the total community in Birmingham, AL, from selected census tracts in Chicago, IL and Minneapolis, MN; and from the Kaiser Permanente health plan membership in Oakland,

CA. The participants were selected so that there would be approximately the same number of people in subgroups of race, gender, education (high school or less and more than high school) and age (18-24 and 25-30). These same participants were asked to participate in follow-up examinations during 1987-1988 (Year 2), 1990-1991 (Year 5), 1992-1993 (Year 7), 1995-1996 (Year 10), 2000-2001 (Year 15), 2005-2006 (Year 20), 2010-2011 (Year 25), 2015-2016 (Year 30), and 2020-2022 (Year 35).

Data collection protocols have been described previously (Friedman, 1988). Weight, height, systolic blood pressure and diastolic blood pressure were measured using standard protocols with participants wearing light clothing and no shoes. Morning venous blood samples were obtained after an overnight fast of at least 8 hours. Total cholesterol and triglycerides were measured enzymatically, HDL-C was determined by precipitation with dextran sulfate magnesium chloride, and LDL cholesterol was calculated using the Friedewald equation. Fasting plasma glucose and insulin were measured with the hexokinase–ultraviolet and radioimmunoassay methods, respectively (Linco Research, St Charles, MO, USA).

Written informed consent was obtained from participants at each examination and all study protocols were approved by the institutional review boards of the participating institutions.

Friedman GD, Cutter GR, Donahue RP, Hughes GH, Hulley SB, Jacobs DR Jr, Liu K, Savage PJ. CARDIA: study design, recruitment, and some characteristics of the examined subjects. J Clin Epidemiol. 1988;41(11):1105-16. PMID: 3204420.

**CCHC (Cameron County Hispanic Cohort):** The CCHC (Cameron County Hispanic Cohort) is an ongoing, longitudinal study that was initiated in 2004 to investigate the burden of metabolic-related conditions and disparities in a Mexican American community. The ongoing study has recruited over 5000 participants through community-based research assistants who through a randomization process contact families living at the US-Mexico border in Cameron County, TX to voluntarily enroll in the program. They are then invited to visit the Clinical Research Unit located in facilities provided by Valley Baptist Medical Center, Brownsville, TX. Comprehensive clinical examinations and collection of specimens have been conducted and archived for further analysis. A subset of 4076 individuals have been genotyped using the Illumina MEGA array and the results have been imputed to TOPMed phase 8 reference data.

**CFS (The Cleveland Family Study)** is the largest family-based study of sleep apnea worldwide, consisting of 2,284 individuals (46% African American) from 361 families studied on up to 4 occasions over a period of 16 years. The study was begun in 1990 with the initial aims of quantifying the familial aggregation of sleep apnea. NIH renewals provided expansion of the original cohort (including increased minority recruitment) and longitudinal follow-up, with the last exam occurring in February 2006. 632 African Americans were genotyped on the Affymetrix array 6.0 platform through the CARe Consortium with suitable genotying quality control. A further 122 African-Americans had genotyping based on the Illumina OmniExpress + Exome platform. Genomes were imputed by TOPMed imputation server separately.

**CHS (Cardiovascular Health Study):** CHS is a population-based cohort study of risk factors for cardiovascular disease in adults 65 years of age or older conducted across four field centers (1). The original predominantly European ancestry cohort of 5,201 persons was recruited in 1989-1990 from random samples of the Medicare eligibility lists and an additional predominately African-American cohort of 687 persons was enrolled in 1992-93 for a total sample of 5,888. Blood samples were drawn from all participants at their baseline examination and DNA was subsequently extracted from available samples. European ancestry participants were excluded from the GWAS study sample due to prevalent coronary heart disease, congestive heart failure, peripheral vascular disease, valvular heart disease, stroke, or transient ischemic attack at baseline. After QC, genotyping was successful for 3271 European ancestry and 823 African-American participants. CHS was approved by institutional review committees at each site and individuals in the present analysis gave informed consent including consent to use of genetic information for the study of cardiovascular disease.

1. Fried LP, Borhani NO, Enright P, Furberg CD, Gardin JM, Kronmal RA, et al. The Cardiovascular Health Study: design and rationale. Ann Epidemiol 1991; 1:263-76.

**CILENTO STUDY:** The Cilento study is a population-based, prospective study including isolated populations from three villages (Campora, Gioi and Cardile) located in an internal hilly area of the "Cilento National Park, Vallo di Diano and Alburni" (http: //www.igb.cnr.it/cilento). The study aim has been to identify genetic risk factors for common diseases and related quantitative traits. Using local church and town records, exhaustive genealogies of these populations were reconstructed and a large population pedigree spanning 15-17 generations was reconstructed for each of the three populations. Using genealogical and genetic data, we have demonstrated that those populations present all features of young genetic isolates (1-3).

The Cilento study started in 2003 and includes 2,331 individuals aged 1-103 years. At baseline, information on personal and familial history of diseases, past and present drug therapies, diet, smoking habit and alcohol use have been collected by questionnaires. In addition, samples of fasting blood, plasma, serum, and PBMCs were collected. All samples were genotyped using the Illumina (370K, Omni-express) chips, exome data are also available for a subset of samples. Participants underwent blood tests with measure of several biochemical parameters, an extensive physical examination, including anthropometry, blood pressure, ECG, echocardiography, and measurements of the carotid artery intima-media thickness by ultrasonography. Further, participants underwent bone density test, dental examination, and a neurological examination including cognitive tests.

The Cilento study was approved by the ethics committee of “Azienda Sanitaria Locale Napoli 1” and the ethics committee “Carlo Romano” University of Naples “Federico II”. The study was conducted according to the criteria set by the declaration of Helsinki and each subject signed an informed consent before participating in the study.

1. V. Colonna et al. Campora: a young genetic isolate in South Italy. Hum Hered. 2007;64,123-35

2. V. Colonna et al. Comparing population structure as inferred from genealogical versus genetic information Eur. J. Hum. Genetics 2009 Dec;17(12):1635-41
3. T. Nutile, et al. Whole-Exome Sequencing in the Isolated Populations of Cilento from South Italy. Sci Reports 2019 Mar 11;9(1):4059. doi: 10.1038/s41598-019-41022-6

**CoLaus:** The CoLaus study is a population-based cohort of 6733 participants from Lausanne (Switzerland) aged 35-75 at recruitment (2003-2006). This study collects detailed and standardized phenotypes to investigate biological, genetic and environmental determinants of cardiovascular disease and their association with mental disorders. Initiated in 2003, the main aims of the CoLaus study were: 1) To better understand the epidemiology of cardiovascular risk factors and diseases in the Swiss population; 2) To discover new genetic determinants of these conditions. 3) To determine the nature of the associations between cardiovascular disease and mental disorders. Since then, three follow-up surveys have been completed (2009-2012, 2014-2016, 2018-2020) and a fourth one is ongoing since 2022. A detailed description of the participants' selection, inclusion criteria and methods used can be found in the following manuscript:
*Firmann M, Mayor V, Marques Vidal P, Bochud M, Pécoud A, Hayoz D, Paccaud F, Preisig M, Song KS, Yuan X, et al. The CoLaus study: a population-based study to investigate the genetic determinants of cardiovascular risk factors and metabolic syndrome. BMC Cardiovascular Disorders. 2008; 8:6. PMID: 18366642; PMCID: PMC2311269; DOI: 10.1186/1471-2261-8-6*

**CRONICAS:** The CRONICAS Cohort Study is an ongoing longitudinal study started in 2010 to compare the prevalence and risk factors of cardiovascular disease and chronic obstructive pulmonary disease, as well as to determine the rate of disease progression to hypertension and diabetes from a disease-free baseline status and the rate of lung function decline between populations. For this, 3601 participants from three Peruvian regions that differ by degree of urbanization, levels of indoor and outdoor pollution, and altitude were enrolled. The three regions include Lima, a highly urbanized setting at sea level; Tumbes, a semi-urban setting at sea level in the north of Peru; and Puno, a region in the highlands with urban and rural settings. A subset of 685 participants from the urban and rural settings in Puno were genotyped and included in these analyses. Details of the CRONICAS Cohort Study are reported in BMJ Open[1].

1. Miranda JJ, Bernabe-Ortiz A, Smeeth L, Gilman RH, Checkley W, CRONICAS Cohort Study Group. Addressing geographical variation in the progression of non-communicable diseases in Peru: the CRONICAS cohort study protocol. BMJ open. 2012 Jan 1;2(1):e000610.

**DHS (Diabetes Heart Study):** The Diabetes Heart Study (DHS) is an ongoing family-based cohort study investigating the epidemiology and genetics of cardiovascular disease (CVD) in a populationbased sample. The DHS recruited T2D-affected siblings without advanced renal insufficiency from 1998 through 2005 in western North Carolina. DHS has collected genetic data on 1,220 self-described European American (EA) individuals from 475 families. Genotyping was completed using an Affymetrix Genome-Wide Human SNP Array 5.0.

**DHS-AA (African American Diabetes Heart Study):** AA-DHS objectives are to improve understanding of ethnic differences in CAC and CP in populations of African and European ancestry. The AA-DHS consists of self-reported African Americans with T2D recruited from two Wake Forest School of Medicine (WFSM) studies: the family-based Diabetes Heart Study (DHS) and unrelated individuals in the AA-DHS. DHS is a cross-sectional study of European American and African American families with siblings concordant for T2D. AA-DHS started after DHS and enrolled unrelated African Americans. The AA-DHS GWAS utilized the Illumina 5M chip.

**DIACORE:** DIACORE (DIAbetes COhoRtE) is a German prospective cohort study of 3,000 adult European diabetes mellitus type 2 outpatients, with a primary focus on renal events. Four follow-up examinations were conducted within 10 years after baseline inclusion. At each visit including, a core phenotyping protocol was performed that included a standardized interview, a physical examination, the determination of standard laboratory parameters from serum, urine and whole blood in a central laboratory and biobanking of biomaterials. The design of the DIACAORE study (Dorhofer L, Lammert A, Krane V, et al. Study design of DIACORE (DIAbetes COhoRtE)—a cohort study of patients with diabetes mellitus type 2. BMC Med Genet. 2013;14:25) and exemplary results (Rheinberger M, Jung B, Segiet T, et al. Poor risk factor control in outpatients with diabetes mellitus type 2 in Germany: The DIAbetes COhoRtE (DIACORE) study. PLoS ONE 2019;14(3): e0213157) were published previously.
DR's EXTRA (Dose-Responses to Exercise Training): The Dose-Responses to Exercise Training (DR's EXTRA) study is a 4-year RCT on the health effects of regular physical exercise and a healthy diet in a population-based random sample of Finnish men and women aged 55-74 years living in the city of Kuopio in 2002. The 3000 men and women who were invited to participate in the study were obtained from the national population registry. Altogether 2062 men and women expressed their willingness to participate in the study, and 1479 of them attended the baseline measurements in 2005 - 2006. The prespecified exclusion criteria were medical or other conditions that prohibit engagement in exercise intervention or the assessments, as judged by a physician. After these exclusions, 1410 individuals aged 57-78 years at baseline in 2005-2006 were randomized into the resistance exercise, aerobic exercise, diet, combined resistance exercise and diet, combined aerobic exercise and diet, or control group.

**ELISABET (Enquête LItoral Souffle Air Biologie EnvironemenT):** The ELISABET (Enquête LItoral Souffle Air Biologie EnvironemenT) Study is a population base study, including a representative sample of 1607 inhabitants of Dunkirk area and 1668 inhabitants of Lille Area, in northern France. Subjects were agef from 40 to 64 and were recruited between 2011 and 2013. The mains objectives of the study were to study the association between air pollution and airway obstruction and the prevalence of cardiovascular risk factors.

1.Quach A, Giovannelli J, Cherot-Kornobis N, Ciuchete A, Clement G, Matran R, et al. Prevalence and underdiagnosis of airway obstruction among middle-aged adults in northern France: The ELISABET study 2011-2013. Respir. Med. 2015;109(12):1553 61.

**EXCEED:** EXCEED is a longitudinal population-based cohort which facilitates investigation of genetic, environmental and lifestyle-related determinants of a broad range of diseases and of multimorbidity through data collected at baseline, via electronic healthcare records linkage and recurrent self-completion questionnaires. Launched in 2013, EXCEED initially recruited males and females, aged 30-69 and living in Leicester, Leicestershire and Rutland via smoking clinics and GP practices. In May 2020, recruitment opened to anyone over the age of 18 years, living in either the East or West Midlands and registration and consent are now completed online via the EXCEED website. To date, some 11,200 participants have consented to follow their electronic health records for up to 25 years, storage and analysis of their DNA sample and to be contacted for 'recall-by-genotype' and 'recall-by-phenotype' sub-studies.
Cohort Profile paper: John et al. 2019 Int J Epidemiol https://doi.org/10.1093/ije/dyz073

**FHS (Framingham Heart Study):** FHS began in 1948 with the recruitment of an original cohort of 5,209 men and women (mean age 44 years; 55 percent women). In 1971 a second generation of study participants was enrolled; this cohort (mean age 37 years; 52% women) consisted of 5,124 children and spouses of children of the original cohort. A third-generation cohort of 4,095 children of offspring cohort participants (mean age 40 years; 53 percent women) was enrolled in 2002-2005 and are seen every 4 to 8 years. Details of study designs for the three cohorts are summarized elsewhere. At each clinic visit, a medical history was obtained with a focus on cardiovascular content, and participants underwent a physical examination including measurement of height and weight from which BMI was calculated.

**FUSION (Finland-United States Investigation of NIDDM Genetics):** The Finland-United States Investigation of NIDDM Genetics (FUSION) study is a long-term effort to identify genetic variants that predispose to type 2 diabetes (T2D) or that impact the variability of T2D-related quantitative traits. The FUSION GWAS sample consists of 1,161 Finnish T2D cases and 1,174 Finnish normal glucose-tolerant (NGT) controls (Scott et al. Science 2007). Cases are defined by fasting plasma glucose ≥ 7.0 mmol/l or 2-h plasma glucose ≥ 11.1 mmol/l, by report of diabetes medication use, or based on medical record review. 789 FUSION cases each reported at least one T2D sibling; 372 Finrisk 2002 T2D cases came from a Finnish population-based risk factor survey. NGT controls are defined by fasting glucose < 6.1 mmol/l and 2-h glucose < 7.8 mmol/l. FUSION controls include 119 subjects from Vantaa, Finland who were NGT at ages 65 and 70 years, 304 NGT spouses from FUSION families, and 651 Finrisk 2002 subjects. The controls were approximately frequency matched to the cases by age, sex, and birth province. Smoking and alcohol data are only available in the FUSION subset of our GWAS samples.

Scott, L.J. et al. A genome-wide association study of type 2 diabetes in Finns detects multiple susceptibility variants. Science 316, 1341–1345, 2007.

**GAPP - Genetic and phenotypic determinants of blood pressure and other cardiovascular risk factors:** GAPP is a population-based prospective cohort study involving a representative sample of initially healthy adults aged 25-41 years at baseline and residing in the Principality of Liechtenstein. Exclusion criteria were the presence of cardiovascular disease, diabetes, obstructive sleep apnea and a body mass index >35kg/m2. A standardized 12-lead ECG was obtained in all participants. Baseline characteristics were obtained in all participants using standard methodology. The main goal of GAPP is to assess the mechanisms for the development of cardiovascular risk factors over time.

**GENOA (Genetic Epidemiology Network of Arteriopathy):** The Genetic Epidemiology Network of Arteriopathy (GENOA) study, a part of the Family Blood Pressure Program (FBPP Investigators, 2002), consists of hypertensive sibships that were recruited for linkage and association studies in order to identify genes that influence blood pressure and its target organ damage (Daniels, 2004). In the initial phase of the GENOA study (Phase I: 1995-2000), all members of sibships containing ≥ 2 individuals with essential hypertension clinically diagnosed before age 60 were invited to participate, including both hypertensive and normotensive siblings. During the first exam, 1,583 European Americans from Rochester, MN and 1,854 African Americans from Jackson, MS were recruited. In the second phase of the GENOA study (Phase II: 2001-2004), 1,241 European American and 1,482 African American participants were successfully re-recruited to measure potential target organ damage due to hypertension. In a GENOA follow-up study (2009–2011), 752 African Americans participated in a third examination. Since African American probands in GENOA were recruited in part through the Jackson, MS field center for Atherosclerosis Risk in Communities (ARIC), we excluded ARIC participants from GENOA analysis.

FBPP Investigators. (2002) Multi-center genetic study of hypertension: The Family Blood Pressure Program (FBPP). *Hypertension* 39(1): 3-9.

Daniels, P.R., Kardia, S.L., Hanis, C.L., Brown, C.A., Hutchinson, R., Boerwinkle, E., Turner, S.T., and Genetic Epidemiology Network of Arteriopathy study. (2004). Familial Aggregation of Hypertension Treatment and Control in the Genetic Epidemiology Network of Arteriopathy (GENOA) Study. *Am J Med* 116(10): 676-681.

**The GenSalt Study:** The Genetic Epidemiology Network of Salt-Sensitivity (GenSalt) study is a unique NHLBI-sponsored family feeding-study designed to examine the interaction between genes and dietary sodium intake on BP. A detailed description of the GenSalt study design and participants has been reported previously[1]. Briefly, 3,142 participants from 633 Han families from rural, north China were ascertained through a proband with untreated pre-hypertension or stage-1 hypertension identified from a population-based BP screening. Individuals who had stage 2 hypertension, secondary hypertension, and a history of clinical cardiovascular disease or diabetes or were pregnant, heavy alcohol drinkers, or currently on a low-sodium diet or BP lowering medication were excluded from the study, with a total of 1,906 GenSalt probands and their siblings, spouses, and offspring eligible for the 7-day low sodium and 7-day high sodium dietary interventions. At baseline, a standard questionnaire was administered by a trained staff member to collect information on family pedigree, demographic characteristics, personal and family medical history, and lifestyle risk factors. Body weight and height were measured twice in light indoor clothing without shoes. Body mass index was calculated as weight in kilograms per height in square meters. BP was measured three times at the same time each morning during the three-day baseline examination by trained and certified. The mean of the 9 BP measures was used in subsequent analyses. Blood specimens were collected by venipuncture to measure lipids, creatinine, and other laboratory values. Among the 1,906 intervention participants, 1,881 underwent genome-wide genotyping and whole genome sequencing.


1. The GenSalt Collaborative Research Group. GenSalt: rationale, design, methods and baseline characteristics of study participants. *J Hyperten.*2007;21:639-646.
2. Perloff D, Grim C, Flack J, Frohglich ED, Hill M, McDonald M, Morgenstern BZ. Human blood pressure determination by sphygmomanometer. *Circulation.* 1993;88:2460–2470.


**GOLDN (Genetics of Diet and Lipid Lowering Network):** GOLDN is a multi-center family pharmacogenetic study that is investigating gene- environment interactions on lipid profiles. 1,200 subjects in extended pedigrees were measured before and after two environmental exposures: 1) a dietary fat challenge to assess genetic regulators of fat uptake and clearance and 2) a 3-week clinical trial of fenofibrate to assess pharmacogenetic influences on response to treatment. The goals of the study are to identify and characterize genetic loci that predict the lipid profile treatment responses. https://dsgweb.wustl.edu/PROJECTS/MP5.html

**HANDLS (Healthy Aging in Neighborhoods of Diversity across the Life Span):** HANDLS is a community-based, longitudinal epidemiologic study examining the influences of race and socioeconomic status (SES) on the development of age-related health disparities among a sample of socioeconomically diverse African Americans and whites. This unique study will assess over a 20-year period physical parameters and also evaluate genetic, biologic, demographic, and psychosocial, parameters of African American and white participants in higher and lower SES to understand the driving factors behind persistent black-white health disparities in overall longevity, cardiovascular disease, and cognitive decline. The study recruited 3,722 participants from Baltimore, MD with a mean age of 47.7 years, 2,200 African Americans and 1,522 whites, with 41% reporting household incomes below the 125% poverty delimiter.

Genotyping was done on a subset of self-reporting African American participants by the Laboratory of Neurogenetics, National Institute on Aging, National Institutes of Health (NIH). A larger genotyping effort included a small subset of self-reporting European ancestry samples. This research was supported by the Intramural Research Program of the NIH, NIA and the National Center on Minority Health and Health Disparities.

**HCHS/SOL:** The Hispanic Community Health Study / Study of Latinos (HCHS/SOL) is a multicenter prospective cohort of 16,000 Hispanic/Latino adults designed to investigate the role of acculturation in disease, and to identify other traits that impact Hispanic/Latino health. HCHS/SOL is the most diverse and comprehensive study of Hispanic/Latino health, with participants of Cuban, Puerto Rican, Dominican, Mexican or Central/South American origin.

Participants were recruited through four sites affiliated with San Diego State University, Northwestern University in Chicago, Albert Einstein College of Medicine in Bronx, New York, and the University of Miami, using a census block and household sampling design. Study participants who were self-identified Hispanic/Latino and aged 18-74 years underwent extensive psycho-social, clinical assessments, and biospecimen collection during the baseline visit (2008-2011). A re-examination of the HCHS/SOL cohort was conducted during 2015-2017, and visit 3, which began in 2020, and was completed in 2024 with sample size 16,415. Annual telephone follow-up interviews have been conducted since study inception to determine health outcomes of interest. (dbGaP study accession number: phs000555).

**HRS (Health & Retirement Study):** The Health and Retirement Study (HRS) is a longitudinal survey of a representative sample of Americans over the age of 50. The total sample ever interviewed consists of over 43,000 persons in 26,500 households. Respondents are interviewed every two years about income and wealth, health and use of health services, work and retirement, and family connections. DNA was extracted from saliva collected during a face-to-face interview in the respondents' homes. These data represent respondents who provided DNA samples and signed consent forms in 2006, 2008, 2010, and 2012. Respondents were removed if they had missing genotype or phenotype data.

1. Juster, F. T., Suzman, R. (1995). An Overview of the Health and Retirement Study, Journal of Human Resources 30:Suppl: S7-S56.
2. Sonnega A, Faul JD, Ofstedal MB, Langa KM, Phillips JWR, Weir DR. Cohort Profile: the Health and Retirement Study (HRS). Int. J. Epidemiol. 2014; 43 (2): 576-585. PMID: 24671021.
3. Crimmins, E.M., Faul, J.,Guyer H., Langa K.M., Ofstedal M.B., Sonnega, A., Wallace R.B., and Weir D.R. (2013). Documentation of Biomarkers in the 2006 and 2008 Health and Retirement Study. HRS Documentation Report DR-012. https://hrsdata.isr.umich.edu/sites/default/files/documentation/data-descriptions/Biomarker2006and2008%20%281%29.pdf

**HTO Study:** 225 families were collected between 1993 and 1996 in the Oxfordshire region of the United Kingdom, for a quantitative family-based genetic study of hypertension and cardiovascular risk. The study complied with the Declaration of Helsinki, informed consent was obtained from all participants, and Research Ethics Committee approval was obtained. Nuclear families containing a sibship of at least three persons eligible to participate were recruited, via a proband family member with blood pressure in the top 5% of the blood pressure distribution ascertained quantitatively, or with hypertension confirmed clinically and requiring at least two drugs for satisfactory control. Baseline phenotyping included cardiovascular risk factors, anthropometry and ambulatory blood pressure monitoring. Between 1997 and 2000, participants were recalled for ECG, Echocardiogram and Carotid IMT measurements. Between 2005 and 2007, they were recalled for Cardiac MRI measurements. GWAS genotyping was performed in

2013 using Illumina 660W-Quad chips containing 557,124 SNPs. Biomarkers were determined in EDTA plasma by Nightingale Health.

**HUFS (Howard University Family Study)**: HUFS followed a population-based selection strategy designed to be representative of African American families living in the Washington, DC metropolitan area. The major objectives of the HUFS were to study the genetic and environmental basis of common complex diseases including hypertension, obesity, and associated phenotypes. Participants were sought through door-to-door canvassing, advertisements in local print media and at health fairs and other community gatherings. In order to maximize the utility of this cohort for the study of multiple common traits, families were not ascertained based on any phenotype. During a clinical examination, demographic information was collected by interview.

**HyperGEN (Hypertension Genetic Epidemiology Network):** HyperGEN is a family-based study that looks at the genetic causes of hypertension and related conditions in individuals of EUR and AFR ancestry groups. HyperGEN recruited hypertensive sibships, along with their normotensive adult offspring, and an age-matched random sample. HyperGEN has collected data on 2,471 EUR and 2,300 AFR ancestry groups, from five field centers in the United States (Alabama, Massachusetts, Minnesota, North Carolina, and Utah).

**INGI-CAR** (INGI-Carlantino) cohort consists of samples collected in a small village of the same name, Carlantino, located in the Puglia region, in Southern Italy. This cohort belongs to the Italian Network of Genetic Isolates (INGI), a project that aims to study the genetic basis of complex diseases and phenotypes, taking advantage of these populations' genetic and environmental homogeneity (doi: 10.1038/s41431-019-0551-x). Participants were voluntarily enrolled in the range from one to 90 years of age. Demographic data and detailed familial and individual medical history were collected for all the participants. In addition, participants underwent specialist evaluations, e.g., cardiovascular, in-depth sensorial, etc. Genotyping and phenotypic data are available for 630 individuals. All participants signed a written informed consent to the study before their enrollment; parents or legal guardians signed the informed consent for underage participants. The project was approved by the local administration of Carlantino, the Health Service of Foggia Province, Italy, and the ethical committee of the IRCCS Burlo-Garofolo of Trieste, Italy. All study procedures were performed under the ethical principles expressed by the Helsinki Declaration.

**INGI-FVG (INGI-Friuli Venezia Giulia)** is a population-based study started in 2008 with follow-up in 2014/2015, carried out in six villages of the Friuli Venezia Giulia (FVG) region, North-Eastern of Italy (Erto-Casso, Clauzetto, Illegio, Sauris, San Martino del Carso and Val di Resia). These populations belong to the Italian Network of Genetic Isolates (INGI), a project that aims to study the genetic basis of complex diseases and phenotypes, taking advantage of these populations' genetic and environmental homogeneity (doi: 10.1038/s41431-019-0551-x). Demographic data and detailed familial and individual medical history were collected for all the voluntarily enrolled participants. In addition, participants underwent specialist evaluations, e.g., cardiovascular, neurological, psychiatric, in-depth sensorial, stomatological, etc. All pathologies were classified according to the International Classification of Diseases-10 (ICD-10). The cohort accounts for 1927 genotyped samples from three to 92 years of age. All participants signed a written informed consent to the study before their enrollment; parents or legal guardians signed the informed consent for underage participants. The ethical committee of the Institute for Maternal and Child Health – IRCCS "Burlo Garofolo" approved the study under the univocal code Prot. CE/V-78, 06/08/2007. All study procedures were performed under the ethical principles expressed by the Helsinki Declaration.

**INGI-VBI (INGI-Val Borbera)**: is a collection of samples from seven small villages in the Borbera Valley, a geographically isolated valley located within the Apennine Mountains, in the southern part of Piedmont, North-Western of Italy. These populations belong to the Italian Network of Genetic Isolates (INGI), a project that aims to study the genetic basis of complex diseases and phenotypes, taking advantage of these populations' genetic and environmental homogeneity (doi: 10.1038/s41431-019-0551-x). Individuals with an age range from 18 to 87 years old were voluntarily enrolled. Demographic data and detailed familial and individual medical history were collected for all the participants. In addition, participants underwent specialist evaluations, e.g., neurological, psychiatric, in-depth sensorial, etc. Genotyping and phenotypic data for 1785 samples are available. Written informed consent for participation was obtained from all subjects. The project was approved by the Ethical committee of the San Raffaele Hospital (Milan, Italy). All study procedures were performed under the ethical principles expressed by the Helsinki Declaration.

**IRAS Family Study (Insulin Resistance Atherosclerosis Study):** The IRASFS was a family study designed to examine the genetic and epidemiologic basis of glucose homeostasis traits and abdominal adiposity. Briefly, self-reported Mexican American pedigrees were recruited in San Antonio, TX and San Luis Valley, CO. Probands with large families were recruited from the initial non-family-based IRAS, which was modestly enriched for impaired glucose tolerance and T2D. Inclusion of IRASFS data is limited to 1040 normoglycemic individuals in 88 pedigrees with genotype data from the Illumina OmniExpress and Omni 1S arrays and imputation to the 1000 Genome Integrated Reference Panel (phase I).

**JHS (Jackson Heart Study):** The JHS is a longitudinal, community-based observational cohort study investigating the role of environmental and genetic factors in the development of cardiovascular disease in African Americans. Between 2000 and 2004, a total of 5301 participants were recruited from a tri-county area (Hinds, Madison, and Rankin Counties) that encompasses Jackson, MS. Details of the design and recruitment for the Jackson Heart Study cohort has been previously published [1-3]. Briefly, approximately 30% of participants were former members of the Atherosclerosis Risk in Communities (ARIC) study. The remainder were recruited by either 1) random selection from the Accudata list, 2) commercial listing, 3) a constrained volunteer sample, in which recruitment was distributed among defined demographic cells in proportions designed to mirror those in the overall population, or through the Jackson Heart Study Family Study.

1. Wyatt SB, Diekelmann N, Henderson F, Andrew ME, Billingsley G, Felder SH, et al. A community-driven model of research participation: the Jackson Heart Study Participant Recruitment and Retention Study. *Ethn Dis*. 2003;13(4):438-55. PubMed PMID: 14632263.

2. Taylor HA, Jr., Wilson JG, Jones DW, Sarpong DF, Srinivasan A, Garrison RJ, et al. Toward resolution of cardiovascular health disparities in African Americans: design and methods of the Jackson Heart Study. *Ethn Dis*. 2005;15(4 Suppl 6):S6-4-17. PubMed PMID: 16320381.

3. Fuqua SR, Wyatt SB, Andrew ME, Sarpong DF, Henderson FR, Cunningham MF, et al. Recruiting African-American research participation in the Jackson Heart Study: methods, response rates, and sample description. *Ethn Dis*. 2005;15(4 Suppl 6):S6-18-29. PubMed PMID: 16317982.

**JUPITER** (Justification for the Use of Statins in Primary Prevention: An Intervention Trial Evaluating Rosuvastatin): Genetic analysis was performed in a sub-population from JUPITER (Justification for the Use of statins in Prevention: an Intervention Trial Evaluating Rosuvastatin), an international, randomized, placebo-controlled trial of rosuvastatin (20mg/day) in the primary prevention of cardiovascular disease conducted among apparently healthy men and women with LDL-C < 130 mg/dL and hsCRP ≥ 2 mg/L (1, 2). Individuals with diabetes or triglyceride concentration >500mg/dL were excluded. The present analysis includes only individuals who provided consent for genetic analysis, had successfully collected genotype information, and who had either verified European or verified South African black ancestry. Both JUPITER and the collection of genetic data in the JUPITER study population were funded by AstraZeneca.

1. Ridker PM, Danielson E, Fonseca FA, Genest J, Gotto AM Jr, Kastelein JJ, Koenig W, Libby P, Lorenzatti AJ, MacFadyen JG, Nordestgaard BG, Shepherd J, Willerson JT, Glynn RJ; JUPITER Study Group. Rosuvastatin to prevent vascular events in men and women with elevated C-reactive protein. N Engl J Med 2008 Nov 20; 359(21):2195-207

2. Chasman DI, Giulianini F, MacFadyen J, Barratt BJ, Nyberg F, Ridker PM. Genetic determinants of statin-induced low-density lipoprotein cholesterol reduction: the Justification for the Use of Statins in Prevention: an Intervention Trial Evaluating Rosuvastatin (JUPITER) trial. Circ Cardiovasc Genet. 2012 Apr 1;5(2):257-64. doi: 10.1161/CIRCGENETICS.111.961144. Epub 2012 Feb 13.

**KoGES (Korean Genome and Epidemiology Study)**: KoGES is a large-scale prospective cohort study with a comprehensive range of phenotypic measures and biological samples collected on approximately 210,000 individuals. KoGES's long-term objective is to develop comprehensive and applicable healthcare guidelines for common complex diseases in Koreans, reduce the burden of chronic diseases and improve the quality of life. KoGES includes population-based cohorts, the community-based Ansan and Ansung study, the urban community-based health examinee study, and the rural community-based cardiovascular disease association study. The cohorts consist of community-dwellers and participants recruited from the national health examinee registry, men and women, aged ≥ 40 years at baseline. A total of 72,000 samples were genotyped with KoreanChip, a customized array optimized for the Korean population, and imputed using IMPUTE4 with 1000 Genomes Project Phase 3 data and the Korean reference genome as a reference panel. Measures of the baseline recruitment, and only genotyped samples that met the following exclusion criteria were used in the analyses: low call rate (<97%), excessive heterozygosity, excessive singletons, gender discrepancy, and cryptic first-degree relatives. SNPs with low HWE p value ($<10^{-6}$) or low call rate (<95%) were excluded. Variants with imputation quality score (IQS) < 0.8 and MAF <1% were excluded after imputation.


1. Kim, Yeonjung, Bok-Ghee Han, and KoGES Group. "Cohort profile: the Korean genome and epidemiology study (KoGES) consortium." International journal of epidemiology 46.2 (2017): e20-e20.
2. Moon, Sanghoon, et al. "The Korea Biobank Array: design and identification of coding variants associated with blood biochemical traits." Scientific reports 9.1 (2019): 1-11.
3. Nam, Kisung, Jangho Kim, and Seunggeun Lee. "Genome-wide study on 72,298 individuals in Korean biobank data for 76 traits." Cell Genomics 2.10 (2022): 100189.


**LBC1921 (Lothian Birth Cohort 1921)**: LBC1921 consists of 550 relatively healthy individuals, most of whom took part in the Scottish Mental Survey of 1932 at the age of ~11 years old. They were recruited to a study to determine influences on cognitive ageing at age ~79 years, when almost all lived independently in the Lothian region of Scotland. They have taken part in five waves of testing in later life (at mean ages 79, 83, 87, 90 and 85 years). At each wave they underwent a series of cognitive and physical tests.[1,2]


1. Deary IJ, Gow AJ, Pattie A, Starr JM. Cohort profile: the Lothian Birth Cohorts of 1921 and 1936. Int J Epidemiol 2012;41:1576-1584.

2. Taylor AM, Pattie A, Deary IJ. Cohort Profile Update: The Lothian Birth Cohorts of 1921 and 1936. Int J Epidemiol 2018;47:1042-1042r


**LBC1936 (Lothian Birth Cohort 1936):** LBC1936 consists of 1091 relatively healthy individuals, most of whom took part in the Scottish Mental Survey of 1947 at the age of ~11 years old. They were recruited to a study to determine influences on cognitive ageing at age ~70 years, when almost all lived independently in the Lothian region of Scotland. They have taken part in six waves of testing in later life (at mean ages 70, 73, 76, 79, 82 and 85 years). At each wave they underwent a series of cognitive and physical tests.[1,2]

1. Deary IJ, Gow AJ, Pattie A, Starr JM. Cohort profile: the Lothian Birth Cohorts of 1921 and 1936. Int J Epidemiol 2012;41:1576-1584.

2. Taylor AM, Pattie A, Deary IJ. Cohort Profile Update: The Lothian Birth Cohorts of 1921 and 1936. Int J Epidemiol 2018;47:1042-1042r

**Lifelines**: (https://lifelines.nl/) Lifelines is a multi-disciplinary prospective population-based cohort study using a unique three-generation design to examine the health and health-related behaviors of 165,000 persons living in the North East region of The Netherlands. It employs a broad range of investigative procedures in assessing the biomedical, socio-demographic, behavioral, physical and psychological factors which contribute to the health and disease of the general population, with a special focus on multimorbidity. In addition, the Lifelines project comprises a number of cross-sectional sub-studies, which investigate specific age-related conditions. These include investigations into metabolic and hormonal diseases, including obesity, cardiovascular and renal diseases, pulmonary diseases and allergy, cognitive function and depression, and musculoskeletal conditions. Recruitment has been going on since the end of 2006, and over 130,000 participants had been included by April 2013. At the baseline examination, the participants in the study were asked to fill in a questionnaire before the first visit. During the first and second visit, the first or second part of the questionnaire, respectively, are checked for completeness, a number of investigations are conducted, and blood and urine samples are taken. Lifelines is a facility that is open for all researchers. Information on application and data access procedure is summarized on https://www.lifelines-biobank.com/ (Scholtens S, Smidt N, Swertz MA, Bakker SJ, Dotinga A, Vonk JM, et al. Cohort Profile: LifeLines, a three-generation cohort study and biobank. Int J Epidemiol. 2014 Dec 14.)

Lifelines was genotyped in 2 stages: the first stage consisted of ~15,000 participants and used the Illumina CytoSNP chip; the second stage included ~35,000 participants and used the Illumina GSA chip. Due to the differences in genotyping, these two groups are analysed separately. Samples that are present, or have close relatives in both groups, are excluded from the CytoSNP dataset prior to analysis.

**Living Biobank – for contributing data LivingBiobank-CHS and LivingBiobank-MAS**

Living Biobank is a collection of population-based Chinese and Malay individuals from two multi-ethnic cohorts from Singapore Population Health Studies: Multi Ethnic Cohort Phase 1 (MEC1) and the Singapore Health 2012 (SH2012) that aims to investigate the genetic and lifestyle factors that affect the risk of developing chronic diseases such as T2D, cardiovascular outcomes, and cancer in Singapore citizens and permanent residents [1]. MEC1 was a population-based cohort initiated in 2007 to investigate the genetic and lifestyle factors that affect the risk of developing chronic diseases, such as diabetes and cardiovascular outcomes, in the 3 ethnic groups (Chinese, Malay, and Indians) [2]. SH2012 was a population-based, cross-sectional survey conducted between 2012 and 2013 in the 3 ethnic groups (Chinese, Malay, and Indian) with oversampling in the Malay and Indian populations [3]. Two readings of blood pressure were taken from participants after 5 min of rest, seated, using an automated blood pressure monitor (Dinamap Pro100V2; Criticon, Norderstedt, Germany) by trained observers. One of two cuff sizes (regular, large) was chosen on the basis of the circumference of the participant's arm. A third reading was performed if the difference between two readings of either the systolic blood pressure was greater than 10mmHg or the diastolic blood pressure was greater than 5mmHg. The mean values of the closest two readings were used.

1. Chai JF, Kao SL, Wang C, Lim VJ, Khor IW, Dou J, Podgornaia AI, Chothani S, Cheng CY, Sabanayagam C, Wong TY, van Dam RM, Liu J, Reilly DF, Paterson AD, Sim X. Genome-Wide Association for HbA1c in Malay Identified Deletion on SLC4A1 that Influences HbA1c Independent of Glycemia. J Clin Endocrinol Metab. 2020 Dec 1;105(12):dgaa658. doi: 10.1210/clinem/dgaa658. PMID: 32936915.

2. Tan KHX, Tan LWL, Sim X, Tai ES, Lee JJ, Chia KS, van Dam RM. Cohort Profile: The Singapore Multi-Ethnic Cohort (MEC) study. Int J Epidemiol. 2018 Jun 1;47(3):699-699j. doi: 10.1093/ije/dyy014. PMID: 29452397.

3. Win, A.M., Yen, L.W., Tan, K.H. et al. Patterns of physical activity and sedentary behavior in a representative sample of a multi-ethnic South-East Asian population: a cross-sectional study. BMC Public Health 15, 318 (2015). https://doi.org/10.1186/s12889-015-1668-7

**LLFS (The Long Life Family Study):** LLFS is a longitudinal, population-based multigenerational family cohort designed to study genetic, behavioral, and environmental factors in families exhibiting exceptional longevity. Families were sampled from four clinical centers: Boston University Medical Center in Boston, MA; Columbia College of Physicians and Surgeons in New York City, NY; the University of Pittsburgh in Pittsburgh, PA, USA; and the University of Southern Denmark, Denmark. The characteristics, recruitment, eligibility, and enrollment were previously described (PMID: 21258136, PMID: 34739053). The first clinical exam started in 2006 and recruited 4,953 individuals in 539 two-generational families that demonstrated clustering for exceptional survival in the upper generation. The second clinical exam (2014-2017) revisited 2,933 European descent individuals from 528 families. The third clinical exam (2021-) is recruiting the participants from second exam and a few new ones from the grandchild generation. The individuals were genotyped using ~2.3 million SNPs from the Illumina Omni chip, then imputed on Version R2 of the TOPMed reference panel using the Michigan Imputation Server (https://imputation.biodatacatalyst.nhlbi.nih.gov/#!), which used Eagle v2.4 for phasing and minimac4 v1.3.3 for imputation.

**MESA (Multi-Ethnic Study of Atherosclerosis)** The Multi-Ethnic Study of Atherosclerosis (MESA) is a study of the characteristics of subclinical cardiovascular disease and the risk factors that predict progression to clinically overt cardiovascular disease or progression of the subclinical disease. MESA consisted of a diverse, population-based sample of an initial 6,814 asymptomatic men and women aged 45-84. 38 percent of the recruited participants self described as White, 28 percent African American, 22 percent Hispanic, and 12 percent Chinese. Participants were recruited from six field centers across the United States: Wake Forest University, Columbia University, Johns Hopkins University, University of Minnesota, Northwestern University and University of California - Los Angeles. Participants are being followed for identification and characterization of cardiovascular disease events, including acute myocardial infarction and other forms of coronary heart disease (CHD), stroke, and congestive heart failure; for cardiovascular disease interventions; and for mortality. The first examination took place over two years, from July 2000 - July 2002. It was followed by five examination periods that were 17-20 months in length, including the recently completed Exam 7 (2022-2024). Participants have been contacted every 9 to 12 months throughout the study to assess clinical morbidity and mortality. Informed consent was obtained for extensive data sharing (dbGaP) and genetic/omic studies, including candidate genes (NHLBI CARe), genome-wide scans (NHLBI SHARe), exome sequencing (NHBLI ESP) and, most recently, the NHLBI TOPMed program.

1. Bild DE, Bluemke DA, Burke GL, Detrano R, Diez Roux AV, Folsom AR, Greenland P, Jacob DR Jr, Kronmal R, Liu K, Nelson JC, O'Leary D, Saad MF, Shea S, Szklo M, Tracy RP. Multi-ethnic study of atherosclerosis: objectives and design. Am J Epidemiol. 2002 Nov 1;156(9):871-81. PubMed PMID: 12397006.

**METSIM:** The participants of our study were selected from the METSIM study comprising 10197 Finnish men randomly selected from the population register of Kuopio, Eastern Finland, aged from 45 to 73 years at baseline. The study was approved by the Ethics Committee of the Kuopio University Hospital. All study participants gave written informed consent.

**Million Veteran Program (MVP):** The MVP is a mega-biobank that was launched in 2011 and supported entirely by the Veterans Health Administration Office of Research and Development in the United States. The MVP received ethical and study protocol approval from the VA Central Institutional Review Board (IRB) in accordance with the principles outlined in the Declaration of Helsinki. The specific design, initial demographics and quality-control procedures of the MVP have been detailed previously (10.1016/j.jclinepi.2015.09.016).

**MONA LISA Lille (Monitoring National du Risque Artériel; National Monitoring of Arterial Risk)**

The MONA LISA (Monitoring National du Risque Artériel; National Monitoring of Arterial Risk) Lille study is a population-based, cross-sectional study of a representative sample of 1578 participants recruited from within the Lille urban area in northern France performed between 2004 and 2006. Inhabitants (aged 35–74) were randomly sampled from electoral rolls. The cities/towns/villages were randomly selected (by the numbers of inhabitants) to be representative of the background population distribution. The goal was to obtain 200 participants for each gender and each ten-year age class. The main objective of the study were to study the prevalence of cardiovascular risk factors. The protocol was approved by the appropriate independent ethics committee. Written, informed consent to participation was provided by all participants.

**NEO (The Netherlands Epidemiology of Obesity study):** The NEO was designed for extensive phenotyping to investigate pathways that lead to obesity-related diseases. The NEO study is a population-based, prospective cohort study that includes 6,671 individuals aged 45–65 years, with an oversampling of individuals with overweight or obesity. At baseline, information on demography, lifestyle, and medical history have been collected by questionnaires. In addition, samples of 24-h urine, fasting and postprandial blood plasma and serum, and DNA were collected. Genotyping was performed using the Illumina HumanCoreExome chip, which was subsequently imputed to the 1000 genome reference panel. Participants underwent an extensive physical examination, including anthropometry, electrocardiography, spirometry, and measurement of the carotid artery intima-media thickness by ultrasonography. In random subsamples of participants, magnetic resonance imaging of abdominal fat, pulse wave velocity of the aorta, heart, and brain, magnetic resonance spectroscopy of the liver, indirect calorimetry, dual energy X-ray absorptiometry, or accelerometry measurements were performed. The collection of data started in September 2008 and completed at the end of September 2012. Participants are currently being followed for the incidence of obesity-related diseases and mortality.

**NESDA:** The Netherlands Study of Depression and Anxiety (NESDA) is an ongoing longitudinal cohort study to examine the prevalence, long-term course and consequences of depressive and anxiety disorders in the adult population. A detailed description of the study can be found elsewhere[1]. Briefly, a total of 2981 participants, consisting of a healthy control group, people with a history of depressive or anxiety disorder and people with current depressive and/or anxiety disorder, were recruited from community (19%), primary care (54%), and outpatient psychiatric clinics (27%), and included at the baseline assessment in 2004-2007. Inclusion criteria were a lifetime diagnosis of major depressive disorder or anxiety disorder, age 18-65 years, and self-reported western European ancestry. Excluded were those who were not fluent in Dutch, and those with a primary diagnosis of psychotic disorder, obsessive compulsive disorder, bipolar disorder, or severe substance use or dependence. Biological sample collection and biobanking procedures (i.e. blood sampling and DNA isolation) took place during the baseline visit and has been previously described in detail [2].

NESDA samples were genotyped using either the Perlegen-Affymetrix 5.0, or Affymetrix 6.0 genotyping chip and called with BirdSeed. SNPs were excluded if unmapped or mapped to multiple locations, call rate<95%, MAF<0.01, HWE p-value<$10^{-5}$, unmapped to build 36, allele frequency difference with 1000Genomes reference >20%, or palindromic SNPs with allele frequency>35%. Samples were removed if call rate<90%, deviant heterozygosity, abs(PLINK F)>0.1, sex mismatch, unexpected relatedness, or non-Caucasian. Imputation was performed with Impute software using the 1000Genomes Phase 1 Integrated Release 3 ALL reference panel.

1. Penninx BW, Beekman AT, Smit JH, et al. The netherlands study of depression and anxiety (NESDA): Rationale, objectives and methods. *International journal of methods in psychiatric research*. 2008;17(3):121-140.

2. Boomsma DI, Willemsen G, Sullivan PF, et al. Genome-wide association of major depression: Description of samples for the GAIN major depressive disorder study: NTR and NESDA biobank projects. *European Journal of Human Genetics*. 2008;16(3):335.

**PROCARDIS:** The **Precocious Coronary Artery Disease Study (PROCARDIS)** consists of coronary artery disease (CAD) cases and controls from four European countries (UK, Italy, Sweden and Germany). CAD (defined as myocardial infarction, acute coronary syndrome, unstable or stable angina, or need for coronary artery bypass surgery or percutaneous coronary intervention) was diagnosed before 66 years of age and 80% of cases had a sibling fulfilling the same criteria for CAD. Subjects with self-reported non-European ancestry were excluded. Among the “genetically-enriched” CAD cases, 70% had suffered myocardial infarction (MI).

PROCARDIS was genotyped using Illumina Human 1M and 610K quad arrays on a total of 6000 patients with CAD and 7,500 control subjects. Genotype quality control excluded SNPs Call rate ≥ 95% for SNPs with MAF ≥ 1%, and call rate ≥ 99% for SNPs with MAF < 1%. Samples with high heterozygosity, with sample call rate <95% or with sex inconsistencies were removed. After quality control, genotypes were imputed to TOPMED freeze 5 reference panel.

**PROSPER:** All data come from the PROspective Study of Pravastatin in the Elderly at Risk (PROSPER). A detailed description of the study has been published elsewhere (1-3). PROSPER was a prospective multicenter randomized placebo-controlled trial to assess whether treatment with pravastatin diminishes the risk of major vascular events in elderly. Between December 1997 and May 1999, we screened and enrolled subjects in Scotland (Glasgow), Ireland (Cork), and the Netherlands (Leiden). Men and women aged 70-82 years were recruited if they had pre-existing vascular disease or increased risk of such disease because of smoking, hypertension, or diabetes. A total number of 5,804 subjects were randomly assigned to pravastatin or placebo. A large number of prospective tests were performed including Biobank tests and cognitive function measurements. A whole genome wide screening has been performed in the sequential PHASE project. Of 5,763 subjects DNA was available for genotyping. Genotyping was performed with the Illumina 660K beadchip, after QC (call rate <95%) 5,244 subjects and 557,192 SNPs were left for analysis. These SNPs were imputed to 2.5 million SNPs based on the HAPMAP built 36 with MACH imputation software. The study was approved by the institutional ethics review boards of centers of Cork University (Ireland), Glasgow University (Scotland) and Leiden University Medical Center (the Netherlands) and all participants gave written informed consent.

**RHS (Ragama Health Study):** The Ragama Health Study (RHS) is a population-based study of South Asian men and women aged 35-64 yrs living in the Ragama Medical Officer of Health (MOH) area, near Colombo, Sri Lanka.* Consenting adults attended a clinic after a 12-h fast with available health records, and were interviewed by trained personnel to obtain information on medical, sociodemographic, and lifestyle variables. A 10-mL sample of venous blood was obtained from each subject. The concurrent study was performed in two tea plantation estates in the Lindula MOH area, near Nuwara Eliya (180 km from Colombo), to investigate the gene-environment interaction in a community with differing lifestyles (e.g., physical activity and diet). The RHS is a collaborative effort between the Faculty of Medicine, University of Kelaniya and the National Center for Global Health and Medicine, Japan.

*Dassanayake, A.S. *et al.* Prevalence and risk factors for non-alcoholic fatty liver disease among adults in an urban Sri Lankan population. *J Gastroenterol Hepatol* **24**, 1284-8 (2009).

**SEED (Singapore Epidemiology of Eye Diseases) – for contributing data SCES-610, SCES-Omni, SiMES, and SINDI**

**SCES (The Singapore Chinese Eye Study): SCES** is a population-based, cross-sectional study of Chinese adults residing in the southwestern part of Singapore, with a focus on studying major eye diseases, including diabetic retinopathy, age-related macular degeneration, glaucoma, refractive errors and cataract. The Ministry of Home Affairs provided an initial computer generated list ethnic Chinese names of adults aged 40–80+ years of age. A final sampling frame of 6350 ethnic Chinese residents was derived from this list using an age-stratified random sampling strategy similar to SiMES and SINDI.

1. Lavanya R, Jeganathan VS, Zheng Y, Raju P, Cheung N, Tai ES, Wang JJ, Lamoureux E, Mitchell P, Young TL, Cajucom-Uy H, Foster PJ, Aung T, Saw SM, Wong TY. Methodology of the Singapore Indian Chinese Cohort (SICC) eye study: quantifying ethnic variations in the epidemiology of eye diseases in Asians. Ophthalmic Epidemiol. 2009 Nov-Dec;16(6):325-36. doi: 10.3109/09286580903144738. PubMed PMID: 19995197.

**SiMES (Singapore Malay Eye Study):** SiMES is a population-based, cross-sectional study of 3280 Malay adults aged 40–79 years, with a focus on studying major eye diseases, including diabetic retinopathy, age-related macular degeneration, glaucoma, refractive errors and cataract. An age-stratified random sampling of all Malay adults, aged 40–80 years, residing in 15 residential districts in the southwestern part of Singapore was drawn from the computer-generated random list of 16069 Malay names provided by the Ministry of Home Affairs. Of the 4168 eligible individuals, 3280 participants (78.7%) took part in the study. The study was conducted from August 2004 to June 2006.

1. Foong AW, Saw SM, Loo JL, Shen S, Loon SC, Rosman M, Aung T, Tan DT, Tai ES, Wong TY. Rationale and methodology for a population-based study of eye diseases in Malay people: The Singapore Malay eye study (SiMES). Ophthalmic Epidemiol. 2007 Jan-Feb;14(1):25-35. PubMed PMID: 17365815.

**SINDI (Singapore Indian Eye Study):** SINDI is a population-based, cross-sectional epidemiological study of ethnic Indian adults aged between 40 and 80+ years residing in Singapore, with a focus on studying major eye diseases, including diabetic retinopathy, age-related macular degeneration, glaucoma, refractive errors and cataract. The Ministry of Home Affairs provided an initial computer-generated list of Indian names derived from a simple random sampling of all ethnic Indian adults aged 40–80+ years of age residing in 15 residential districts in southwestern Singapore. From this list, a final sampling frame of 6350 ethnic Indian residents was derived using an age-stratified random sampling strategy similar to SiMES. SINDI was conducted from March 2007 to December 2009 and recruited 3400 (75% response rate) participants.

1. Lavanya R, Jeganathan VS, Zheng Y, Raju P, Cheung N, Tai ES, Wang JJ, Lamoureux E, Mitchell P, Young TL, Cajucom-Uy H, Foster PJ, Aung T, Saw SM, Wong TY. Methodology of the Singapore Indian Chinese Cohort (SICC) eye study: quantifying ethnic variations in the epidemiology of eye diseases in Asians. Ophthalmic Epidemiol. 2009 Nov-Dec;16(6):325-36. doi: 10.3109/09286580903144738. PubMed PMID: 19995197.

**SHIP (Study of Health in Pomerania):** The Study of Health In Pomerania (SHIP) is a prospective longitudinal population-based cohort study in Mecklenburg-Western Pomerania assessing the prevalence and incidence of common diseases and their risk factors (PMID: 20167617 and 35348705). SHIP encompasses the two independent cohorts SHIP-START and SHIP-TREND. Participants aged 20 to 79 with German citizenship and principal residency in the study area were recruited from a random sample of residents living in the three local cities, 12 towns as well as 17 randomly selected smaller towns. Individuals were randomly selected stratified by age and sex in proportion to population size of the city, town or small towns, respectively. A total of 4,308 participants were recruited between 1997 and 2001 in the SHIP-START cohort. Between 2008 and 2012 a total of 4,420 participants were recruited in the SHIP-TREND cohort. Individuals were invited to the SHIP study center for a computer-assisted personal interviews and extensive physical examinations. The study protocol was approved by the medical ethics committee of the University of Greifswald. Oral and written informed consent was obtained from each of the study participants.
Genome-wide SNP-typing was performed using the Affymetrix Genome-Wide Human SNP Array 6.0 (SHIP-START samples), the Illumina Infinium HumanOmni2.5 BeadChip, or the Illumina Infinium Global Screening Array (SHIP-TREND samples). Array processing was carried out in accordance with the manufacturer's standard recommendations. Genotypes were determined using the Birdseed2 clustering algorithm for SHIP-START, GenomeStudio Genotyping Module v1.0, and GenomeStudio 2.0 Genotyping Module (GenCall) for SHIP-TREND.

**Singapore Prospective Study Program (SP2) – for contributing data SP2-1M and SP2-610**
SP2 (Singapore Prospective Study Program): The SP2 is a population-based study of diabetes and cardiovascular disease in Singapore. It first surveyed subjects (Chinese, Malay and Indian) from four cross-sectional studies that were conducted in Singapore between 1982 and 1998. Subjects were between the ages of 24-95 years and represented a random sample of the Singapore population. Subjects were re-visited between 2003 and 2007. Among the 10,747 individuals who were eligible, 5,157 subjects completed a questionnaire and the subsequent clinical examinations. Of the 5,517 subjects, 2,098 Chinese were genotyped on a combination of Illumina 610, 1M arrays. Two readings of blood pressure were taken from participants after 5 min of rest, seated, using an automated blood pressure monitor (Dinamap Pro100V2; Criticon,

Norderstedt, Germany) by trained observers. One of two cuff sizes (regular, large) was chosen on the basis of the circumference of the participant's arm. A third reading was performed if the difference between two readings of either the systolic blood pressure was greater than 10mmHg or the diastolic blood pressure was greater than 5mmHg. The mean values of the closest two readings were used.

1. Nang EE, Khoo CM, Tai ES, Lim SC, Tavintharan S, Wong TY, Heng D, Lee J. Is there a clear threshold for fasting plasma glucose that differentiates between those with and without neuropathy and chronic kidney disease?: the Singapore Prospective Study Program. Am J Epidemiol. 2009 Jun 15;169(12):1454-62. doi: 10.1093/aje/kwp076. Epub 2009 Apr 30. PMID: 19406920.
2. Tan KHX, Tan LWL, Sim X, Tai ES, Lee JJ, Chia KS, van Dam RM. Cohort Profile: The Singapore Multi-Ethnic Cohort (MEC) study. Int J Epidemiol. 2018 Jun 1;47(3):699-699j. doi: 10.1093/ije/dyy014. PMID: 29452397.

**SWAN (The Study of Women's Health Across the Nation):**

The **S**tudy of **W**omen's Health **A**cross the **N**ation (SWAN) is a multi-site, multiracial/ethnic longitudinal study of women's health designed to describe the biological, behavioral, and psychosocial characteristics that occur during midlife and the menopausal transition. In addition to characterizing reproductive aging, SWAN focuses on the impact of menopause on age-related chronic diseases, such as diabetes, cardiovascular disease, depression, bone loss and osteoporosis, as well as physical and cognitive functioning. The SWAN cohort was enrolled in 1996-97 and consists of 3302 community-based women from seven sites with data from five race/ethnic groups: Black (n=935), Chinese (n=250), Hispanic (n=286), Japanese (n=281), and White (n=1550). To be eligible for enrollment women had to be aged 42 to 52 years old, have an intact uterus and at least one ovary, have had a menstrual period in the previous three months, and not be taking hormones. SWAN participants have completed 15 approximately annual follow-up visits since enrollment. During follow-up visits 5 and 6 (2001-2003), 1757 women from six sites were consented to provide genetic materials for whom extracted DNA was successfully obtained for 1536 women: Black (n=410), Chinese (n=151), Hispanic (n=0), Japanese (n=168), and White (n=807). This analysis was done separately for Whites and Blacks.

Sowers MF, Crawford S, Sternfeld B, Morganstein DG, Gold EB, Greendale G, Evans D, Neer R, Matthews K, Sherman S, Lo A, Weiss G, Kelsey J. SWAN: A multi-center, multi-ethnic, community-based cohort study of women and the menopausal transition. In: Lobo RA, Kelsey JL, Marcus R, editors. Menopause: biology and pathobiology. San Diego, CA: Academic Press; 2000. p. 175-88

**UKBB (UK Biobank):** UK Biobank (UKB, www.ukbiobank.ac.uk) is a large longitudinal biobank study in the United Kingdom which was established to improve understanding of the genetic and environmental causes of common diseases including cardiovascular diseases. In addition to self-reported disease outcomes and extensive health and life-style questionnaire data, UKB participants are being tracked through their NHS records and national registries (including cause of death and Hospital Episode Statistics). In 2017, UKB released the genotypes of 488,377 participants profiled with a custom SNP array. Genotyping QC was performed centrally by UKB, and genotypes imputed to Haplotype Reference Consortium (HRC) panel were released for 488,377 participants. Sample selection for the Gene-Lifestyle Interaction projects was based on available datasets for traits and lifestyle measures.

**WGHS** (Women's Genome Health Study): WGHS is a prospective cohort of female North American health care professionals representing participants in the Women's Health Study (WHS) trial who provided a blood sample at baseline and consent for blood-based analyses (1). Participants in the WHS were 45 years or older at enrollment and free of cardiovascular disease, cancer or other major chronic illness. The current data are derived from 23,294 WGHS participants for whom whole genome genotype information was available at the time of analysis and for whom self-reported European ancestry could be confirmed by multidimensional scaling analysis of 1,443 ancestry informative markers in PLINK v. 1.06. At baseline, BP and lifestyle habits related to smoking, consumption of alcohol, and physical activity as well as other general clinical information were ascertained by a self-reported questionnaire, an approach that has been validated in the WGHS demographic, namely female health care professionals. The WGHS is supported by the National Heart, Lung, and Blood Institute (HL043851 and HL080467) and the National Cancer Institute (CA047988 and UM1CA182913), with support for genotyping provided by Amgen.

1. Ridker PM, Chasman DI, Zee RY, Parker A, Rose L, Cook NR, Buring JE. Rationale, design, and methodology of the Women's Genome Health Study: a genome-wide association study of more than 25,000 initially healthy american women. Clin Chem. 2008;54(2):249-55. PubMed PMID: 18070814.

**WHI (Women's Health Initiative)**: is a long-term national health study that focuses on strategies for preventing common diseases such as heart disease, cancer and fracture in postmenopausal women. A total of 161,838 women aged 50–79 years old were recruited from 40 clinical centers in the US between 1993 and 1998(1, 2). WHI consists of an observational study, two clinical trials of postmenopausal hormone therapy (HT, estrogen alone or estrogen plus progestin), a calcium and vitamin D supplement trial, and a dietary modification trial. Study recruitment and exclusion criteria have been described previously(1, 2). Recruitment was done through mass mailing to age-eligible women obtained from voter registration, driver's license and Health Care Financing Administration or other insurance list, with emphasis on recruitment of minorities and older women. Exclusions included participation in other randomized trials, predicted survival < 3 years, alcoholism, drug dependency, mental illness and dementia. For the CT, women were ineligible if they had a systolic BP > 200 mm Hg or diastolic BP > 105 mm Hg, a history of hypertriglyceridemia or breast cancer. Study protocols and consent forms were approved by the IRB at all participating institutions. Socio-demographic characteristics, lifestyle, medical history and self-reported medications were collected using standardized questionnaires at the screening visit.

Genome wide association study (GWAS) non-overlapping samples are composed of (a) a case-control study (WHI Genomics and Randomized Trials Network – GARNET, which included all coronary heart disease, stroke, venous thromboembolic events and selected diabetes cases that happened during the active intervention phase in the WHI HT clinical trials and aged matched controls), (b) women selected to be "representative" of the HT trial (mostly younger white HT subjects that were also enrolled in the WHI memory study - WHIMS) and (c) the WHI SNP Health Association Resource (WHI SHARe), a randomly selected sample of 8,515 African American and 3,642 Hispanic women from WHI. Genotyping was performed using Affymetrix 6.0 (WHI-SHARe), HumanOmniExpressExome-8v1_B (WHIMS) and Illumina HumanOmni1-Quad v1-0 B (GARNET). Quality control of the GWAS data included variant and sample call rates >95%, and included identification of genetically related individuals. Principal components were computed using methods developed by Price et al(3). Imputation was performed using the TOPMed Imputation Server and freeze 8 reference multi-ethnic panel (build hg38). Due to some overlap of WHI participants with the TOPMed reference panel, we re-calculated the estimated imputation quality based on only the samples not on TOPMed reference panel, to account for the over-estimation of imputation quality given by the imputation software(4). Variants with an imputation quality (Rsq) <0.3 were filtering out. After QC and exclusions from analysis protocol, the number of women included in analysis is 4,423 whites for GARNET, 5,202 white for WHIMS, 7,919 for SHARe African American and 3,377 for SHARe Hispanics. Analyses were performed using LinGxEScanR software.


1. Design of the Women's Health Initiative clinical trial and observational study. The Women's Health Initiative Study Group. Control Clin Trials. 1998;19(1):61-109. Epub 1998/03/11. doi: S0197245697000780 [pii]. PubMed PMID: 9492970.
2. Anderson GL, Manson J, Wallace R, Lund B, Hall D, Davis S, Shumaker S, Wang CY, Stein E, Prentice RL. Implementation of the Women's Health Initiative study design. Ann Epidemiol. 2003;13(9 Suppl):S5-17. Epub 2003/10/25. PubMed PMID: 14575938.
3. Price AL, Patterson NJ, Plenge RM, Weinblatt ME, Shadick NA, Reich D. Principal components analysis corrects for stratification in genome-wide association studies. Nat Genet. 2006;38(8):904-9. doi: 10.1038/ng1847. PubMed PMID: 16862161.
4. Sun Q, Liu W, Rosen JD, Huang L, Pace RG, Dang H, Gallins PJ, Blue EE, Ling H, Corvol H, Strug LJ, Bamshad MJ, Gibson RL, Pugh EW, Blackman SM, Cutting GR, O'Neal WK, Zhou YH, Wright FA, Knowles MR, Wen J, Li Y, Cystic Fibrosis Genome P. Leveraging TOPMed imputation server and constructing a cohort-specific imputation reference panel to enhance genotype imputation among cystic fibrosis patients. HGG Adv. 2022;3(2):100090. doi: 10.1016/j.xhgg.2022.100090. PubMed PMID: 35128485; PMCID: PMC8804187.


**YFS (The Cardiovascular Risk in Young Finns Study):** The YFS is a population-based follow up-study started in 1980. The main aim of the YFS is to determine the contribution made by childhood lifestyle, biological and psychological measures to the risk of cardiovascular diseases in adulthood. In 1980, over 3,500 children and adolescents all around Finland participated in the baseline study. The follow-up studies have been conducted mainly with 3-year intervals. The latest 30-year follow-up study was conducted in 2010-11 (ages 33-49 years) with 2,063 participants. The study was approved by the local ethics committees (University Hospitals of Helsinki, Turku, Tampere, Kuopio and Oulu) and was conducted following the guidelines of the Declaration of Helsinki. All participants gave their written informed consent.

### 2. Study Acknowledgments

**AGES (Age Gene/Environment Susceptibility Reykjavik Study)**: The AGES-Reykjavik study was financed by National Institute on Aging (NIA) contracts N01-AG-12100 and HHSN271201200022C for Vi.G. and Althingi (the Icelandic Parliament).

**ARIC (Atherosclerosis Risk in Communities) Study:** The ARIC study has been funded in whole or in part with Federal funds from the National Heart, Lung, and Blood Institute, National Institutes of Health, Department of Health and Human Services, under Contract nos. (75N92022D00001, 75N92022D00002, 75N92022D00003, 75N92022D00004, 75N92022D00005). The authors thank the staff and participants of the ARIC study for their important contributions. Funding was also supported by R01HL087641 and R01HL086694; National Human Genome Research Institute contract U01HG004402; and National Institutes of Health contract HHSN268200625226C. Infrastructure was partly supported by Grant Number UL1RR025005, a component of the National Institutes of Health and NIH Roadmap for Medical Research.

**ASCOT - Anglo-Scandinavian Outcomes Trial:** This work was supported by Pfizer, New York, NY, USA, for the ASCOT study and the collection of the ASCOT DNA repository; by Servier Research Group, Paris, France; and by Leo Laboratories, Copenhagen, Denmark. We thank all ASCOT trial participants, physicians, nurses, and practices in the participating countries for their important contribution to the study. In particular we thank Clare Muckian and David Toomey for their help in DNA extraction, storage, and handling. Genotyping of ASCOT-SC and the Exome chip in ASCOT-UK was funded by the National Institutes of Health Research, UK. This work forms part of the research program of the National Institutes of Health and Care Research (NIHR) Barts Biomedical Centre at Queen Mary University of London, UK.

**ASPREE (ASPirin in Reducing Events in the Elderly):** The ASPREE study and Healthy Ageing Biobank were supported by an ASPREE Flagship cluster grant (including the Commonwealth Scientific and Industrial Research Organisation, Monash University, Menzies Research Institute, Australian National University, University of Melbourne); and grants (U01AG029824 and U19AG062682) from the National Institute on Aging and the National Cancer Institute at the National Institutes of Health, by grants (334047 and 1127060) from the National Health and Medical Research Council (NHMRC) of Australia, and by Monash University and the Victorian Cancer Agency. Paul Lacaze is supported by a National Heart Foundation Future Leader Fellowship (102604) and a NHMRC Investigator Grant (2026325). Chenglong Yu is supported by a Vanguard Grant (108071-2024_VG) from the National Heart Foundation of Australia. John J. McNeil is supported by an NHMRC Leadership Fellowship (IG1173690).

**ASPS / ASPS-Fam:** The authors thank the staff and the participants for their valuable contributions. We thank Birgit Reinhart for her long-term administrative commitment, Elfi Hofer for the technical assistance at creating the DNA bank, Ing. Johann Semmler and Anita Harb for DNA sequencing and DNA analyses by TaqMan assays and Irmgard Poelzl for supervising the quality management processes after ISO9001 at the biobanking and DNA analyses.
The Medical University of Graz and the Steiermärkische Krankenanstaltengesellschaft support the databank of the ASPS/ASPS-Fam. The research reported in this article was funded by the Austrian Science Fund (FWF) grant numbers PI904, P20545-P05 and P13180 and supported by the Austrian National Bank Anniversary Fund, P15435 and the Austrian Ministry of Science under the aegis of the EU Joint Programme-Neurodegenerative Disease Research (JPND)-www.jpnd.eu.

**AugUR:** Cohort recruiting and cohort management was funded by the Federal Ministry of Education and Research (BMBF-01ER1206, BMBF-01ER1507, to Iris M. Heid) and by the German Research Foundation (DFG HE 3690/7-1 to Iris M. Heid and DFG BR 6028/2-1 to Caroline Brandl). Genome-wide genotyping and biomarker phenotyping was funded by the University of Regensburg for the Department of Genetic Epidemiology. The computational work supervised by Iris M. Heid was funded by the German Research Foundation (Project-ID 387509280 – SFB 1350 Subproject C6, Project-ID 509149993 - TRR 374/1 Subproject C6, INF).

**AWI-Gen (H3Africa)**

We acknowledge the funders, the AWI-Gen participants, field teams and investigators at all six study centres across the duration of the study. The AWI-Gen Collaborative Centre is funded by the National Human Genome Research Institute (NHGRI), the National Institute of Environmental Health Sciences (NIEHS), of the National Institutes of Health (NIH) under award number U54HG006938 and its supplements, as part of the H3Africa Consortium, and by the Department of Science and Innovation, South Africa, award number DST/CON 0056/2014.

**BBJ (Biobank Japan Project):** The investigators acknowledge the cooperation of our BBJ participants. This project and BBJ are supported by the Japan Agency for Medical Research and Development (AMED) (Grant Numbers: JP19km0605001, JP21kk0305013, JP21tm0424220, and JP21ck0106642) and the Japan Society for the Promotion of Science KAKENHI (Grant Numbers: JP20H00462 and 22K16090).

**BES (Beijing Eye Study)**: The Beijing Eye Study has been funded by

1. the National Natural Science Foundation of China (grant 82271086)
2. the Beijing Municipal of Health Reform and Development Project (grant 2019-4).

**BioMe:** The Mount Sinai Bio*Me* Biobank has been supported by The Andrea and Charles Bronfman Philanthropies and in part by Federal funds from the NHLBI and NHGRI (U01HG00638001; U01HG007417; X01HL134588). We thank all participants and all our recruiters who have assisted and continue to assist in data collection and management. This work was supported in part through the computational and data resources and staff expertise provided by Scientific Computing and Data at the Icahn School of Medicine at Mount Sinai and supported by the Clinical and Translational Science Awards (CTSA) grant UL1TR004419 from the National Center for Advancing Translational Sciences. Research reported in this publication was also supported by the Office of Research Infrastructure of the National Institutes of Health under award number S10OD026880 and S10OD030463. The content is solely the responsibility of the authors and does not necessarily represent the official views of the National Institutes of Health.

**BioVU (VUMC):** The dataset(s) used for the analyses described were obtained from Vanderbilt University Medical Center's BioVU which is supported by numerous sources: institutional funding, private agencies, and federal grants. These include the NIH funded Shared Instrumentation Grant S10RR025141; and CTSA grants UL1TR002243, UL1TR000445, and UL1RR024975. Genomic data are also supported by investigator-led projects that include U01HG004798, R01NS032830, RC2GM092618, P50GM115305, U01HG006378, U19HL065962, R01HD074711; and additional funding sources listed at: https://victr.vumc.org/biovu-funding/.

**BRIGHT (British Genetics of Hypertension):** The BRIGHT study is extremely grateful to all the patients who participated in the study and the BRIGHT nursing team. This work forms part of the research program of the National Institutes of Health and Care Research (NIHR) Barts Biomedical Centre at Queen Mary University of London, UK. The study was funded by the Medical Research Council of Great Britain (grant number: G9521010D) and the Wellcome Trust Strategic Award (grant number: 083948). The funders had no role in study design, data collection and analysis.

**CAGE-Amagasaki (Cardio-metabolic Genome Epidemiology Network, Amagasaki Study):** The CAGE Network studies were supported by grants for the Core Research for Evolutional Science and Technology (CREST) from the Japan Science Technology Agency; the Program for Promotion of Fundamental Studies in Health Sciences, National Institute of Biomedical Innovation Organization (NIBIO); and the Grant of National Center for Global Health and Medicine (NCGM).

**CAGE-KING** This study was supported by grants from AMED (JP19km0405216) and JSPS KAKENHI (16H06277, 22H04923, 22H03350, 23K24608, 21H03206, 24K02690).

**CARDIA (The Coronary Artery Risk Development in Young Adults)**: The Coronary Artery Risk Development in Young Adults Study (CARDIA) is conducted and supported by the National Heart, Lung, and Blood Institute (NHLBI) in collaboration with the University of Alabama at Birmingham (75N92023D00002 & 75N92023D00005), Northwestern University (75N92023D00004), University of Minnesota (75N92023D00006), and Kaiser Foundation Research Institute (75N92023D00003). Genotyping was funded as part of the NHLBI Candidate-gene Association Resource (N01-HC-65226) and the NHGRI Gene Environment Association Studies (GENEVA) (U01-HG004729, U01-HG04424, and U01-HG004446).

**CCHC (Cameron County Hispanic Cohort):** The CCHC is funded by the National Institutes of Health (NIH) CTSA UL1 TR00371, R01 HL142302-05A1, R01 DK127084-01A1, R01AG078452-01A1, CPRIT RP230063. We are grateful to all participants and members of the study team without whom this work would not be possible. The authors would like to thank the CCHC cohort team, particularly Rocío Uribe who recruited and interviewed the participants. Marcela Morris, BS, her teams for laboratory support; Valley Baptist Medical Center, Brownsville, Texas, for providing us space for our Center for Clinical and Translational Science Clinical Research Unit is located; and the community of Brownsville and the participants who so willingly participated in this study in their city. This study was funded in part by Center for Clinical and Translational Sciences, National Institutes of Health Clinical and Translational Award grant no. UL1 TR000371 from the National Center for Advancing Translational Sciences.

**CFS (The Cleveland Family Study)** was supported by grants from the National Institutes of Health (HL46380, M01 RR00080-39, T32-HL07567, RO1-46380).

**CHS (Cardiovascular Health Study):** This CHS research was supported by NHLBI contracts HHSN268201200036C, HHSN268200800007C, HHSN268201800001C, N01HC55222, N01HC85079, N01HC85080, N01HC85081, N01HC85082, N01HC85083, N01HC85086, 75N92021D00006; and NHLBI grants U01HL080295, R01HL085251, R01HL087652, R01HL105756, R01HL103612, R01HL120393, U01HL130114, and R01HL172803 with additional contribution from the National Institute of Neurological Disorders and Stroke (NINDS). Additional support was provided through R01AG023629 from the National Institute on Aging (NIA). A full list of principal CHS investigators and institutions can be found at CHS-NHLBI.org. The provision of genotyping data was supported in part by the National Center for Advancing Translational Sciences, CTSI grant UL1TR001881, and the National Institute of Diabetes and Digestive and Kidney Disease Diabetes Research Center (DRC) grant DK063491 to the Southern California Diabetes Endocrinology Research Center. The content is solely the responsibility of the authors and does not necessarily represent the official views of the National Institutes of Health.

**CILENTO STUDY:** The investigators acknowledge the populations of Campora, Gioi, and Cardile for their participation in the study. The Cilento study was supported by grants from the Assessorato Ricerca Regione Campania (POR CAMPANIA 2000/2006 MISURA 3.16); CNR project FOE-2021 DBA.AD005.225; the National Recovery and Resilience Plan (NRRP), Mission 4 Component 2 Investment 1.4 - Call for tender No. 3138 of 16 December 2021, rectified by Decree n.3175 of 18 December 2021 of Italian Ministry of University and Research funded by the European Union – NextGenerationEU; Project code CN_00000033, Concession Decree No. 1034 of 17 June 2022 adopted by the Italian Ministry of University and Research, CUP B83C22002930006, Project title “National Biodiversity Future Center – NBFC.

**CoLaus:** The CoLaus study was supported by research grants from GlaxoSmithKline, the Faculty of Biology and Medicine of Lausanne (Switzerland), the Swiss National Science Foundation (grants 3200B0–105993, 3200B0-118308, 33CSCO-122661, 33CS30-139468, 33CS30-148401, 33CS30_177535 and 3247730_204523) and the Swiss Personalized Health Network (grant 2018DRI01). The authors would like to express their gratitude to the participants in the Lausanne CoLaus study.

**CRONICAS:** The CRONICAS Cohort Study was supported by the National Heart, Lung and Blood Institute Global Health Initiative under the contract Global Health Activities in Developing Countries to Combat Non-Communicable Chronic Diseases (Project Number HHSN268200900033C). Genotyping was supported by American Heart Association grant number 18CDA34110116. We thank Yesenia Castillo who coordinated selection and shipment of the samples, Megan Grove, Claudette Huitt, Irina Strelets, Barbara Jones, Malini Udtha, Susan Rossi, and Facheng Luo for carrying out the DNA extraction, genotyping, and genotype calling, and Ninad Chaudhary for performing quality control and genotype imputation. We thank the research staff and participants of the CRONICAS Cohort Study, without whom this research would not be possible.

1. Miranda JJ, Bernabe-Ortiz A, Smeeth L, et al Addressing geographical variation in the progression of non-communicable diseases in Peru: the CRONICAS cohort study protocol BMJ Open 2012;2:e000610. doi: 10.1136/bmjopen-2011-000610

**DHS (Diabetes Heart Study):** The authors thank the investigators, staff, and participants of the DHS for their valuable contributions. This study was supported by the National Institutes of Health through R01 HL067348, R01 HL092301, and R01 AG058921.

**DHS-AA (African American Diabetes Heart Study):** The investigators acknowledge the cooperation of our Diabetes Heart Study (DHS) and AA-DHS participants. This work was supported by NIH R01 DK071891, R01 HL092301, R01 AG058921 and the General Clinical Research Center of Wake Forest School of Medicine M01-RR-07122.

**DIACORE:** Cohort recruiting and management was funded by the KfH Stiftung Präventivmedizin e.V. (Carsten A. Böger). Genome-wide genotyping was funded the Else Kröner-Fresenius-Stiftung (2012_A147), the KfH Stiftung Präventivmedizin and the University Hospital Regensburg. The German Research Foundation supported this work (DFG BO 3815/4-1 to Iris M. Heid and Carsten A. Böger, and Project-ID 387509280 – SFB 1350 subproject C6 to Iris M. Heid). The computational work supervised by Iris M. Heid was funded by the German Research Foundation (Project-ID 387509280 – SFB 1350 Subproject C6, Project-ID 509149993 - TRR 374/1 Subproject C6, INF).

**DR’s EXTRA (Dose-Responses to Exercise Training):** The DR's EXTRA Study was supported by the Ministry of Education and Culture of Finland (722 and 627;2004-2011), Academy of Finland (102318; 104943;123885; 211119), Kuopio University Hospital, Finnish Diabetes Association, Finnish Foundations for Cardiovascular Research, Päivikki and Sakari Sohlberg Foundation, by European Commission FP6 Integrated Project (EXGENESIS); LSHM-CT-2004-005272, City of Kuopio and Social Insurance Institution of Finland (4/26/2010).

**ELISABET (Enquête LItoral Souffle Air Biologie EnvironemenT):** We thank the University of Lille, the Pasteur Institute of Lille (especially the Preventive Medicine Centre, Specialized Biology Department and Occupational Medicine, and the Biological Resources Centre) and the Centre Hospitalier Général de Dunkerque (especially the Departments of Biology and Pneumology). We particularly thank the nurses, physicians and secretarial staff at the University of Lille and the Pasteur Institute of Lille. Lastly, we thank the French Ministère de l'Enseignement Supérieur et de la Recherche, the Hauts de France Region and the European Regional Development Fund for financial support.
This work was funded by Lille University Hospital (CHU de Lille, Lille, France), the Nord Pas-de-Calais Regional Council, and the European Regional Development Fund (ERDF-FEDER Presage N°36034) as part of the CPER Institut de Recherche en ENvironnement Industriel (IRENI) program. The work was also supported by the French government through the Programme Investissement d’Avenir (I-SITE ULNE / ANR-16-IDEX-0004 ULNE), managed by the Agence Nationale de la Recherche as part of the project “Santé Environnement: du risque territorial au risque individual” (also funded by Lille European Metropole). Lastly, this work is part of the CPER’s CLIMIBIO research project.

**EXCEED:** EXCEED is supported by the University of Leicester, the NIHR Leicester Respiratory Biomedical Research Centre, by Wellcome [202849, https://doi.org/10.35802/202849], by BREATHE - The Health Data Research Hub for Respiratory Health [UKR_PC_19004] in partnership with SAIL Databank, and by Cohort Access fees from studies funded by the Medical Research Council (MRC), BBRSC, NIHR, the UK Space Agency, and GSK. The views expressed are those of the author(s) and not necessarily those of the NIHR or the Department of Health and Social Care. It was previously supported by MRC grant G0902313. Our analyses of EXCEED used the ALICE and SPECTRE High Performance Computing Facilities at the University of Leicester. The EXCEED study gratefully acknowledges the support of all participants and staff who have contributed to the study. CJ is supported by a British Heart Foundation Research Excellence Award (RE/24/130031).

**FHS (Framingham Heart Study):** This research was conducted in part using data and resources from the Framingham Heart Study of the National Heart Lung and Blood Institute of the National Institutes of Health and Boston University School of Medicine. The analyses reflect intellectual input and resource development from the Framingham Heart Study investigators participating in the SNP Health Association Resource (SHARe) project. This work was partially supported by the National Heart, Lung and Blood Institute's Framingham Heart Study (Contract Nos. N01-HC-25195 and HSN268201500001I) and its contract with Affymetrix, Inc for genotyping services (Contract No. N02-HL-6-4278). This research was partially supported by grant R01DK122503 R01DK122503from the National Institute of Diabetes and Digestive and Kidney Diseases (MPIs: Kari North, Anne Justice, and Ching-Ti Liu). This research was supported in part by the Intramural Research Program of the National Institutes of Health (NIH). The contributions of the NIH authors were made as part of their official duties as NIH federal employees, are in compliance with agency policy requirements, and are considered Works of the United States Government. The findings and conclusions presented in this paper are those of the authors and do not necessarily reflect the views of the NIH or the U.S. Department of Health and Human Services.

**FUSION (Finland-United States Investigation of NIDDM Genetics):** The FUSION study was supported by DK093757, DK072193, DK062370, and ZIA-HG000024. Genotyping was conducted at the Genetic Resources Core Facility (GRCF) at the Johns Hopkins Institute of Genetic Medicine.

**GAPP - Genetic and phenotypic determinants of blood pressure and other cardiovascular risk factors:** The GAPP study was supported by the Liechtenstein Government, the Swiss Heart Foundation, the Swiss Society of Hypertension, the University of Basel, the University Hospital Basel, the Hanela Foundation, the Mach-Gaensslen Foundation, Schiller AG, and Novartis.

We thank the GAPP staff and all GAPP study participants for their important contributions.

**GENOA (Genetic Epidemiology Network of Arteriopathy):** Support for the Genetic Epidemiology Network of Arteriopathy (GENOA) was provided by the National Heart, Lung and Blood Institute (U01 HL054464, U01 HL054457, U01 HL054481, R01 HL087660, R01 HL119443, R01 HL085571). Genotyping was performed at the Mayo Clinic by Stephen Turner,

Mariza de Andrade, and Julie Cunningham. We thank Eric Boerwinkle and Megan Grove from the Human Genetics Center and Institute of Molecular Medicine and Division of Epidemiology, University of Texas Health Science Center, Houston, Texas, for their help with genotyping. We would also like to thank the families that participated in the GENOA study.

**GenSalt:** This work was supported by a cooperative agreement project grant (U01HL072507, R01HL087263, and R01HL090682) from the National Heart, Lung and Blood Institute, National Institutes of Health, Bethesda, MD

**GOLDN (Genetics of Diet and Lipid Lowering Network):** Support for the genome-wide association studies in GOLDN was provided by the National Heart, Lung, and Blood Institute grant U01HL072524-04 and R01HL091357.

**HANDLS (Healthy Aging in Neighborhoods of Diversity across the Life Span):** The Healthy Aging in Neighborhoods of Diversity across the Life Span (HANDLS) study was supported by the Intramural Research Program of the NIH, National Institute on Aging and the National Center on Minority Health and Health Disparities (project # Z01-AG000513 and human subjects protocol number 09-AG-N248). Data analyses for the HANDLS study utilized the high-performance computational resources of the Biowulf Linux cluster at the National Institutes of Health, Bethesda, MD. (http://biowulf.nih.gov; http://hpc.nih.gov)).

**HCHS/SOL:** The Hispanic Community Health Study/Study of Latinos is a collaborative study supported by contracts from the National Heart, Lung, and Blood Institute (NHLBI) to the University of North Carolina (HHSN268201300001I / N01-HC-65233), University of Miami (HHSN268201300004I / N01-HC-65234), Albert Einstein College of Medicine (HHSN268201300002I / N01-HC-65235), University of Illinois at Chicago (HHSN268201300003I / N01- HC-65236 Northwestern Univ), and San Diego State University (HHSN268201300005I / N01-HC-65237). The following Institutes/Centers/Offices have contributed to the HCHS/SOL through a transfer of funds to the NHLBI: National Institute on Minority Health and Health Disparities, National Institute on Deafness and Other Communication Disorders, National Institute of Dental and Craniofacial Research, National Institute of Diabetes and Digestive and Kidney Diseases, National Institute of Neurological Disorders and Stroke, NIH Institution-Office of Dietary Supplements. The Genetic Analysis Center at the University of Washington was supported by NHLBI and NIDCR contracts (HHSN268201300005C AM03 and MOD03).

**HRS (Health & Retirement Study):** HRS is supported by the National Institute on Aging (NIA U01AG009740). Genotyping was funded separately by the NIA (RC2 AG036495, RC4 AG039029). Our genotyping was conducted by the NIH Center for Inherited Disease Research

(CIDR) at Johns Hopkins University. Genotyping quality control and final preparation of the data were performed by the Kardia lab at the University of Michigan.

**HTO Study:** The principal acknowledgement is to the blood pressure patients and their families who contributed to the study. We also acknowledge the many contributions made by research nurses, technicians and students since the study commenced in 1993. Finally we acknowledge the important contributions of colleagues now deceased or retired, to this long-term project. The study has received funding from the UK Medical Research Council, the Wellcome Trust and the British Heart Foundation. B Keavney holds a British Heart Foundation personal chair.

**HUFS (Howard University Family Study)**: The Howard University Family Study was supported by National Institutes of Health grants S06GM008016-320107 to Charles Rotimi and S06GM008016-380111 to Adebowale Adeyemo. We thank the participants of the study, for which enrollment was carried out at the Howard University General Clinical Research Center, supported by National Institutes of Health grant 2M01RR010284. The contents of this publication are solely the responsibility of the authors and do not necessarily represent the official view of the National Institutes of Health. Genotyping support was provided by the Coriell Institute for Medical Research. This work was supported in part by the Intramural Program of the National Human Genome Research Institute (NHGRI), the National Institutes of Health. through the Center for Research on Genomics and Global Health (CRGGH). The CRGGH is also supported by the National Institute of Diabetes and Digestive and Kidney Diseases (NIDDK) and the Office of the Director at the NIH (Z01HG200362).

**HyperGEN (Hypertension Genetic Epidemiology Network):** The hypertension network is funded by cooperative agreements (U10) with NHLBI: HL54471, HL54472, HL54473, HL54495, HL54496, HL54497, HL54509, HL54515, and 2 R01 HL55673-12. The study involves: University of Utah: (Network Coordinating Center, Field Center, and Molecular Genetics Lab); Univ. of Alabama at Birmingham: (Field Center and Echo Coordinating and Analysis Center); Medical College of Wisconsin: (Echo Genotyping Lab); Boston University: (Field Center); University of Minnesota: (Field Center and Biochemistry Lab); University of North Carolina: (Field Center); Washington University: (Data Coordinating Center); Weil Cornell Medical College: (Echo Reading Center); National Heart, Lung, & Blood Institute. For a complete list of HyperGEN Investigators: http://www.biostat.wustl.edu/hypergen/Acknowledge.html

**INGI-CAR** (INGI-Carlantino)**:** The authors wish to thank the staff and the study participants. This work was supported by SENSAGING-Sensory decays and ageing (D70-PRINSENSAGING-19: CUP J94I19000930006).

**INGI-FVG:** The authors wish to thank the staff and the study participants. This work was supported by Italian Ministry of Health—RC 01/21 to MPC and D70-RESRICGIROTTO to GG.

**ING-VBI:** The authors wish to thank the staff and the study participants. This work was supported by Italian Ministry of Health - RC 35/17 and BENEFICENTIA Stiftung to GG.

**IRAS Family Study (Insulin Resistance Atherosclerosis Family Study):** The IRASFS was supported by the National Heart Lung and Blood Institute (HL060944, HL061019, and HL060919). Genotyping and analysis for this study was supported by the GUARDIAN Consortium with grant support from the National Institute of Diabetes, Digestive, and Kidney Diseases (NIDDK; DK085175 and DK118062) and in part by UL1TR000124 (CTSI) and DK063491 (DRC). The authors thank study investigators, staff, and participants for their valuable contributions.

**JHS (Jackson Heart Study)**: The Jackson Heart Study is supported by Contracts HHSN268201800010I, HHSN268201800011I, HHSN268201800012I, HHSN268201800013I, HHSN268201800014I, HHSN268201800015I from the National Heart Lung and Blood Institute (NHLBI) with additional support from the National Institute of Minority Health and Health Disparities (NIMHD).

The Jackson Heart Study (phs000964.v1.p1) whole genome sequencing was performed at the Northwest Genomic Center (HHSN268201100037C). Core support including centralized genomic read mapping and genotype calling along with variant quality metrics and filtering was provided by the “TOPMED Informatics Research Center (3R01HL-117626-02S1; contract HHSN26820100002I). Core support including phenotype harmonization, data management, sample-identity QC, and general program coordination were provided by the TOPMED Data Coordinating Center (R01HL-120393; contract HHSN262801800001I).”

The manuscript has been reviewed by JHS for scientific content. The views expressed in this manuscript are those of the authors and do not necessarily represent the views of the National Heart, Lung, and Blood Institute; the National Institutes of Health; or the U.S. Department of Health and Human Services.

**JUPITER** (Justification for the Use of Statins in Primary Prevention: An Intervention Trial Evaluating Rosuvastatin): The JUPITER trial was supported by AstraZeneca (Paul Ridker, PI). Funding for genotyping in JUPITER was provided by a grant from AstraZeneca to PMR and DC.

**KoGES (Korean Genome and Epidemiology Study)**: Data were collected and supported by the National Research Institute of Health, Centers for Disease Control and Prevention, and Ministry for Health and Welfare, Republic of Korea. The authors thank the staff and participants of the KoGES study for their important contributions. Additional support was provided through grants the Brain Pool Plus (BP+, Brain Pool+) Program (2020H1D3A2A03100666) and Sejong

Science Fellowship (2022R1C1C2006474) through the National Research Foundation of Korea (NRF) funded by the Ministry of Science and ICT.

**LBC1921 (Lothian Birth Cohort 1921):** The authors thank all LBC1921 study participants and research team members who have contributed, and continue to contribute, to ongoing studies. LBC1921 was supported by the UK's Biotechnology and Biological Sciences Research Council (BBSRC), The Royal Society, and The Chief Scientist Office of the Scottish Government. Genotyping was funded by the BBSRC (BB/F019394/1).

**LBC1936 (Lothian Birth Cohort 1936):** The authors thank all LBC1936 study participants and research team members who have contributed, and continue to contribute, to ongoing studies. LBC1936 is supported by the Biotechnology and Biological Sciences Research Council (BBSRC), and the Economic and Social Research Council [BB/W008793/1] (which supports SEH), Age UK (Disconnected Mind project), and the University of Edinburgh. SRC is supported by a Sir Henry Dale Fellowship jointly funded by the Wellcome Trust and the Royal Society (221890/Z/20/Z). Genotyping was funded by the BBSRC (BB/F019394/1).

**Lifelines Cohort Study:**
Raul Aguirre-Gamboa (1), Patrick Deelen (1), Lude Franke (1), Jan A Kuivenhoven (2), Esteban A Lopera Maya (1), Ilja M Nolte (3), Serena Sanna (1), Harold Snieder (3), Morris A Swertz (1), Peter M. Visscher (3,4), Judith M Vonk (3), Cisca Wijmenga (1), Naomi Wray (4)

(1) *Department of Genetics, University of Groningen, University Medical Center Groningen, The Netherlands*
(2) *Department of Pediatrics, University of Groningen, University Medical Center Groningen, The Netherlands*
(3) *Department of Epidemiology, University of Groningen, University Medical Center Groningen, The Netherlands*
(4) *Institute for Molecular Bioscience, The University of Queensland, Brisbane, Queensland, Australia.*

The Lifelines Biobank initiative has been made possible by funding from the Dutch Ministry of Health, Welfare and Sport, the Dutch Ministry of Economic Affairs, the University Medical Center Groningen (UMCG the Netherlands), University of Groningen and the Northern Provinces of the Netherlands. The generation and management of GWAS genotype data for the Lifelines Cohort Study is supported by the UMCG Genetics Lifelines Initiative (UGLI). UGLI is partly supported by a Spinoza Grant from NWO, awarded to Cisca Wijmenga.
The authors wish to acknowledge the services of the Lifelines Cohort Study, the contributing research centers delivering data to Lifelines, and all the study participants.

**Living Biobank – for contributing data LivingBiobank-CHS and LivingBiobank-MAS**

The MEC1 and SH2012 studies were funded by individual research and clinical scientist award schemes from the National Medical Research Council (NMRC) and the Biomedical Research Council (BMRC) of Singapore, and infrastructure funding from the Singapore Ministry of Health (Population Health Metrics and Analytics PHMA), National University of Singapore and National University Health System, Singapore. Genome Institute of Singapore provided services for genotyping. Genotyping and whole-exome sequencing for Living Biobank was jointly funded by Agency for Science, Technology and Research, Singapore and Merck Sharp & Dohme Corp., Whitehouse Station, NJ USA.

**LLFS (The Long Life Family Study):** This work was supported by the National Institute on Aging (U01AG023746, U01AG023712, U01AG023749, U01AG023755, U01AG023744, and U19AG063893).

**MESA (Multi-Ethnic Study of Atherosclerosis)** MESA and the MESA SHARe project are conducted and supported by the National Heart, Lung, and Blood Institute (NHLBI) in collaboration with MESA investigators. Support for MESA is provided by contracts 75N92025D00022, 75N92020D00001, HHSN268201500003I, N01-HC-95159, 75N92025D00026, 75N92020D00005, N01-HC-95160, 75N92020D00002, N01-HC-95161, 75N92025D00024, 75N92020D00003, N01-HC-95162, 75N92025D00027, 75N92020D00006, N01-HC-95163, 75N92025D00025, 75N92020D00004, N01-HC-95164, 75N92025D00028, 75N92020D00007, N01-HC-95165, N01-HC-95166, N01-HC-95167, N01-HC-95168, N01-HC-95169, UL1-TR-000040, UL1-TR-001079, UL1-TR-001420, UL1TR001881, DK063491, and R01HL105756. The authors thank the MESA participants and the MESA investigators and staff for their valuable contributions. A full list of participating MESA investigators and institutions can be found at http://www.mesa-nhlbi.org.

**METSIM:** The METSIM study was supported by grants from the Academy of Finland (321428), Sigrid Juselius Foundation, Finnish Foundation for Cardiovascular Research, and Centre of Excellence of Cardiovascular and Metabolic Diseases supported by the Academy of Finland (0245896-3).

**Million Veteran Program (MVP):** VA Million Veteran Program: MVP work was supported by MVP000 as well as a BLR&D merit award #BX003360 title Pharmacogenomics of risk factors and therapies outcomes for kidney disease Kidney disease (PI: Adriana M. Hung) & #CX001897 Genetic of Kidney Disease and Hypertension in MVP II (PI. Adriana M. Hung).

This publication does not represent the views of the Department of Veteran Affairs or the United States Government. This research is based on data from the Million Veteran Program, Office of Research and Development, Veterans Health Administration.

MVP Program Office

- Sumitra Muralidhar, Ph.D., Program Director
  US Department of Veterans Affairs, 810 Vermont Avenue NW, Washington, DC 20420
- Jennifer Moser, Ph.D., Associate Director, Scientific Programs
  US Department of Veterans Affairs, 810 Vermont Avenue NW, Washington, DC 20420
- Jennifer E. Deen, B.S., Associate Director, Cohort & Public Relations
  US Department of Veterans Affairs, 810 Vermont Avenue NW, Washington, DC 20420

**MVP Executive Committee**

- Co-Chair: Philip S. Tsao, Ph.D.
  VA Palo Alto Health Care System, 3801 Miranda Avenue, Palo Alto, CA 94304
- Co-Chair: Sumitra Muralidhar, Ph.D.
  US Department of Veterans Affairs, 810 Vermont Avenue NW, Washington, DC 20420
- J. Michael Gaziano, M.D., M.P.H.
  VA Boston Healthcare System, 150 S. Huntington Avenue, Boston, MA 02130
- Elizabeth Hauser, Ph.D.
  Durham VA Medical Center, 508 Fulton Street, Durham, NC 27705
- Amy Kilbourne, Ph.D., M.P.H.
  VA HSR&D, 2215 Fuller Road, Ann Arbor, MI 48105
- Michael Matheny, M.D., M.S., M.P.H.
  VA Tennessee Valley Healthcare System, 1310 24$^{th}$ Ave. South, Nashville, TN 37212
- Dave Oslin, M.D.
  Philadelphia VA Medical Center, 3900 Woodland Avenue, Philadelphia, PA 19104

**MVP Co-Principal Investigators**

- J. Michael Gaziano, M.D., M.P.H.
  VA Boston Healthcare System, 150 S. Huntington Avenue, Boston, MA 02130
- Philip S. Tsao, Ph.D.
  VA Palo Alto Health Care System, 3801 Miranda Avenue, Palo Alto, CA 94304

**MVP Core Operations**

- Lori Churby, B.S., Director, MVP Regulatory Affairs
  VA Palo Alto Health Care System, 3801 Miranda Avenue, Palo Alto, CA 94304
- Stacey B. Whitbourne, Ph.D., Director, MVP Cohort Management
  VA Boston Healthcare System, 150 S. Huntington Avenue, Boston, MA 02130
- Jessica V. Brewer, M.P.H., Director, MVP Recruitment & Enrollment
  VA Boston Healthcare System, 150 S. Huntington Avenue, Boston, MA 02130
- Shahpoor (Alex) Shayan, M.S., Director, MVP Recruitment and Enrollment Informatics
  VA Boston Healthcare System, 150 S. Huntington Avenue, Boston, MA 02130
- Luis E. Selva, Ph.D., Executive Director, MVP Biorepositories

VA Boston Healthcare System, 150 S. Huntington Avenue, Boston, MA 02130

- Saiju Pyarajan Ph.D., Director, Data and Computational Sciences
  VA Boston Healthcare System, 150 S. Huntington Avenue, Boston, MA 02130
- Kelly Cho, M.P.H, Ph.D., Director, MVP Phenomics Data Core
  VA Boston Healthcare System, 150 S. Huntington Avenue, Boston, MA 02130
- Scott L. DuVall, Ph.D., Director, VA Informatics and Computing Infrastructure (VINCI)
  VA Salt Lake City Health Care System, 500 Foothill Drive, Salt Lake City, UT 84148
- Mary T. Brophy M.D., M.P.H., Director, VA Central Biorepository
  VA Boston Healthcare System, 150 S. Huntington Avenue, Boston, MA 02130
- MVP Coordinating Centers
  - MVP Coordinating Center, Boston - J. Michael Gaziano, M.D., M.P.H.
    VA Boston Healthcare System, 150 S. Huntington Avenue, Boston, MA 02130
  - MVP Coordinating Center, Palo Alto – Philip S. Tsao, Ph.D.
    VA Palo Alto Health Care System, 3801 Miranda Avenue, Palo Alto, CA 94304
  - MVP Information Center, Canandaigua – Brady Stephens, M.S.
    Canandaigua VA Medical Center, 400 Fort Hill Avenue, Canandaigua, NY 14424
  - Cooperative Studies Program Clinical Research Pharmacy Coordinating Center, Albuquerque – Todd Connor, Pharm.D.; Dean P. Argyres, B.S., M.S.
    New Mexico VA Health Care System, 1501 San Pedro Drive SE, Albuquerque, NM 87108

**MVP Publications and Presentations Committee**

- Co-Chair: Themistocles L. Assimes, M.D., Ph. D
  VA Palo Alto Health Care System, 3801 Miranda Avenue, Palo Alto, CA 94304
- Co-Chair: Adriana Hung, M.D.; M.P.H
  VA Tennessee Valley Healthcare System, 1310 24th Ave. South, Nashville, TN 37212
- Co-Chair: Henry Kranzler, M.D.
  Philadelphia VA Medical Center, 3900 Woodland Avenue, Philadelphia, PA 19104

**MVP Local Site Investigators**

- Samuel Aguayo, M.D., Phoenix VA Health Care System
  650 E. Indian School Road, Phoenix, AZ 85012
- Sunil Ahuja, M.D., South Texas Veterans Health Care System
  7400 Merton Minter Boulevard, San Antonio, TX 78229
- Kathrina Alexander, M.D., Veterans Health Care System of the Ozarks
  1100 North College Avenue, Fayetteville, AR 72703
- Xiao M. Androulakis, M.D., Columbia VA Health Care System
  6439 Garners Ferry Road, Columbia, SC 29209
- Prakash Balasubramanian, M.D., William S. Middleton Memorial Veterans Hospital
  2500 Overlook Terrace, Madison, WI 53705
- Zuhair Ballas, M.D., Iowa City VA Health Care System
  601 Highway 6 West, Iowa City, IA 52246-2208

- Elizabeth S. Bast, M.D., M.P.H., Miami VA Health Care System
  1201 NW 16th Street, 11 GRC, Miami FL 33125
- Jean Beckham, Ph.D., Durham VA Medical Center
  508 Fulton Street, Durham, NC 27705
- Sujata Bhushan, M.D., VA North Texas Health Care System
  4500 S. Lancaster Road, Dallas, TX 75216
- Edward Boyko, M.D., VA Puget Sound Health Care System
  1660 S. Columbian Way, Seattle, WA 98108-1597
- David Cohen, M.D., Portland VA Medical Center
  3710 SW U.S. Veterans Hospital Road, Portland, OR 97239
- Louis Dellitalia, M.D., Birmingham VA Medical Center
  700 S. 19th Street, Birmingham AL 35233
- Gerald Wayne Dryden, Jr., M.D., Ph.D., Louisville VA Medical Center
  800 Zorn Avenue, Louisville, KY 40206
- L. Christine Faulk, M.D., Robert J. Dole VA Medical Center
  5500 East Kellogg Drive, Wichita, KS 67218-1607
- Joseph Fayad, M.D., VA Southern Nevada Healthcare System
  6900 North Pecos Road, North Las Vegas, NV 89086
- Daryl Fujii, Ph.D., VA Pacific Islands Health Care System
  459 Patterson Rd, Honolulu, HI 96819
- Saib Gappy, M.D., John D. Dingell VA Medical Center
  4646 John R Street, Detroit, MI 48201
- Frank Gesek, Ph.D., White River Junction VA Medical Center
  163 Veterans Drive, White River Junction, VT 05009
- Jennifer Greco, M.D., Sioux Falls VA Health Care System
  2501 W 22nd Street, Sioux Falls, SD 57105
- Michael Godschalk, M.D., Richmond VA Medical Center
  1201 Broad Rock Blvd., Richmond, VA 23249
- Todd W. Gress, M.D., Ph.D., Hershel “Woody” Williams VA Medical Center
  1540 Spring Valley Drive, Huntington, WV 25704
- Samir Gupta, M.D., M.S.C.S., VA San Diego Healthcare System
  3350 La Jolla Village Drive, San Diego, CA 92161
- Salvador Gutierrez, M.D., Edward Hines, Jr. VA Medical Center
  5000 South 5th Avenue, Hines, IL 60141
- John Harley, M.D., Ph.D., Cincinnati VA Medical Center
  3200 Vine Street, Cincinnati, OH 45220
- Mark Hamner, M.D., Ralph H. Johnson VA Medical Center
  109 Bee Street, Mental Health Research, Charleston, SC 29401
- Daniel J. Hogan, M.D., Bay Pines VA Healthcare System
  10,000 Bay Pines Blvd Bay Pines, FL 33744
- Adriana Hung, M.D., M.P.H., VA Tennessee Valley Healthcare System
  1310 24th Avenue, South Nashville, TN 37212
- Robin Hurley, M.D., W.G. (Bill) Hefner VA Medical Center
  1601 Brenner Ave, Salisbury, NC 28144

- Pran Iruvanti, D.O., Ph.D., Hampton VA Medical Center
  100 Emancipation Drive, Hampton, VA 23667
- Frank Jacono, M.D., VA Northeast Ohio Healthcare System
  10701 East Boulevard, Cleveland, OH 44106
- Darshana Jhala, M.D., Philadelphia VA Medical Center
  3900 Woodland Avenue, Philadelphia, PA 19104
- Scott Kinlay, M.B.B.S., Ph.D., VA Boston Healthcare System
  150 S. Huntington Avenue, Boston, MA 02130
- Michael Landry, Ph.D., Southeast Louisiana Veterans Health Care System
  2400 Canal Street, New Orleans, LA 70119
- Peter Liang, M.D., M.P.H., VA New York Harbor Healthcare System
  423 East 23rd Street, New York, NY 10010
- Suthat Liangpunsakul, M.D., M.P.H., Richard Roudebush VA Medical Center
  1481 West 10th Street, Indianapolis, IN 46202
- Jack Lichy, M.D., Ph.D., Washington DC VA Medical Center
  50 Irving St, Washington, D. C. 20422
- Tze Shien Lo, M.D., Fargo VA Health Care System
  2101 N. Elm, Fargo, ND 58102
- C. Scott Mahan, M.D., Charles George VA Medical Center
  1100 Tunnel Road, Asheville, NC 28805
- Ronnie Marrache, M.D., VA Maine Healthcare System Center, Augusta, ME 04330
- Stephen Mastorides, M.D., James A. Haley Veterans' Hospital
  13000 Bruce B. Downs Blvd, Tampa, FL 33612
- Kristin Mattocks, Ph.D., M.P.H., Central Western Massachusetts Healthcare System
  421 North Main Street, Leeds, MA 01053
- Paul Meyer, M.D., Ph.D., Southern Arizona VA Health Care System
  3601 S 6th Avenue, Tucson, AZ 85723
- Jonathan Moorman, M.D., Ph.D., James H. Quillen VA Medical Center
  Corner of Lamont & Veterans Way, Mountain Home, TN 37684
- Providencia Morales, R.N., Northern Arizona VA Health Care System
  500 Highway 89 North, Prescott, AZ 86313
- Timothy Morgan, M.D., VA Long Beach Healthcare System
  5901 East 7th Street Long Beach, CA 90822
- Maureen Murdoch, M.D., M.P.H., Minneapolis VA Health Care System
  One Veterans Drive, Minneapolis, MN 55417
- Eknath Naik, M.D., Ph.D., West Palm Beach VA Medical Center,
  7305 North Military Trail, West Palm Beach, FL 33410-6400
- James Norton, Ph.D., VA Health Care Upstate New York
  113 Holland Avenue, Albany, NY 12208
- Olaoluwa Okusaga, M.D., Michael E. DeBakey VA Medical Center
  2002 Holcombe Blvd, Houston, TX 77030
- Michael K. Ong, M.D., VA Greater Los Angeles Health Care System
  11301 Wilshire Blvd, Los Angeles, CA 90073
- Kris Ann Oursler, M.D., Salem VA Medical Center

1970 Roanoke Blvd, Salem, VA 24153

- Ismene Petrakis, M.D., VA Connecticut Healthcare System
  950 Campbell Avenue, West Haven, CT 06516
- Samuel Poon, M.D., Manchester VA Medical Center
  718 Smyth Road, Manchester, NH 03104
- Emily Potter, Pharm.D., VA Eastern Kansas Health Care System
  4101 S 4th Street Trafficway, Leavenworth, KS 66048
- Michael Rauchman, M.D., St. Louis VA Health Care System
  915 North Grand Blvd, St. Louis, MO 63106
- Amneet S. Rai, Pharm.D., VA Sierra Nevada Health Care System
  975 Kirman Avenue, Reno, NV 89502
- Richard Servatius, Ph.D., Syracuse VA Medical Center
  800 Irving Avenue, Syracuse, NY 13210
- Satish Sharma, M.D., Providence VA Medical Center
  830 Chalkstone Avenue, Providence, RI 02908
- River Smith, Ph.D., Eastern Oklahoma VA Health Care System
  1011 Honor Heights Drive, Muskogee, OK 74401
- Peruvemba Sriram, M.D., N. FL/S. GA Veterans Health System
  1601 SW Archer Road, Gainesville, FL 32608
- Patrick Strollo, Jr., M.D., VA Pittsburgh Health Care System
  University Drive, Pittsburgh, PA 15240
- Neeraj Tandon, M.D., Overton Brooks VA Medical Center
  510 East Stoner Ave, Shreveport, LA 71101
- Philip Tsao, Ph.D., VA Palo Alto Health Care System
  3801 Miranda Avenue, Palo Alto, CA 94304-1290
- Gerardo Villareal, M.D., New Mexico VA Health Care System
  1501 San Pedro Drive, S.E. Albuquerque, NM 87108
- Jessica Walsh, M.D., VA Salt Lake City Health Care System
  500 Foothill Drive, Salt Lake City, UT 84148
- John Wells, Ph.D., Edith Nourse Rogers Memorial Veterans Hospital
  200 Springs Road, Bedford, MA 01730
- Jeffrey Whittle, M.D., M.P.H., Clement J. Zablocki VA Medical Center
  5000 West National Avenue, Milwaukee, WI 53295
- Mary Whooley, M.D., San Francisco VA Health Care System
  4150 Clement Street, San Francisco, CA 94121
- Peter Wilson, M.D., Atlanta VA Medical Center
  1670 Clairmont Road, Decatur, GA 30033
- Junzhe Xu, M.D., VA Western New York Healthcare System
  3495 Bailey Avenue, Buffalo, NY 14215-1199
- Shing Shing Yeh, Ph.D., M.D., Northport VA Medical Center
  79 Middleville Road, Northport, NY 11768
- Andrew W. Yen, M.D., VA Northern California Health Care System
  10535 Hospital Way, Mather, CA 95655

**MONA LISA Lille (Monitoring National du Risque Artériel; National Monitoring of Arterial Risk):** The MONA LISA Study was funded by an unrestricted grant from Pfizer and by the French National Research Agency (ANR-05-PNRA-018).

The authors would like to thank the study nurses, physicians, dieticians, computer scientists and secretaries, the University of Lille, the Pasteur Institute of Lille (the Preventive Medicine Centre, Specialized Biology Department and Biological Resources Centre), and participating municipal authorities.

**NEO (The Netherlands Epidemiology of Obesity study):** The authors of the NEO study thank all individuals who participated in the Netherlands Epidemiology in Obesity study, all participating general practitioners for inviting eligible participants and all research nurses for collection of the data. We thank the NEO study group, Petra Noordijk, Pat van Beelen and Ingeborg de Jonge for the coordination, lab and data management of the NEO study. The genotyping in the NEO study was supported by the Centre National de Génotypage (Paris, France), headed by Jean-Francois Deleuze. The NEO study is supported by the participating Departments, the Division and the Board of Directors of the Leiden University Medical Center, and by the Leiden University, Research Profile Area Vascular and Regenerative Medicine.

**NESDA:** The infrastructure for the NESDA study (www.nesda.nl) is funded through the Geestkracht program of the Netherlands Organisation for Health Research and Development (ZonMw, grant number 10-000-1002) and financial contributions by participating universities and mental health care organizations (VU University Medical Center, GGZ inGeest, Leiden University Medical Center, Leiden University, GGZ Rivierduinen, University Medical Center Groningen, University of Groningen, Lentis, GGZ Friesland, GGZ Drenthe, Rob Giel Onderzoekscentrum).

**PROCARDIS:** PROCARDIS was supported by the European Community Sixth Framework Program (LSHM-CT-2007-037273), AstraZeneca, the Swedish Research Council, the Knut and Alice Wallenberg Foundation, the Swedish Heart-Lung Foundation, the Torsten and Ragnar Soderberg Foundation, the Strategic Cardiovascular Program of Karolinska Institutet and Stockholm County Council, the Foundation for Strategic Research and the Stockholm County Council (560283). Wellcome Trust core award (090532/Z/09/Z, 203141/Z/16/Z, 201543/B/16/Z); HEALTH-F2-2013-601456 (CVGenes@Target), the TriPartite Immunometabolism Consortium [TrIC]-Novo Nordisk Foundation's Grant number NNF15CC0018486, VIAgenomics (SP/19/2/344612) and support from the NIHR Oxford Biomedical Research Centre. The views expressed are those of the author(s) and not necessarily those of the NHS, the NIHR or the Department of Health.

M. Sabater-Lleal is supported by a Miguel Servet contract from the ISCIII Spanish Health Institute (CPII22/00007) and co-financed by the European Social Fund, and acknowledges funding from the CERCA Programme/Generalitat de Catalunya.

**PROSPER:** The PROSPER study was supported by an investigator initiated grant obtained from Bristol-Myers Squibb. Prof. Dr. J. W. Jukema is an Established Clinical Investigator of the Netherlands Heart Foundation (grant 2001 D 032). Support for genotyping was provided by the seventh framework program of the European commission (grant 223004) and by the Netherlands Genomics Initiative (Netherlands Consortium for Healthy Aging grant 050-060-810).

PMID: 10569329; PMID:12457784; PMID: 21977987

**References:**
(1-3)

(1) Shepherd J, Blauw GJ, Murphy MB, Cobbe SM, Bollen EL, Buckley BM, et al. The design of a prospective study of Pravastatin in the Elderly at Risk (PROSPER). PROSPER Study Group. PROspective Study of Pravastatin in the Elderly at Risk. Am J Cardiol 1999 Nov 15;84(10):1192-7. PMID: 10569329

(2) Shepherd J, Blauw GJ, Murphy MB, Bollen EL, Buckley BM, Cobbe SM, et al. Pravastatin in elderly individuals at risk of vascular disease (PROSPER): a randomised controlled trial. Lancet 2002 Nov 23;360(9346):1623-30. PMID:12457784

(3) Trompet S, Craen de AJM, Postmus I, Ford I, Sattar N, Stott DJ, et al. Replication of LDL GWAs hits in PROSPER/PHASE as validation for future (pharmaco)genetic analyses. BMC Med Genetics 2011 Oct 6;12:131. PMID: 21977987

**RHS (Ragama Health Study):** The RHS was supported by the Grant of National Center for Global Health and Medicine (NCGM).

**SEED (Singapore Epidemiology of Eye Diseases) – for contributing data SCES-610, SCES-Omni, SiMES, and SINDI**
SCES (Singapore Chinese Eye Study), SiMES (Singapore Malay Eye Study), SINDI (Singapore Indian Eye Study): The Singapore Malay Eye Study (SiMES), the Singapore Indian Eye Study (SINDI), and the Singapore Chinese Eye Study (SCES) are supported by the National Medical Research Council (NMRC), Singapore (grants 0796/2003, 1176/2008, 1149/2008, STaR/0003/2008, 1249/2010, CG/SERI/2010, CIRG/1371/2013, and CIRG/1417/2015), and Biomedical Research Council (BMRC), Singapore (08/1/35/19/550 and 09/1/35/19/616). Ching-

Yu Cheng is supported by an award from NMRC (CSA/033/2012). The Singapore Tissue Network and the Genome Institute of Singapore, Agency for Science, Technology and Research, Singapore provided services for tissue archival and genotyping, respectively.

**SHIP (Study of Health in Pomerania):** SHIP is part of the Community Medicine Research net of the University of Greifswald, Germany, which is funded by the Federal Ministry of Education and Research (grants no. 01ZZ9603, 01ZZ0103, and 01ZZ0403), the Ministry of Cultural Affairs as well as the Social Ministry of the Federal State of Mecklenburg-West Pomerania, and the network 'Greifswald Approach to Individualized Medicine (GANI_MED)' funded by the Federal Ministry of Education and Research (grant 03IS2061A). Genome-wide data were supported by the Federal Ministry of Education and Research (grant no. 03ZIK012) and a joint grant from Siemens Healthcare, Erlangen, Germany and the Federal State of Mecklenburg- West Pomerania.

**Singapore Prospective Study Program (SP2) – for contributing data SP2-1M and SP2-610**

SP2 (Singapore Prospective Study Program): The SP2 study is supported by individual research and clinical scientist award schemes from the National Medical Research Council (NMRC) and the Biomedical Research Council (BMRC) of Singapore, and infrastructure funding from the Singapore Ministry of Health (Population Health Metrics and Analytics PHMA), National University of Singapore and National University Health System, Singapore. The Genome Institute of Singapore, Agency for Science, Technology and Research, Singapore provided services for genotyping.

**The Study of Women's Health Across the Nation (SWAN)**: The Study of Women's Health Across the Nation (SWAN) has grant support from the National Institutes of Health (NIH), DHHS, through the National Institute on Aging (NIA), the National Institute of Nursing Research (NINR) and the NIH Office of Research on Women's Health (ORWH) (Grants U01NR004061; U01AG012505, U01AG012535, U01AG012531, U01AG012539, U01AG012546, U01AG012553, U01AG012554, U01AG012495 and U19AG063720 and The SWAN Repository (U01AG017719)). The content of this article is solely the responsibility of the authors and does not necessarily represent the official views of the NIA, NINR, ORWH or the NIH. Clinical Centers: University of Michigan, Ann Arbor – Carrie Karvonen-Gutierrez PI, 2021-present, Siobán Harlow, PI 2011 – 2021, MaryFran Sowers, PI 1994-2011; Massachusetts General Hospital, Boston, MA – Massachusetts General Hospital, Boston, MA – Sherri-Ann Burnett-Bowie, PI 2020 – Present; Joel Finkelstein, PI 1999 – 2020; Robert Neer, PI 1994 – 1999; Rush University, Rush University Medical Center, Chicago, IL – Imke Janssen, PI 2020 –Present; Howard Kravitz, PI 2009 – 2020; Lynda Powell, PI 1994 – 2009; University of California, Davis/Kaiser – Elaine Waetjen and Monique Hedderson, PIs 2020 – Present; Ellen Gold, PI 1994 - 2020; University of California, Los Angeles – Arun Karlamangla, PI 2020 – Present; Gail Greendale, PI 1994 - 2020; Albert Einstein College of Medicine, Bronx, NY – Carol Derby, PI

2011 – present, Rachel Wildman, PI 2010 – 2011; Nanette Santoro, PI 2004 – 2010; University of Medicine and Dentistry – New Jersey Medical School, Newark – Gerson Weiss, PI 1994 – 2004; and the University of Pittsburgh, Pittsburgh, PA – Rebecca Thurston, PI 2020 – Present; Karen Matthews, PI 1994 - 2020.

NIH Program Office: National Institute on Aging, Bethesda, MD – Rosaly Correa-de-Araujo 2020 - present; Chhanda Dutta 2016- present; Winifred Rossi 2012–2016; Sherry Sherman 1994 – 2012; Marcia Ory 1994 – 2001; National Institute of Nursing Research, Bethesda, MD – Program Officers. Central Laboratory: University of Michigan, Ann Arbor – Daniel McConnell (Central Ligand Assay Satellite Services). NIA Biorepository - Rosaly Correa-de-Araujo 2019 - Present; SWAN Repository: University of Michigan, Ann Arbor – Siobán Harlow 2013- present; Dan McConnell 2011 - 2013; MaryFran Sowers 2000 – 2011. Coordinating Center: University of Pittsburgh, Pittsburgh, PA – Maria Mori Brooks, PI 2012 - present; Kim Sutton-Tyrrell, PI 2001 – 2012; New England Research Institutes, Watertown, MA - Sonja McKinlay, PI 1995 – 2001. Steering Committee: Susan Johnson, Current Chair; Chris Gallagher, Former Chair. We thank the study staff at each site and all the women who participated in SWAN.

**UKBB (UK Biobank):** This research has been conducted using the UK Biobank Resource under Application Number 8343. This research used data assets made available by National Safe Haven as part of the Data and Connectivity National Core Study, led by Health Data Research UK in partnership with the Office for National Statistics and funded by UK Research and Innovation (grant ref MC_PC_20029). Copyright © (2022), NHS Digital. Re-used with the permission of the NHS Digital [and/or UK Biobank]. All rights reserved.

**WGHS** (Women's Genome Health Study): The WGHS is supported by the National Heart, Lung, and Blood Institute (HL043851 and HL080467) and the National Cancer Institute (CA047988 and UM1CA182913), with collaborative scientific support and funding for genotyping provided by Amgen.

**WHI (Women's Health Initiative)**: The WHI program is funded by the National Heart, Lung, and Blood Institute, National Institutes of Health, U.S. Department of Health and Human Services through 75N92021D00001, 75N92021D00002, 75N92021D00003, 75N92021D00004, 75N92021D00005. The authors thank the WHI investigators and staff for their dedication, and the study participants for making the program possible. A full listing of WHI investigators can be found at: https://www-whi-org.s3.us-west-2.amazonaws.com/wp-content/uploads/WHI-Investigator-Long-List.pdf. Additional support to NF was provided by NIH DK117445 and MD012765.

**YFS (The Cardiovascular Risk in Young Finns Study):** The Young Finns Study has been financially supported by the Academy of Finland: grants 356405, 322098, 286284, 134309 (Eye), 126925, 121584, 124282, 129378 (Salve), 117797 (Gendi), and 141071 (Skidi); the

Social Insurance Institution of Finland; Competitive State Research Financing of the Expert Responsibility area of Kuopio, Tampere and Turku University Hospitals (grant X51001); Juho Vainio Foundation; Paavo Nurmi Foundation; Finnish Foundation for Cardiovascular Research ; Finnish Cultural Foundation; The Sigrid Juselius Foundation; Tampere Tuberculosis Foundation; Emil Aaltonen Foundation; Yrjö Jahnsson Foundation; Signe and Ane Gyllenberg Foundation; Diabetes Research Foundation of Finnish Diabetes Association; EU Horizon 2020 (grant 755320 for TAXINOMISIS and grant 848146 for To Aition); European Research Council (grant 742927 for MULTIEPIGEN project); Tampere University Hospital Supporting Foundation, Finnish Society of Clinical Chemistry, the Cancer Foundation Finland, and pBETTER4U_EU (Preventing obesity through Biologically and bEhaviorally Tailored inTERventions for you; project number: 101080117).

We thank the teams that collected data at all measurement time points; the persons who participated as both children and adults in these longitudinal studies; and biostatisticians Irina Lisinen, Johanna Ikonen, Noora Kartiosuo, Ville Aalto, and Jarno Kankaanranta for data management and statistical advice.
