## Supplementary Figures for "Large-Scale Gene-Smoking Interactions and Fine Mapping Study Identifies Multiple Novel Blood Pressure Loci in over 1 Million Individuals"

#### Supplementary Figure S1. Manhattan plots of the novel Locus

The Manhattan plots illustrate the novel loci identified in cross-population meta-analysis (CPMA) of the association between smoking exposures, current smoker (CURSMK), cigarettes per day (CPD), and pack year (PY), and blood pressure traits. Triangles represent the lead SNPs within these loci, while colored dots (excluding light blue and grey) indicate other SNPs within the corresponding loci. The marker names represent *chr:position:allele1_allele2*. A dotted red line marks the threshold for genome-wide significance at $-\log_{10} 5 \times 10^{-9}$.

A. The 2df joint test of DBP-CURSMK of males in CPMA

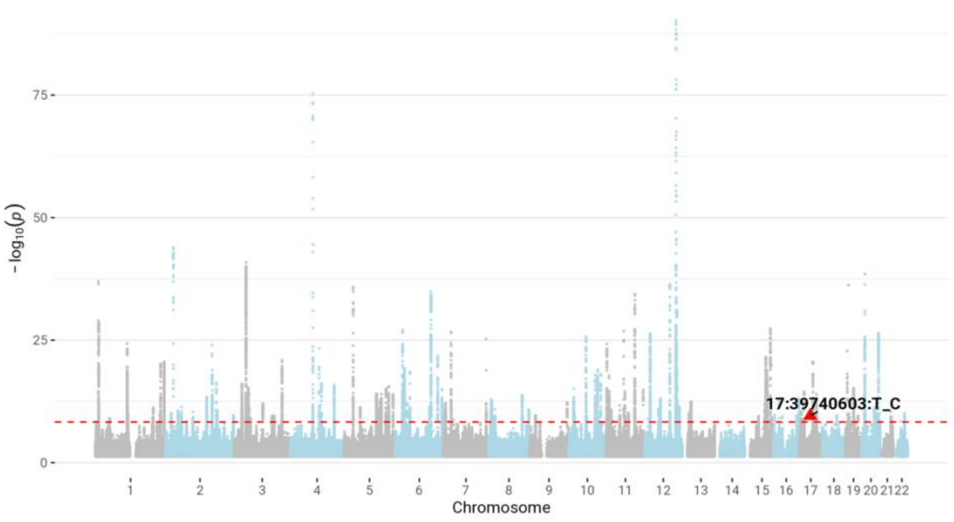


B. The 2df joint test of PP-CPD of combined sex analyses in CPMA

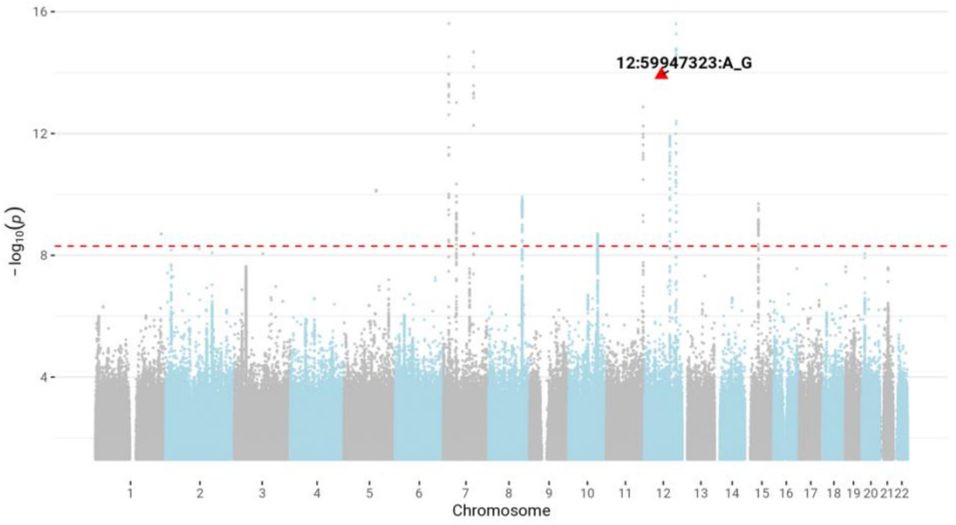


C. The 2df joint test of DBP-CURSMK of combined sex analyses in CPMA

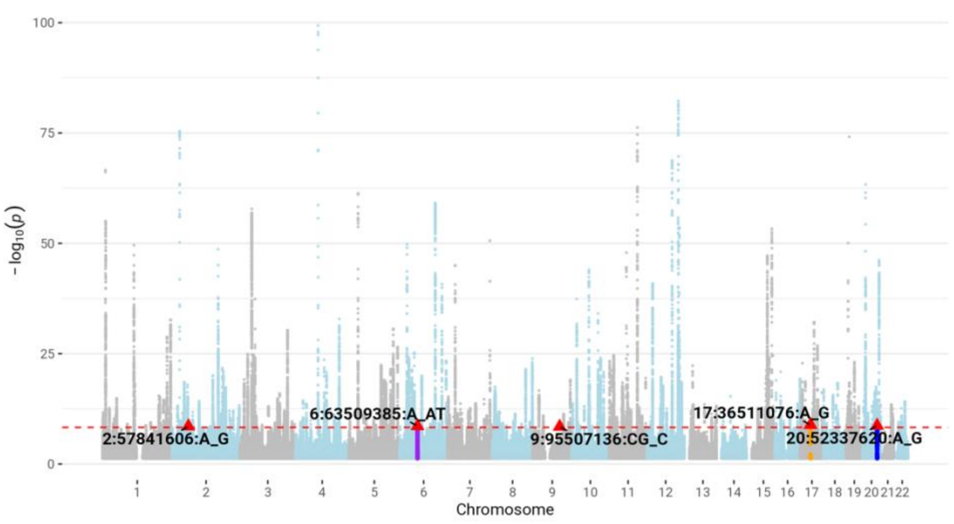


D. The 2df joint test of SBP-CURSMK of combined sex analyses in CPMA

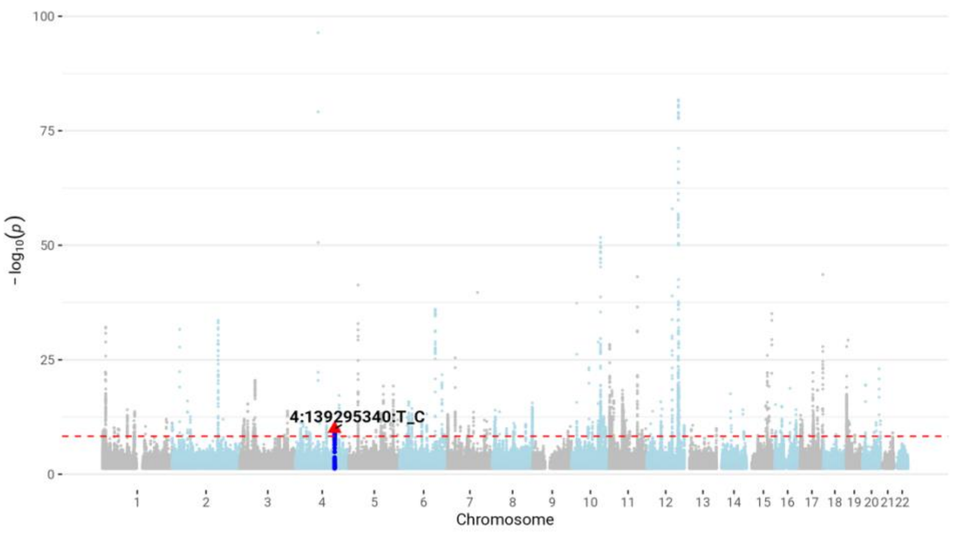

E. The 2df joint test of PP-CURSMK of combined sex analyses in CPMA

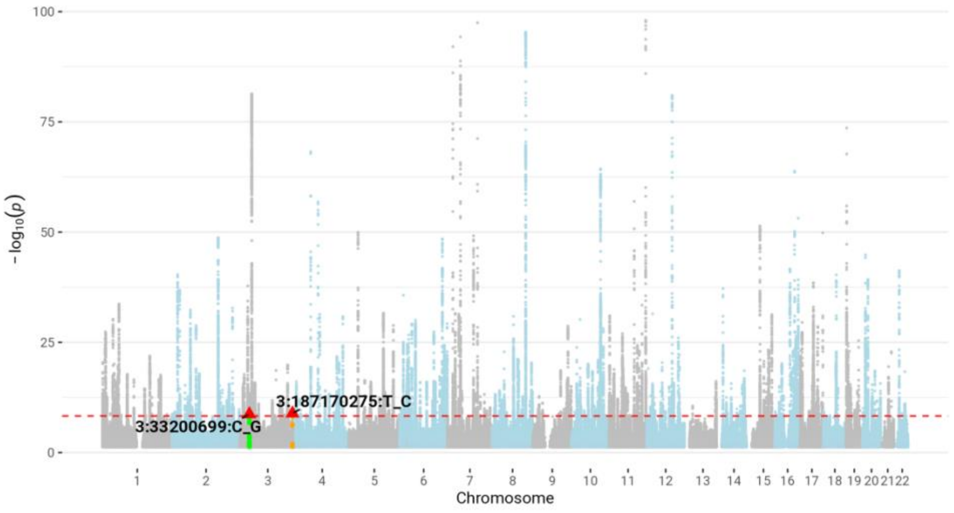


F. The 1df GxE test of PP-CPD of combined sex analyses in CPMA

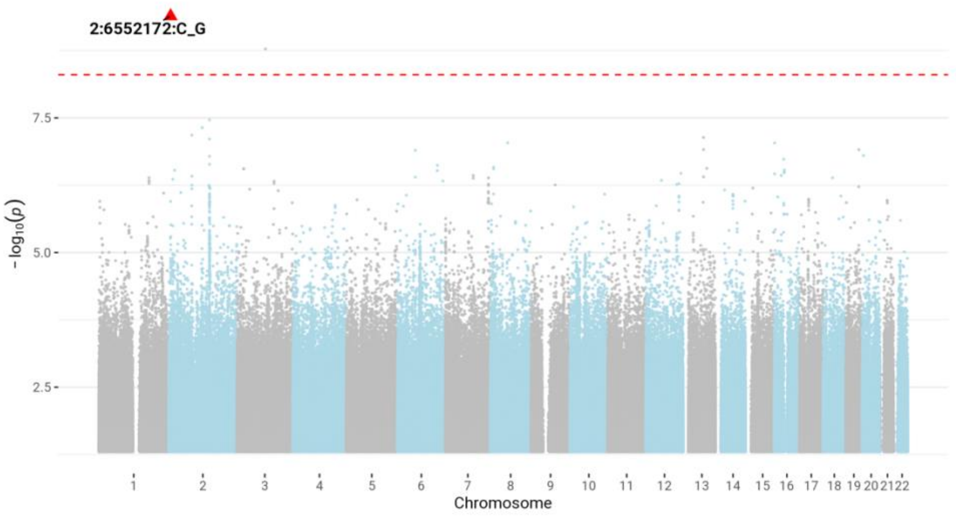


G. The 1df GxE test of SBP-CURSMK of combined sex analyses in CPMA

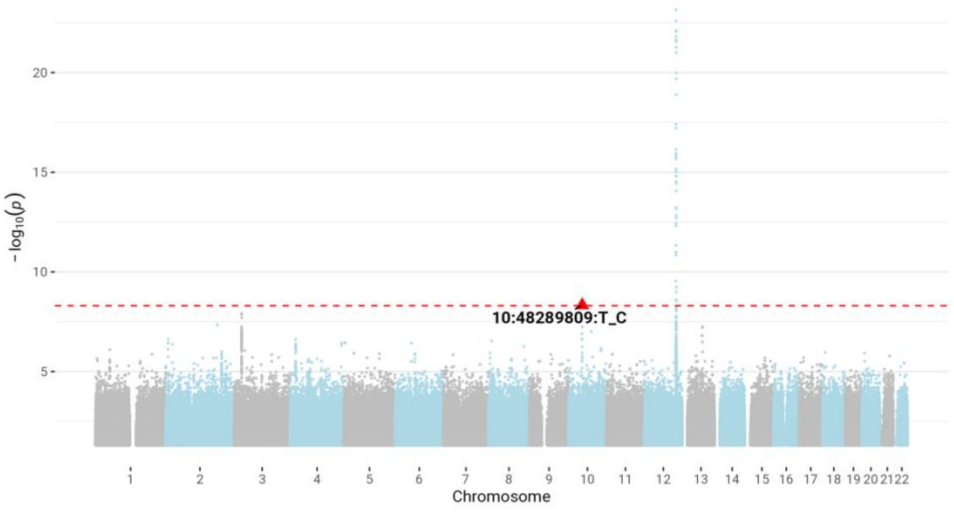

### Supplementary Figure S2. Miami plots and QQ plots

The QQ plots and Miami plots illustrate the association between smoking exposures, current smoker (CURSMK), cigarettes per day (CPD), and pack year (PY), and blood pressure traits in combined sex analyses. Population-specific results are shown for African (AFR), East Asian (EAS), European (EUR), Hispanic/Latino (HIS), and South Asian (SAS) populations, along with a cross-population meta-analysis (CPMA). In the Miami plots, the top panel displays the 1 degree of freedom GxE interaction p-values, while the bottom panel shows the 2 df joint p-values. A red line marks the threshold for genome-wide significance at $-\log_{10} 5 \times 10^{-9}$.

#### A. Cross-Population Meta-Analysis

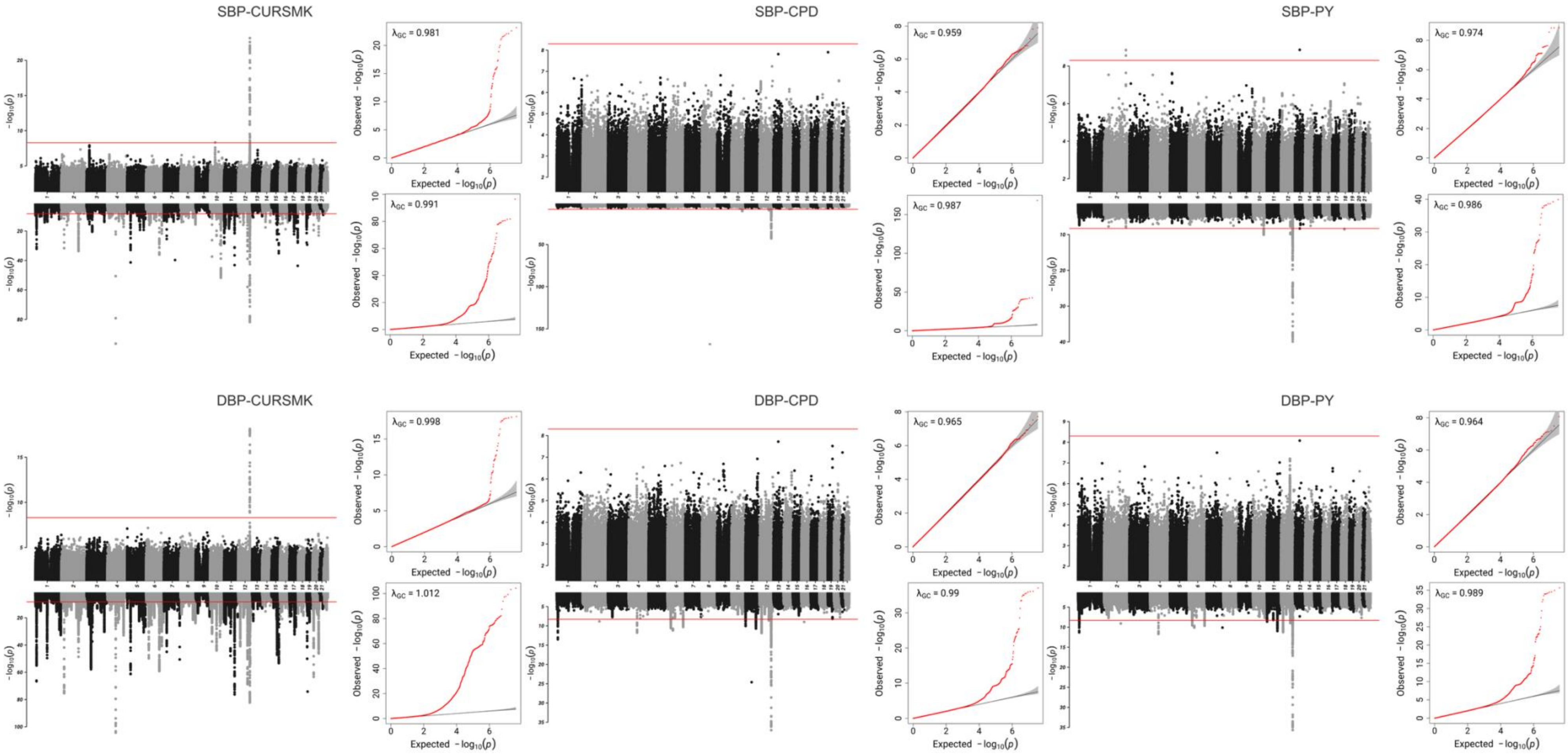

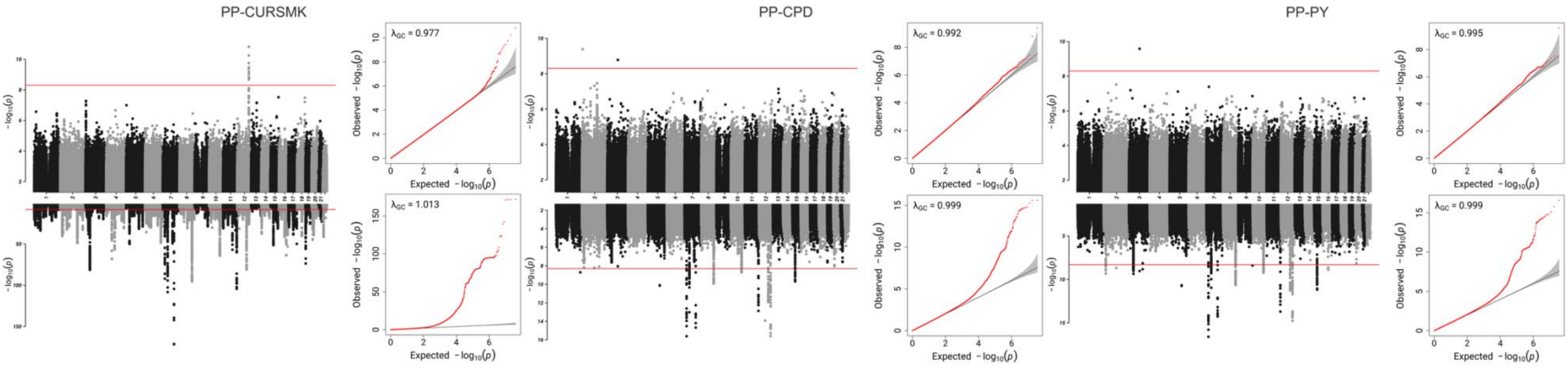


B. EUR-Specific Meta-Analysis

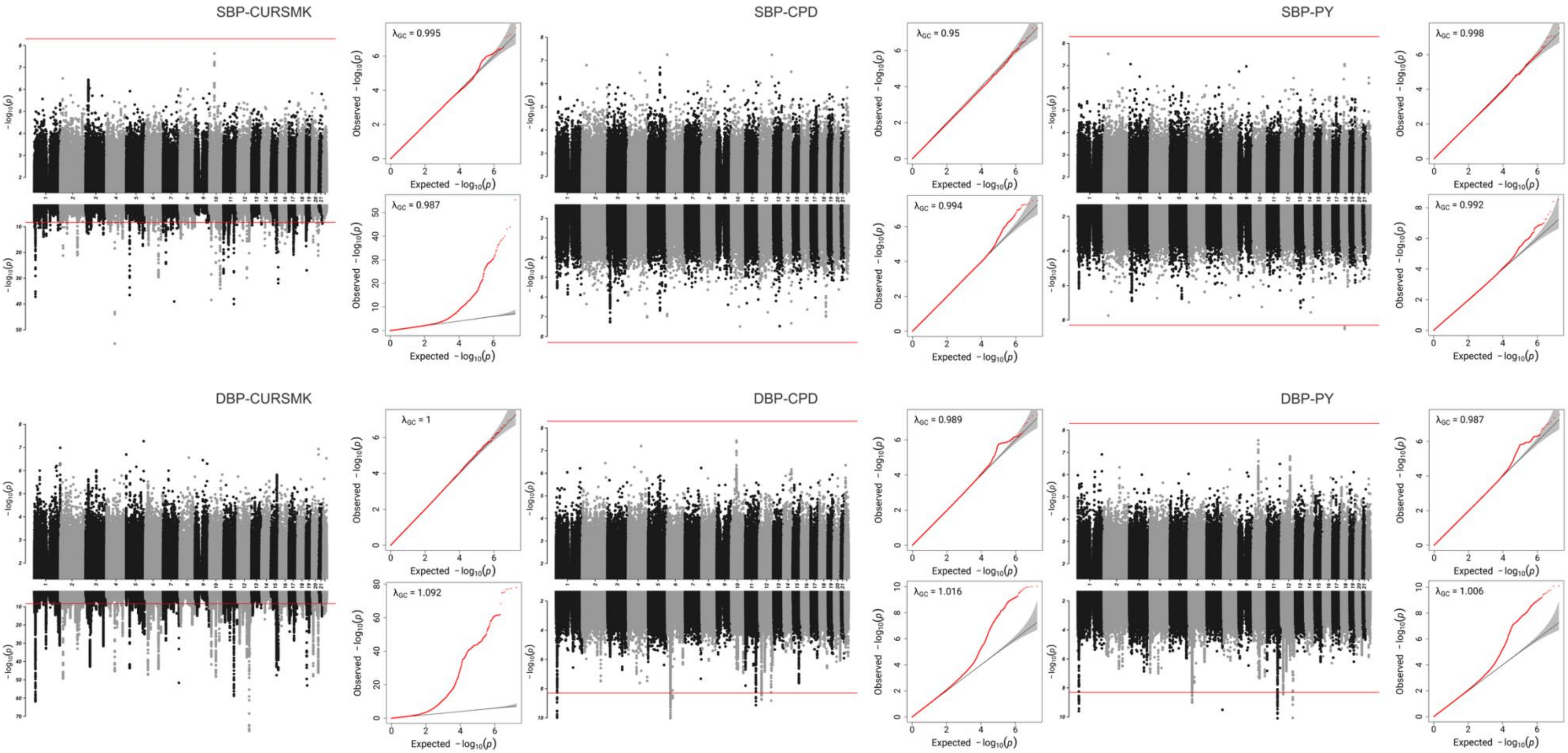

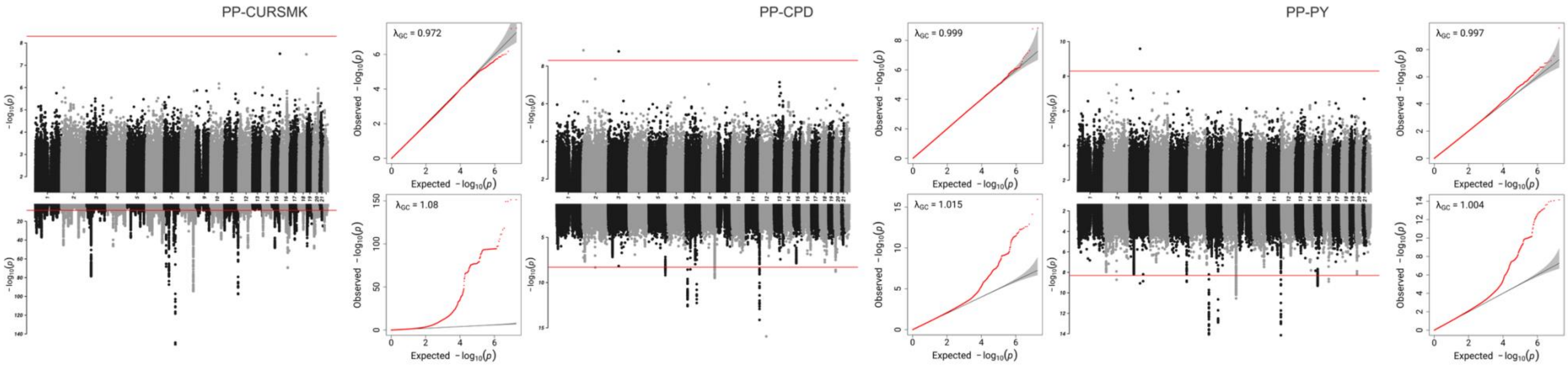


C. EAS-Specific Meta-Analysis

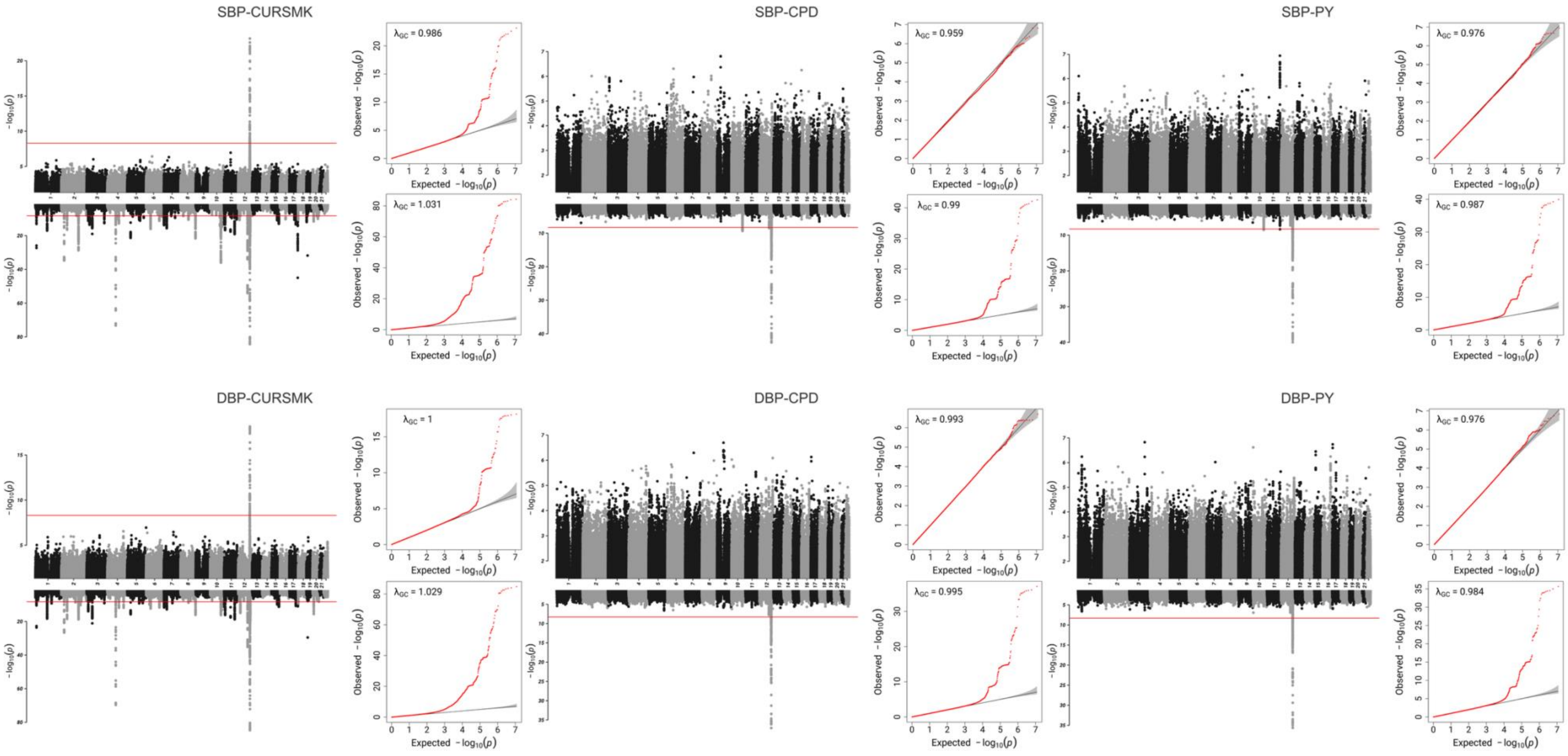

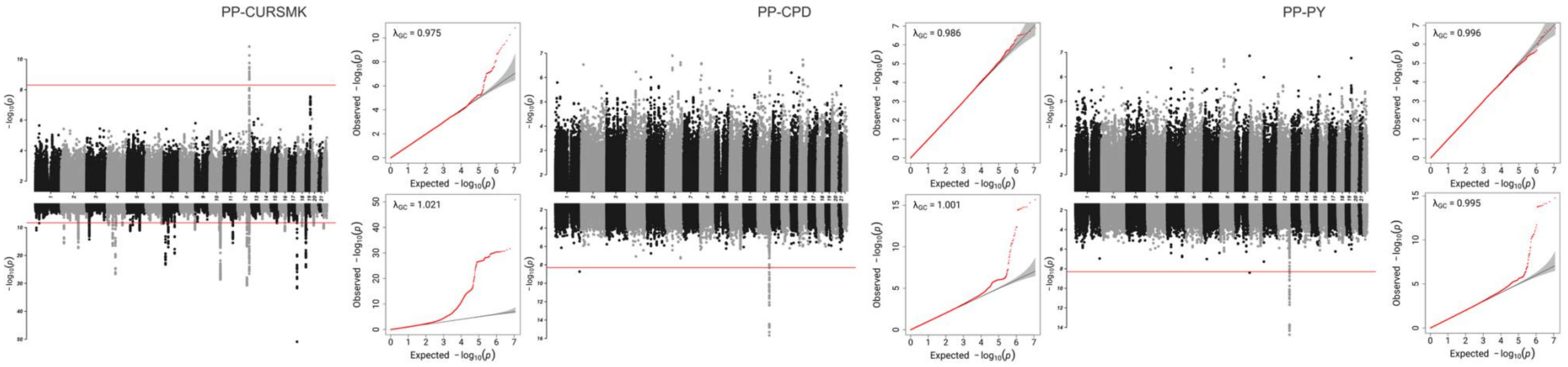


D. AFR-Specific Meta-Analysis

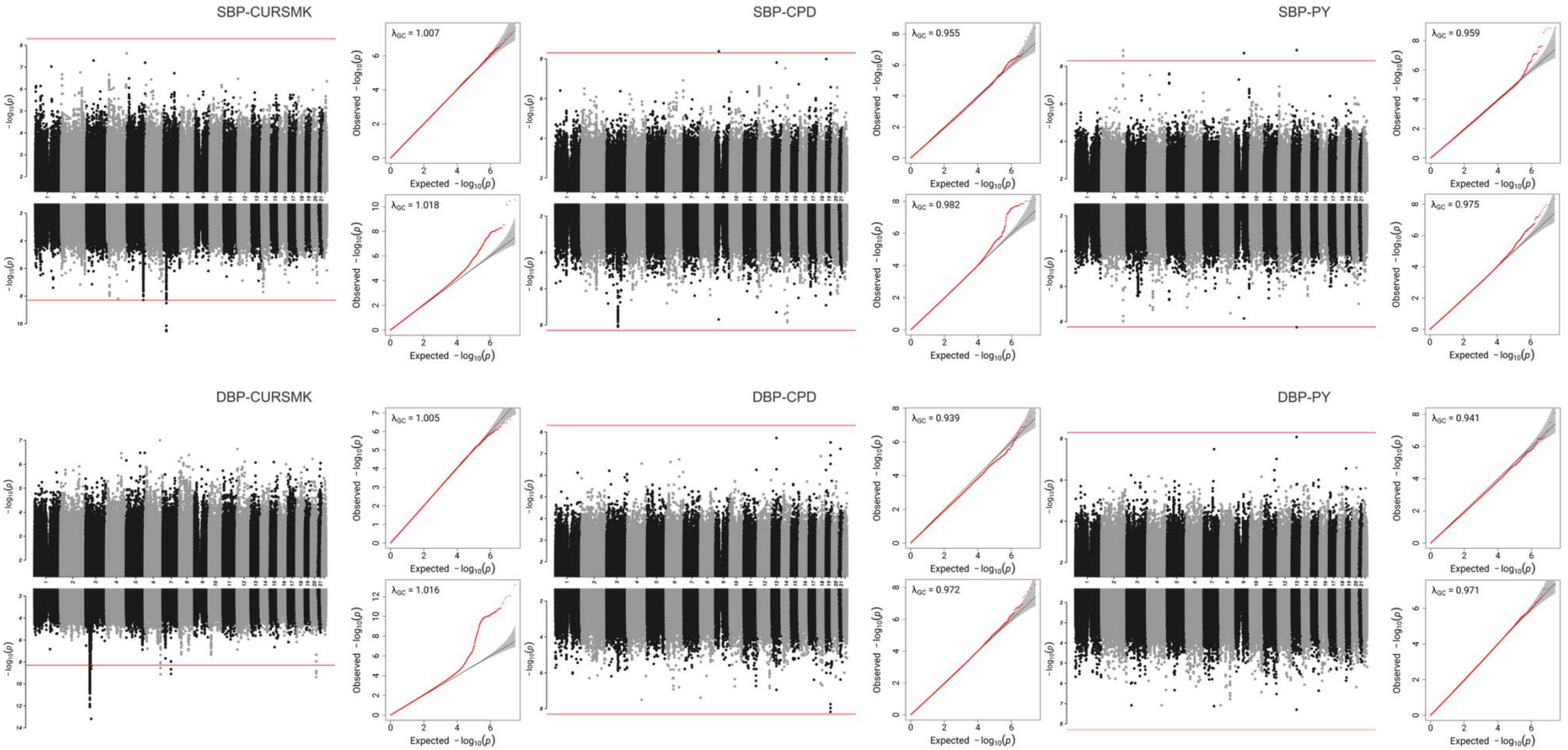

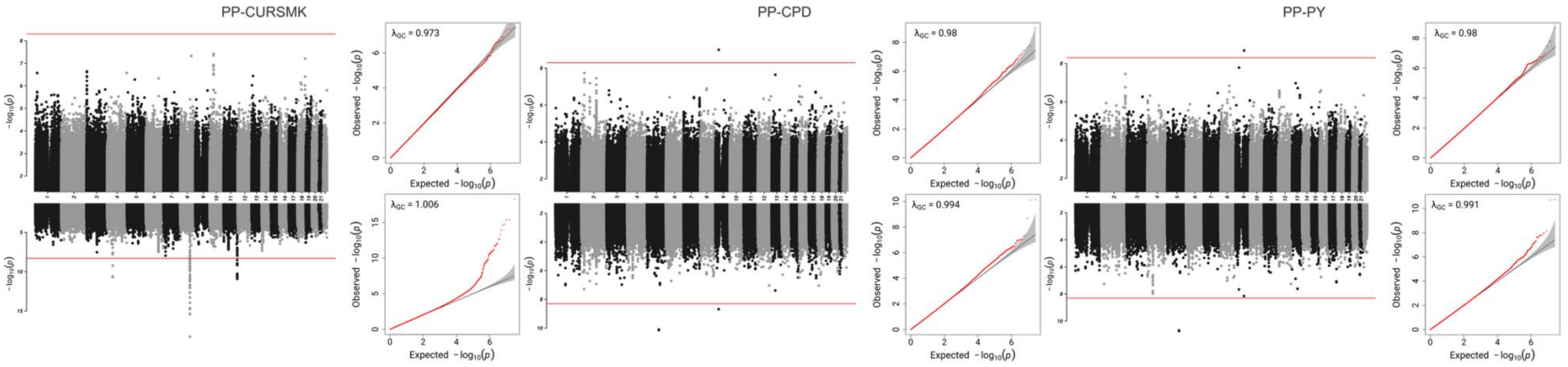


E. HIS-Specific Meta-Analysis

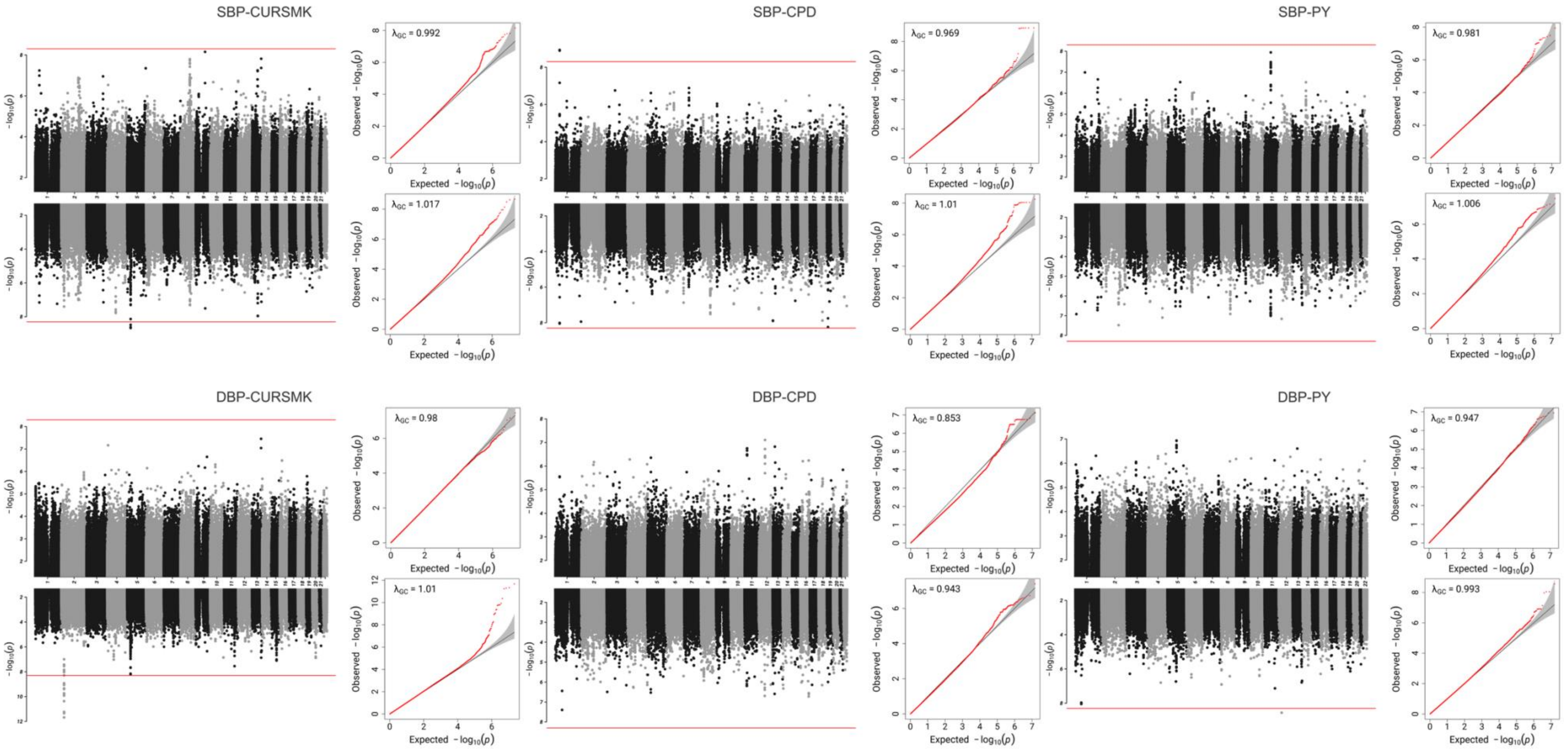

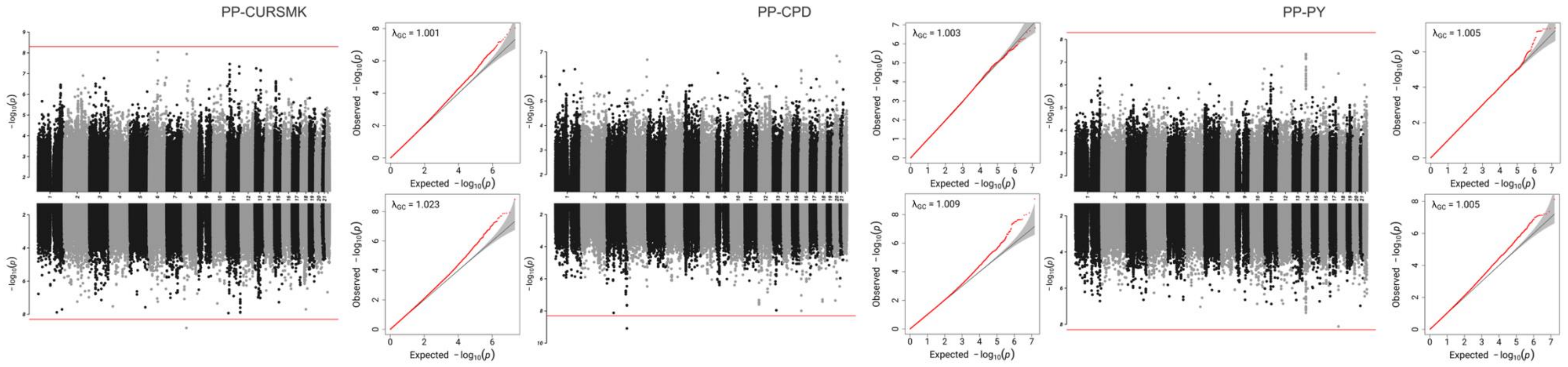


F. SAS-Specific Meta-Analysis

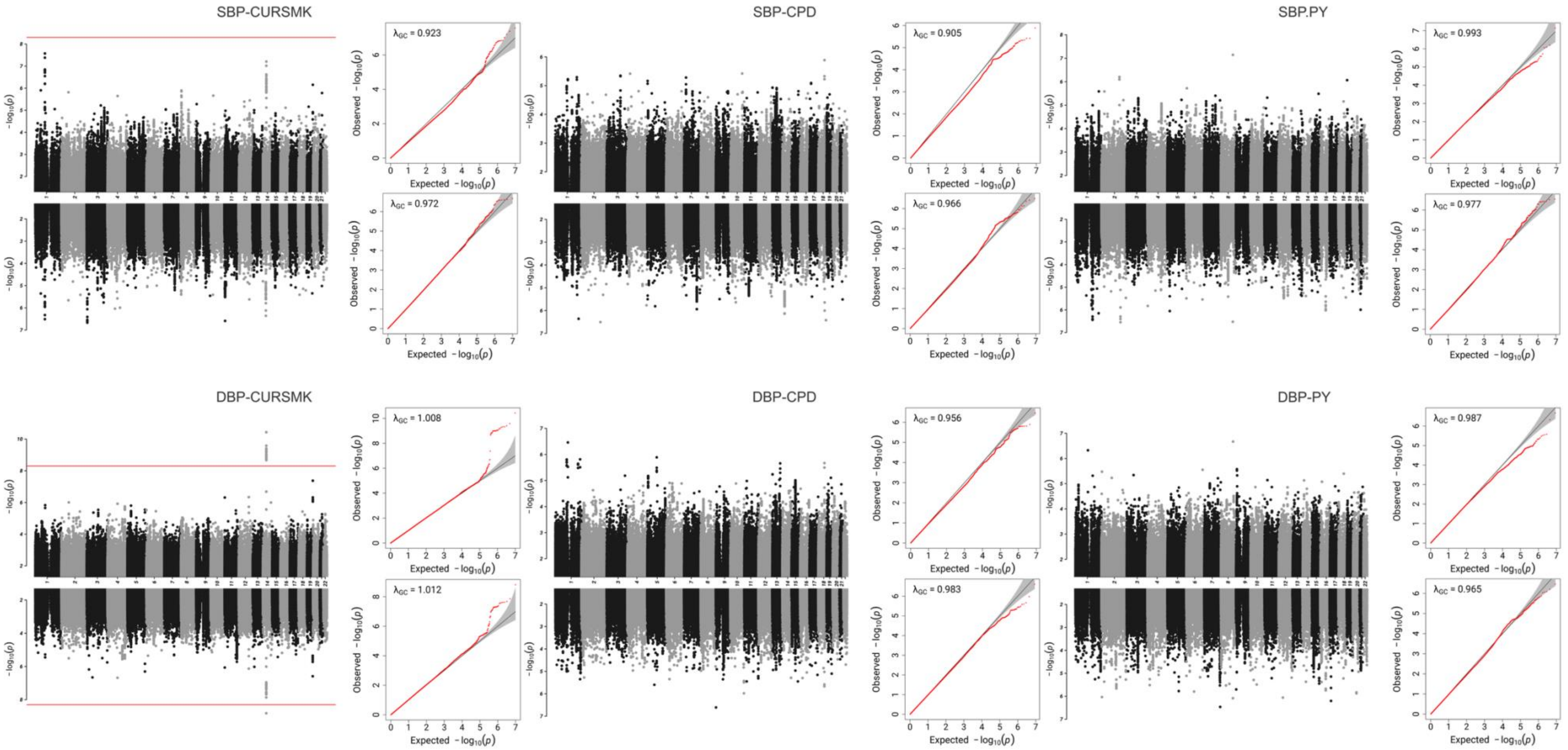

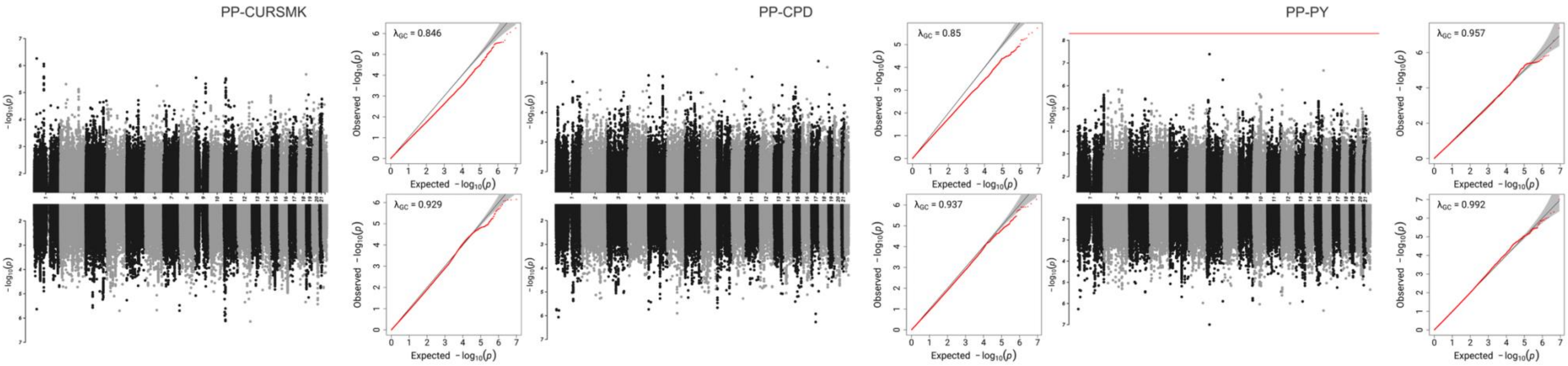
PP-CURSMK
PP-CPD
PP-PY
λGC = 0.846
λGC = 0.929
λGC = 0.85
λGC = 0.937
λGC = 0.957
λGC = 0.992
Observed −log10(p)
Expected −log10(p)
−log10(p)

#### Supplementary Figure S3. Novel loci gene-set enrichment from GENE2FUNC

A. Gene-set enrichment in molecular function for locus 9:95507136:CG_C identified in DBP-CURSMK CPMA 2df joint test in combined sex analyses.

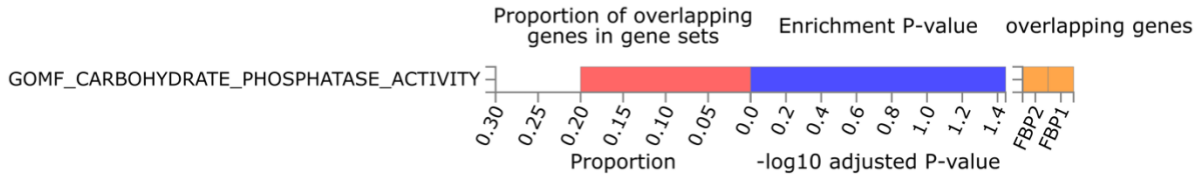


B. Gene-set enrichment in canonical pathway for locus rs8070260 identified in DBP-CURSMK CPMA 2df joint test in combined sex analyses.

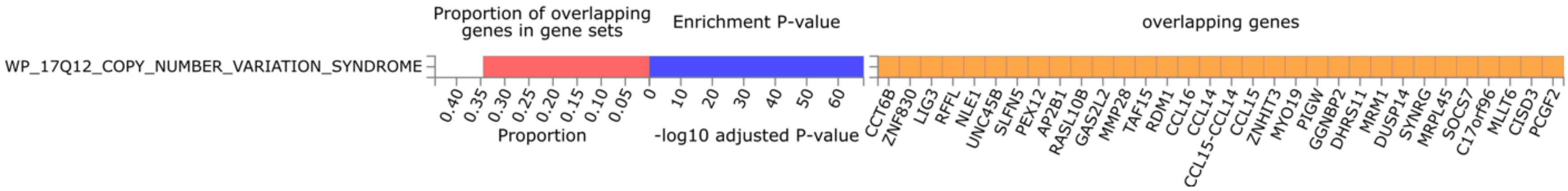


C. Gene-set enrichment in GWAS catalog reported genes for locus rs72832924 identified in DBP-CURSMK CPMA 2df joint test in males.

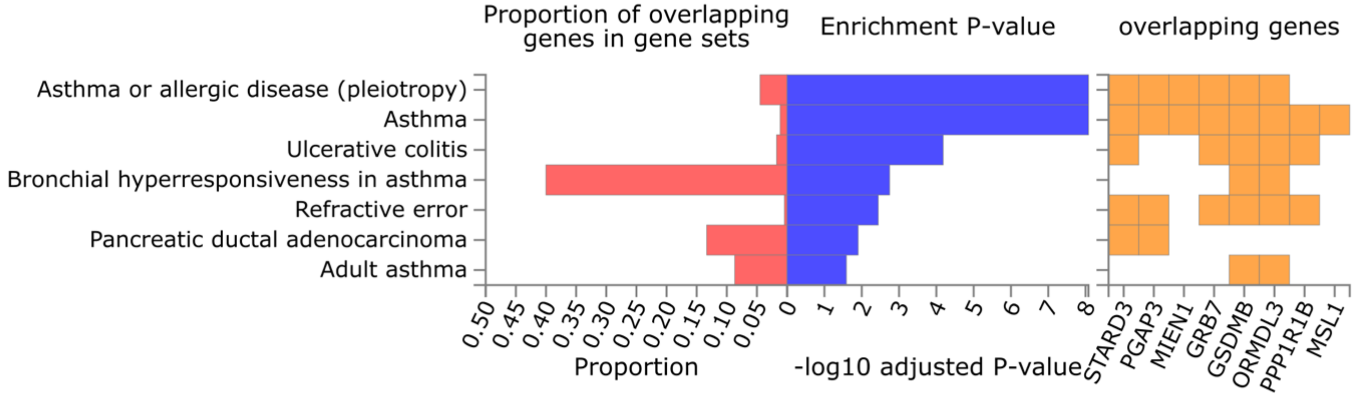

**Supplementary Figure S4. Gene-set enrichment from GENE2FUNC for causal SNPs from 2df fine-mapping**

A. Gene-set enrichment in canonical pathways for locus rs12476527 identified in DBP-CURSMK EAS and CMPA 2df joint test in combined sex analyses.

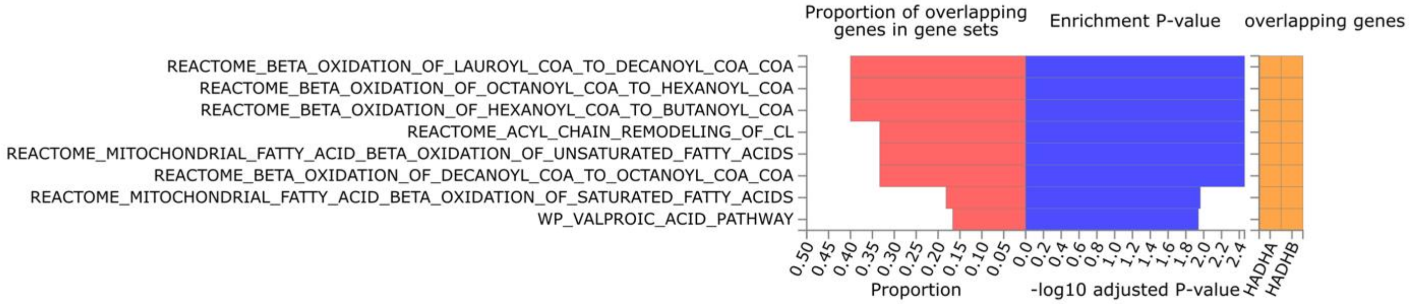


B. Gene-set enrichment in canonical pathways for locus rs8070260 identified in DBP-CURSMK CPMA 2df joint test in combined sex analyses.

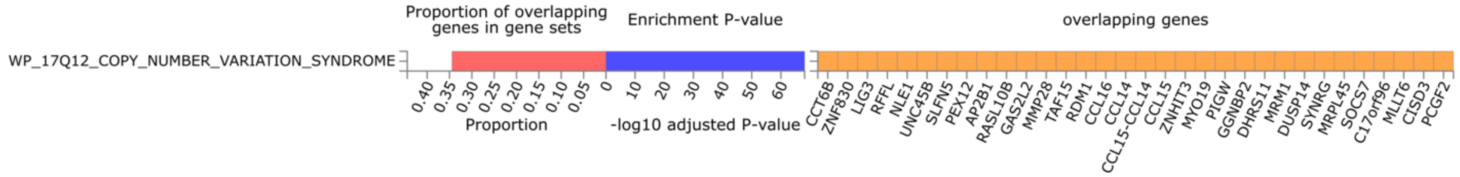

C. Gene-set enrichment in GWAS catalog reported genes for locus rs11066015 identified in DBP/SBP-CURSMK EAS 2df joint test in combined sex analyses.

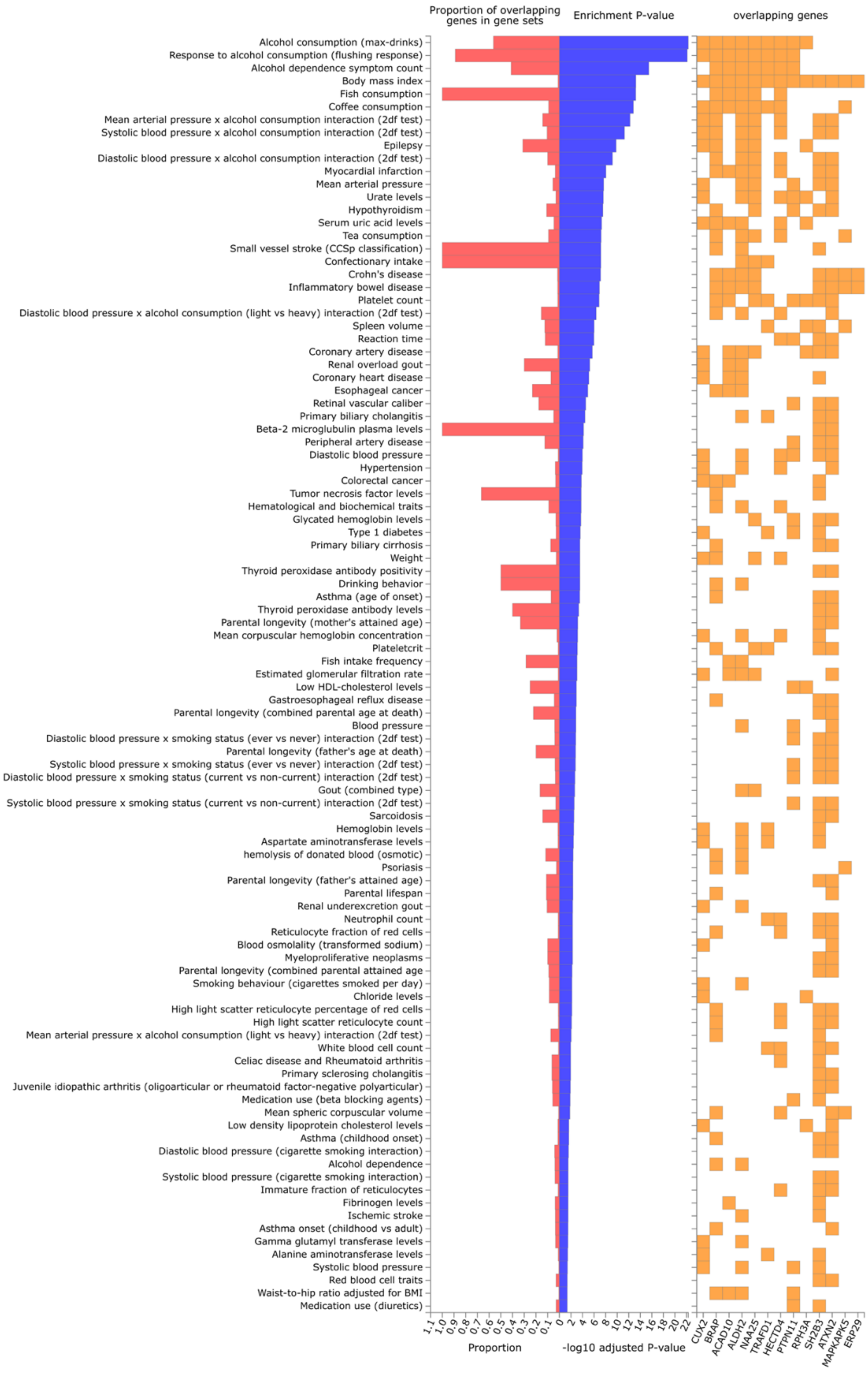

#### Supplementary Figure S5. Causal SNPs identified by two-degree-of-freedom fine-mapping.

Green dots were identified causal SNPs. **Panel A** showed causal SNPs on gene *MYO19* for gene-CURSMK interaction on blood pressure traits DBP in CPMA. **Panel B** showed causal SNPs on gene *MAEA* for gene-CURSMK interaction on blood pressure traits PP in CPMA. **Panel C** showed causal SNPs on *CCDC162P* for gene-CURSMK on trait DBP in CPMA. **Panel D** showed causal SNPs on *ASB3* for gene-CURSMK on trait PP in CPMA. **Panel E** showed causal SNPs on *ACAD10* for gene-CURSMK on trait SBP (left) and DBP (right) in EAS.

A. Novel locus rs8070260 on gene *MYO19*

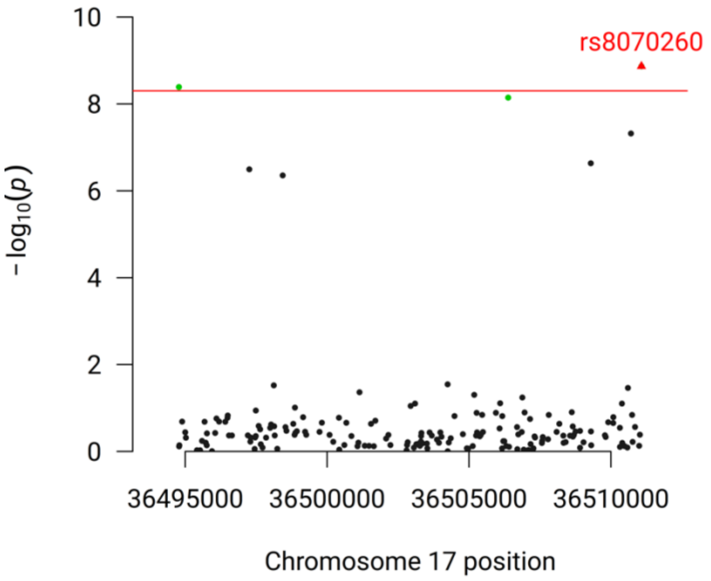


B. Locus rs60722337 on *MAEA*

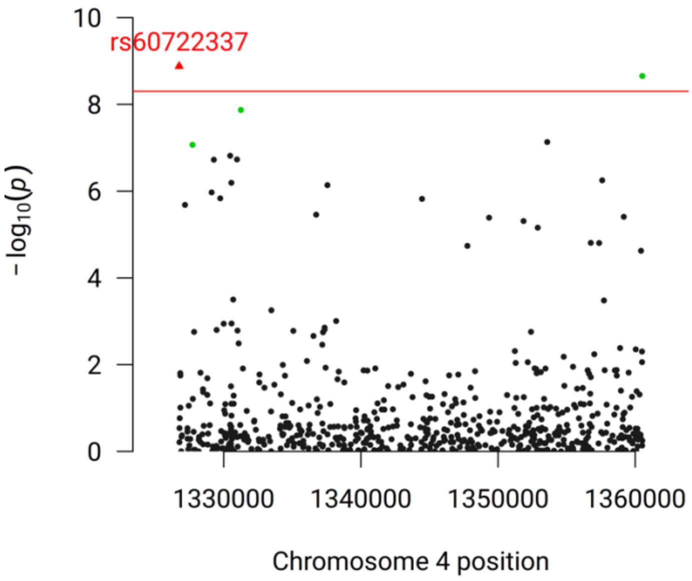


C. Locus rs6902892 on *CCDC162P*

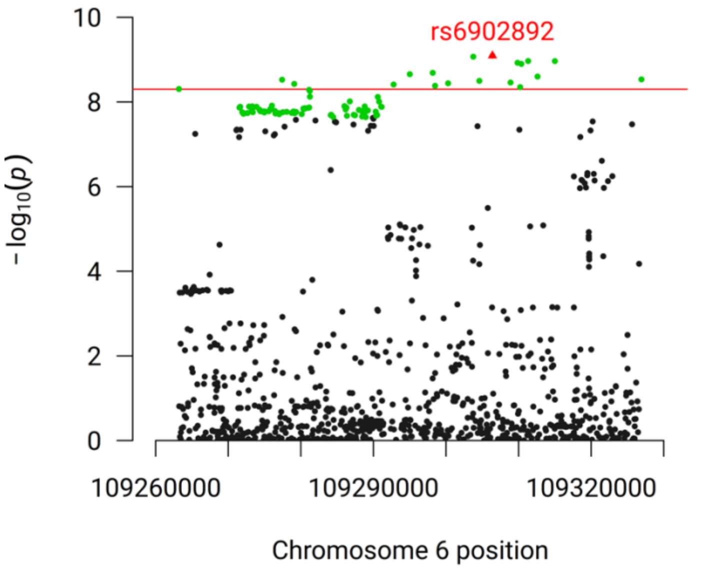


D. Locus rs74598641 on *ASB3*.

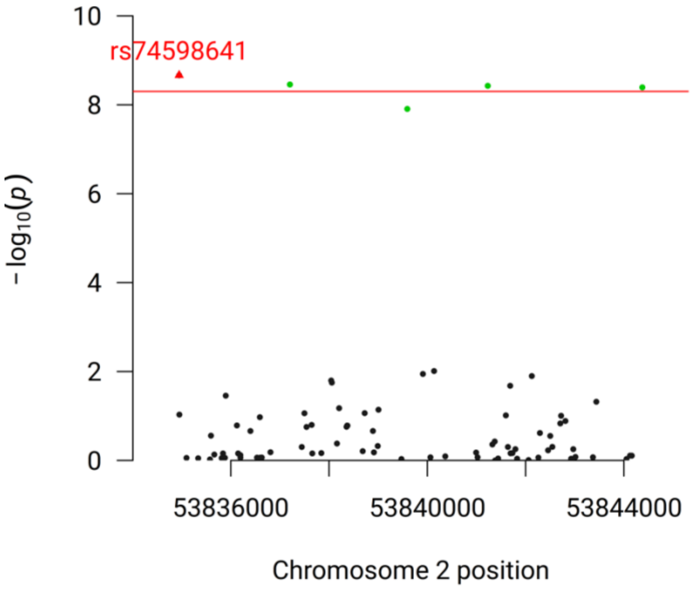

E. Locus rs11066015 on gene *ACAD10*

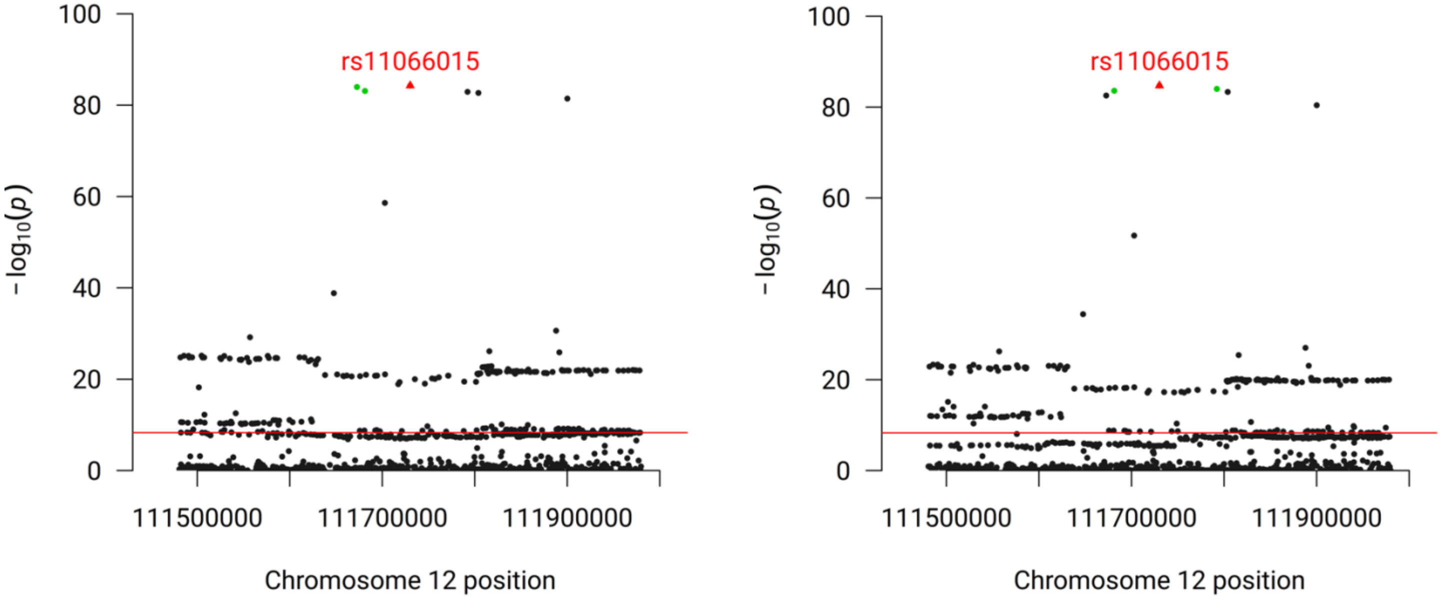
